# Influential drivers of the cost-effectiveness of respiratory syncytial virus vaccination in European older adults: A multi-country analysis

**DOI:** 10.1101/2024.08.06.24311440

**Authors:** Xiao Li, Lander Willem, Caroline Klint Johannesen, Arantxa Urchueguía-Fornes, Toni Lehtonen, Richard Osei-Yeboah, Heini Salo, Alejandro Orrico-Sánchez, Javier Díez-Domingo, Mark Jit, PROMISE investigators, Joke Bilcke, Harish Nair, Philippe Beutels

**Affiliations:** Centre for Health Economics Research and Modelling Infectious Diseases (CHERMID), University of Antwerp, Belgium; Department of Virology and Microbiological Special Diagnostics, Statens Serum Institut, Copenhagen, Denmark; Vaccine Research Department, Foundation for the Promotion of Health and Biomedical Research in the Valencian Region (FISABIO), Valencia, Spain; CIBER de Epidemiología y Salud Pública, Instituto de Salud Carlos III, Valencia, Spain; Finnish Institute for Health and Welfare, Helsinki, Finland; Centre for Global Health, The University of Edinburgh, Edinburgh, United Kingdom; Catholic University of Valencia, Valencia, Spain; Department of Infectious Disease Epidemiology & Dynamics, London School of Hygiene & Tropical Medicine

**Keywords:** RSV, respiratory, vaccination, policy, ageing population, economic evaluation, cost-utility analysis, cost-effectiveness analysis, uncertainty

## Abstract

**Background:** We aimed to identify influential drivers of the cost-effectiveness of older adult respiratory syncytial virus (RSV) vaccination in Denmark, Finland, the Netherlands and Valencia-Spain.

**Methods:** A static multi-cohort model was parameterised using country-and age-specific hospitalisations using three approaches: (1) the International Classification of Diseases (ICD)-coded hospitalisations, (2) laboratory RSV-confirmed hospitalisations and (3) time-series modelling (TSM). Plausible hypothetical RSV vaccine characteristics were derived from two protein subunit vaccines for adults aged ³60 years (“60y+”). Costs and quality-adjusted life-years (QALYs) were compared between four strategies: (a) “no intervention” and RSV vaccination in adults (b) 60y+; (c) 65y+; (d) 75y+, from both the healthcare payers’ and societal perspectives. Value of information, probabilistic sensitivity and scenario analyses identified influential drivers.

**Results:** Besides vaccine price, the hospitalisation estimates were most influential: Using adjusted RSV-ICD-coded hospitalisations at a vaccine price of €150 per dose, no intervention was cost-effective up to willingness-to-pay (WTP) values of €150 000 per QALY gained in Denmark and the Netherlands, and up to €125 000 per QALY gained in Finland. Using the adjusted RSV-confirmed dataset, the findings were consistent in Denmark and comparable in Finland. In Spain-Valencia, the 75y+ strategy became cost-effective at WTP >€55 000. Using TSM-based estimates, the 75y+ strategy was cost-effective at WTP >€45 000, >€101 000, >€41 000 and >€114 000 in Denmark, Finland, the Netherlands and Spain-Valencia, respectively. The (in-hospital) case fatality ratio and the specification of its age dependency were both influential. Duration of protection was found more influential than a variety of plausible waning patterns over the duration of protection.

**Conclusion:** Data gaps and uncertainties on the RSV-related burden in older adults persist and influence the cost-effectiveness of RSV vaccination. More refined age-and country-specific data on the RSV attributable burden are crucial to aid decision making.

## Introduction

Respiratory syncytial virus (RSV) is a leading cause of respiratory tract infections (RTI) in both young children and older adults. Among adults aged 60 years and above (60y+), a meta-analysis estimated 5.2 million RSV cases, 470 000 hospitalisations and 33 000 in-hospital deaths across high-income countries in 2019 [1]. With ageing populations in these countries, this disease burden is expected to increase.

RSV is contagious and seasonal, and it typically peaks during the winter months in Europe, in line with several other respiratory pathogens. RSV leads to a substantial burden, exerting pressure on healthcare resources [2]. The RSV-related disease burden among European older adults has not been well established, in terms of country-and age-specific hospitalisation rates, which are essential ingredients for cost-effectiveness analyses. Previous systematic reviews included multiple studies that reported hospitalisation rates, but the majority of these studies were conducted outside of Europe [1, 3].

In 2023, two protein subunit RSV vaccines, Arexvy^®^ (GSK) and Abrysvo^®^ (Pfizer) were approved for the prevention of RSV disease among adults aged 60y+ and marketed in several high-income countries. In 2024, an mRNA-based vaccine mResvia^®^ (Moderna) was also approved [4]. All vaccines have demonstrated protective efficacy lasting for more than one season, with waning immunity observed in the second season [5–8].

Recommendations on RSV immunisation in older adults were made by several National Immunisation Technical Advisory Groups. In the US, the Advisory Committee on Immunization Practices (ACIP) recommended a single-dose RSV vaccine for all adults 75y+ and adults aged 60-74 years who are at increased risk for severe RSV disease [9]. In the United Kingdom (UK), the Joint Committee on Vaccination and Immunisation (JCVI) advised a programme for older adults aged 75 years and above (75y+)[10]. Cost-effectiveness analyses were conducted to inform these ACIP and JCVI recommendations.

The PROMISE (Preparing for RSV Immunisation and Surveillance in Europe) Consortium (https://imi-promise.eu/) aimed to produce new evidence on the burden of RSV disease in Europe, before and after the emergence of severe acute respiratory syndrome coronavirus 2 (SARS-CoV-2) causing coronavirus disease 2019 (COVID-19). For instance, data on RSV International Classification of Diseases (ICD)-coded and RSV-confirmed hospitalisations were collected using national or regional registries in several countries. Moreover, time-series modelling (TSM) was employed to estimate the RSV-attributable hospitalisations for these countries.

We used the data on RSV hospitalisations generated by the PROMISE project to identify and explore the influential parameters and assumptions driving the cost-effectiveness of RSV vaccination strategies among older adults in four European countries: Denmark, Finland, the Netherlands and Spain-Valencia. The choice of these countries was guided by the timely availability of data of sufficient quality and detail within the PROMISE project (June 2024).

## Methods

### Cost-effectiveness model

We developed a static multi-cohort model (Figure 1) named Multi-Country Model Application for RSV Cost-Effectiveness policy for Adults (MCMARCELA). The modelled population consists of adults aged 60 to 99 years, who were tracked monthly over a five-year time horizon, which aligns with the expected maximum protective duration of RSV vaccine [11]. The model considered costs and quality-adjusted life-years (QALYs) lost for RSV cases seen in primary care, admitted to hospitals, and symptomatic RSV cases requiring no medical attendance (non-MA). It also assumed that RSV-related mortality occurs only in a hospital setting. Premature RSV-related deaths were calculated from the model using hospital-fatality ratios to estimate QALYs lost over the remaining life-expectancy at the age of death. The model did not separately account for RSV cases requiring only an outpatient visit in hospital or an emergency department visit without admission, because such type of data was not available for most of the countries considered.

**Figure 1:**
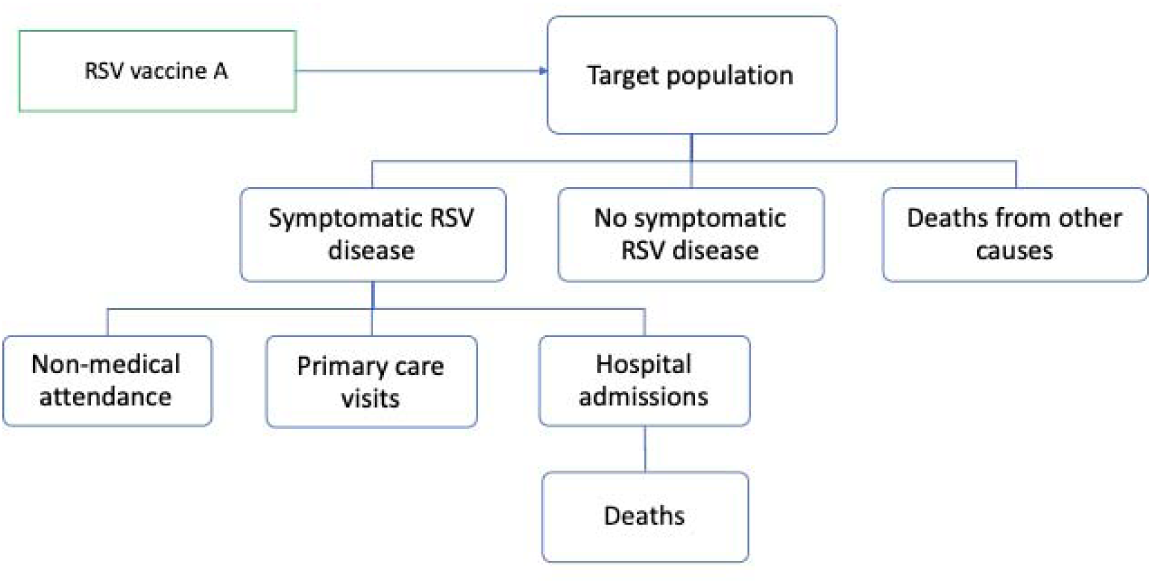
Model structure. Footnote: primary care visits were estimated from hospitalisations, and non-medical attendance were estimated from primary care visits.

The model made comparisons between four strategies: “no intervention” and three age-based single-dose universal RSV vaccination strategies among older adults. Each vaccination strategy was modelled as if a single dose were offered to everyone in an age band at the same time in October, prior to the start of the first RSV season considered over the 5-year time horizon:

1. 60 years and above up to 99 years (‘60y+’ strategy) based on the approved age-indication,
2. 65 years and above up to 99 years (‘65y+’ strategy) considering the universal influenza vaccination programmes in many European countries,
3. 75 years and above up to 99 years (‘75y+’ strategy) based on the JCVI and ACIP recommendation.

This analysis was conducted separately from a healthcare payers’ (HCP) perspective considering only direct medical costs and from a societal perspective, additionally including costs associated with productivity losses of patients. Both costs and QALYs were discounted at rates recommended by the country-specific pharmacoeconomic guidelines (see Supplement Table 6) [12–15].

### Model input parameters and assumptions

All parameters are listed in Supplement Table 6, and each input parameter is described in detail in Supplement section 1.1.

In brief, for Denmark, Finland and the Netherlands, RSV-ICD-coded hospitalisations by age and calendar month were estimated retrospectively using national registries. The RSV-confirmed data were also obtained in Denmark and Finland using the same registries [16, 17]. In Valencia, a region of Spain, the RSV-confirmed hospitalisations were collected retrospectively through their regional active hospital-based surveillance network (descriptions in previous publications [18, 19]). The catchment area represented 21% of the overall population in Valencia (approximately 1 million), or 0.2% of the population in Spain. Patients meeting preliminary inclusion criteria and meeting the influenza-like-illness (ILI) case definition [20] were included and systematically tested for RSV with a multiplex polymerase chain reaction (PCR) test. RSV-confirmed data was adjusted to the RSV season as active surveillance was not performed throughout the whole year [16, 21]. The brief description of the data collection methods is in Supplement section 1.1.1, and the full details are available elsewhere [16, 17].

Subsequently, a TSM analysis was conducted to attribute respiratory tract infection (RTI) hospital admissions to RSV, using the number of positive virological isolates of respiratory pathogens, including influenza A and B, SARS-CoV-2, and RSV, over time as covariates. Under this approach influenza and RSV laboratory test frequency was regressed against RTI incidence over time, to determine the proportion of RTI cases that were attributable to the two viruses [22]. In Spain-Valencia, ILI was used instead of RTI in the TSM analysis and an inference on missing weeks without active surveillance was performed in order to account for the fact that data was not available throughout the whole year. The brief description of the TSM analysis is in Supplement section 1.1.1.3 and the full details are available elsewhere [21].

To account for uncertainty on the choice of source data for the hospitalisation estimates, we explored the cost-effectiveness of using

1. the RSV-ICD-coded hospitalisations with an adjustment factor of 2.2 for diagnostic under-ascertainment, except for Spain-Valencia due to insufficient sample size [23].
2. the RSV-confirmed hospitalisations with the adjustment factor of 2.2 (except for the Netherlands due to data unavailability),
3. the TSM estimated RSV-attributable hospitalisations.

The differences among the three hospitalisation datasets were illustrated in Supplement Figures 1-4. We used the average data over three or four seasons (depending on data availability) prior to the COVID-19 pandemic (2016/2017 to 2019/2020).

The RSV-related primary care episodes by age were estimated based on a meta-analysis of US studies [24], which found for every RSV hospitalisation in adults aged 60y+, 8.5 RSV primary care episodes occurred on average. We additionally used higher estimates of RSV-related primary care episodes based on a UK modelling study in scenario analysis [25]. A multi-country prospective study among European community-dwelling older adults provided an estimate of 2.27 non-MA episodes per primary care episode [26, 27]. We used 7.13% (95% CI: 5.4-9.36) for the in-hospital case fatality ratio (hCFR) for all 60y+ based on a meta-analysis [1]. The age-specific RSV-confirmed hCFRs in Finland were used in scenario analysis.

In order to focus on identifying general drivers of cost-effectiveness of RSV vaccination among older adults, and to avoid detractions due to minute interpretative differences in differently defined product-specific clinical endpoints, we consider in this analysis the use of a hypothetical RSV vaccine. We used plausible efficacy (mean and 95% CI) values against non-MA episodes, primary care episodes, hospitalisations, and deaths by combining information available at the time of the analysis for both Arexvy^®^ and Abrysvo^®^ (Table 1, Supplement 2.1.5) [5, 6]. Based on waning assumptions made in published cost-effectiveness analyses for older adults [28–32], we explored 13 waning scenarios accommodating a range for the duration of protection from 18 months to 48 months (Supplement section 1.15). The RSV vaccine price was assumed to be €150 per dose for all countries in this study [33]. RSV vaccination coverage was based on country-and age-specific influenza vaccination coverage (Supplement Table 6).

**Table 1:**
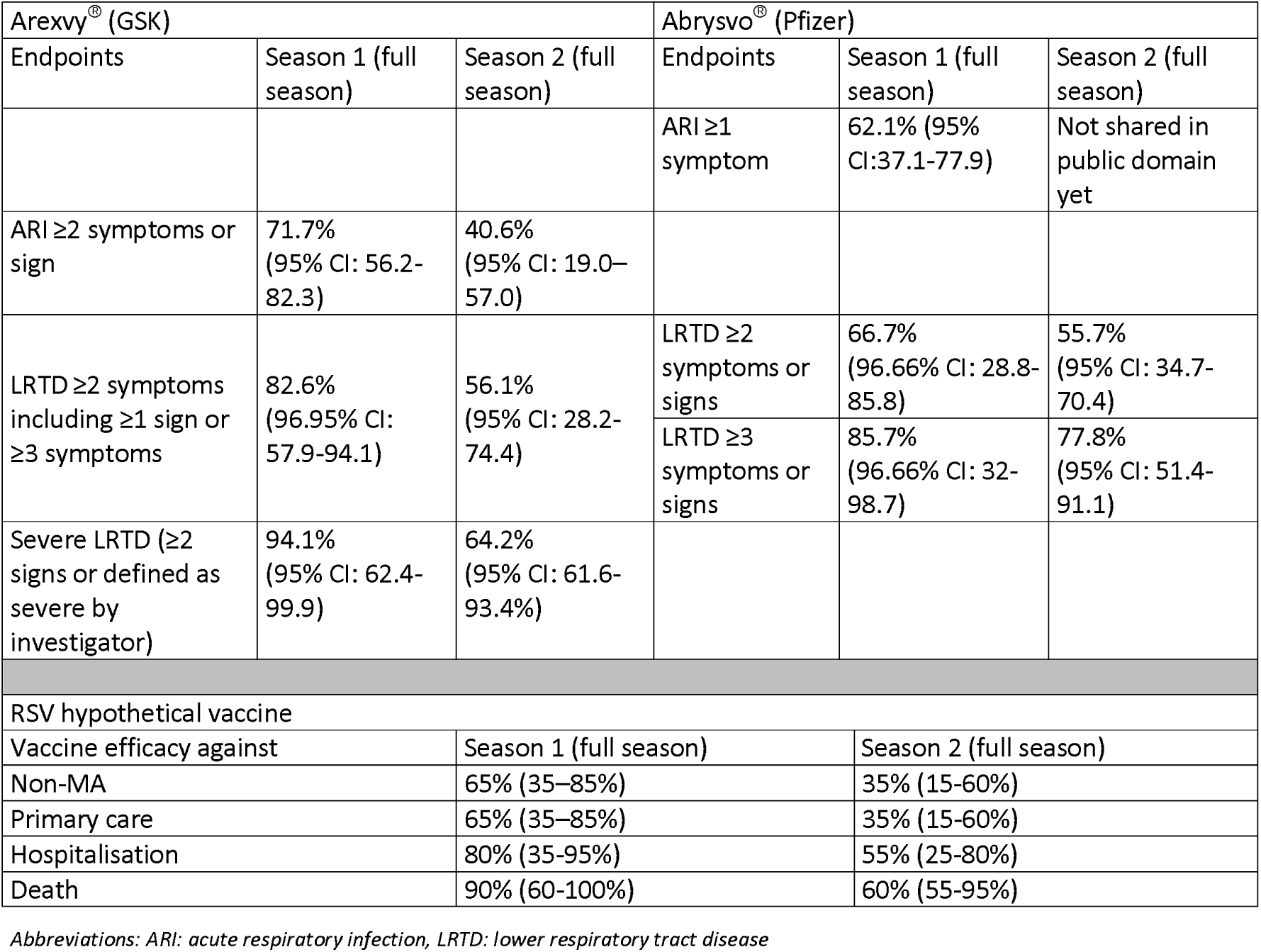
Phase 3 vaccine efficacy data of Arexvy^®^ and Abrysvo^®^ and the efficacy values of an RSV hypothetical vaccine.

Country-specific costs were obtained for hospitalisations (including intensive care unit admissions), primary care consultations, and vaccine administration (Supplement section 1.1.7). A cost of €4.06 per non-MA episode was used for all countries based on a European multi-country prospective study [27]. From a societal perspective, the costs of productivity losses were estimated using the human capital approach: the age-and country-specific daily salaries were adjusted by employment rates and multiplied by the number of workdays lost. We accounted for one and two lost workdays per non-MA and primary care episode, respectively [34], and the country-specific length of hospital stay plus three days per hospitalised case. Productivity losses due to RSV-associated premature deaths were notincluded in order to avoid double counting [35]. All costs were inflated to their 2023 value using the country-specific consumer price index (CPI) of all sectors, and those reported in local currency were converted to euro (€) using the annual exchange rates of 2023 [36, 37].

QALY losses due to RSV deaths were estimated using remaining life expectancy at age of death and standard age-specific utility values by country [38, 39] (Supplement section 1.1.8 and Table 6). A prospective study in European older adults estimated an average QALY loss of 0.004 (95% CI: 0.001-0.010) per non-MA or primary care episode [34]. There was no utility value available for individuals aged 60y+ who were hospitalised with RSV. Hence, we assumed QALY losses of 0.01023 (95% CI: 0.0089-0.0170) based on another European multi-country prospective study among RSV-hospitalised infants’ parents [40], this value was comparable to QALY losses in hospitalised influenza patients [41]. However, in scenario analysis, we also assumed alternative average QALY losses of 0.0185 (CI: 0.0053-0.0347) for non-hospitalised patients and 0.0193 (CI: 0.0095-0.0316) for hospitalised patients, based on a 2022 US ACIP meeting, subsequently published in Hutton et al [42].

### Analytical approach

We conducted a full incremental analysis of costs and effects arising in all strategies to identify the cost-effective program for willingness-to-pay (WTP) values ranging from €0 to €150 000 per QALY gained [43, 44].

To evaluate the impact of uncertain input parameters on the estimated cost-effectiveness, the expected value of partial perfect information (EVPPI) was calculated over a range of WTP values through probabilistic sensitivity analysis. A probabilistic price threshold analysis was also performed for each country applying the method demonstrated by Pieters and colleagues [45].

To explore the impact of key assumptions and input parameters, we also conducted deterministic scenario analyses including 13 scenarios on vaccine characteristics (i.e., efficacy, duration of protection, waning curves), and 16 scenarios on input parameters such as burden of disease, QALY losses, RSV seasonality, price per dose, and the impact of the COVID-19 pandemic. An overview of scenarios analyses was summarised in Supplement Tables 1 and 7 and section 1.4. All analyses were conducted in R version 4.3.2.

## Results

### RSV health and economic burden without vaccination

Over a five-year time horizon, (1) using the adjusted RSV-ICD-coded dataset (Supplement Table 10), we estimated 30 910, 252 679, and 316 814 RSV cases (including non-MA, primary care episodes and hospitalisations), as well as 618, 4 780, and 8 905 discounted QALY losses among adults 60y+ in Denmark, Finland and the Netherlands, respectively. From the HCP and societal perspectives, the discounted costs were €3.8 and 4.3 million in Demark, €32.6 and €35.2 million in Finland, and €80.3 and €95.3 million in the Netherlands, respectively.

(2) Using the adjusted RSV-confirmed hospitalisation dataset, the estimated RSV cases were 84 852 in Denmark, 326 588 in Finland, and 74 083 in Spain-Valencia, as well as 1 677, 6 223, and 1 615 discounted QALY losses among adults 60y+ in Denmark, Finland and Spain-Valencia, respectively. From the HCP and societal perspectives, the discounted costs were €10.5 and 11.7 million in Demark, €42.2 and €45.6 million in Finland, and €11.0 and €11.3 million in Spain-Valencia, respectively.

(3) Using the TSM RSV-attributable estimates, we observed 24-and 9-fold higher RSV-attributable cases (0.7 million) in Demark compared with the adjusted RSV-ICD-coded and adjusted RSV-confirmed hospitalisations, respectively. Consequently, there were higher QALY losses and direct medical and indirect medical costs. In the Netherlands, 4-fold higher RSV-attributable cases (1.3 million) were estimated compared with using the adjusted RSV-ICD-coded dataset. However, both in Finland and Spain-Valencia, the RSV-attributable cases from the TSM were 11% (289 516) and 49% (37 895) lower than the adjusted RSV-confirmed estimates, respectively. Consequently, large differences were observed in the discounted QALY losses, and the direct and indirect costs attributable to RSV. Despite the large differences in baseline disease burden while using different datasets, it was consistent that the 75-84y age group had the highest number of hospitalisations (except the Netherlands using adjusted RSV-ICD-coded dataset); consequently, the highest number of primary care and non-MA episodes, and discounted medical costs. However, the highest discounted QALY losses mainly occurred in the 65-74y age group (except Spain-Valencia). Most indirect costs due to productivity losses occurred in the 60-64y age group, as it had the highest active labour force participation.

### RSV-burden averted with single-dose one-time vaccination over two years protection

Irrespective of the hospitalisation data source, in every country the 60y+ strategy had the greatest impact on disease burden in terms of cases, hospitalisations and deaths averted, QALYs gained, and direct and indirect medical costs averted. However, this was also the most expensive strategy with one-time vaccination costs of €195 million, €139 million, €482 million and €30 million in Denmark, Finland, the Netherlands, and Spain-Valencia, respectively. Detailed RSV burden averted estimates are presented in Supplement Tables 9, 11 and 13 by strategy, country and methodological approach.

### Cost-effectiveness

In general, more expansive vaccination strategies became cost-effective for the HCP with increasing WTP values per QALY gained (Figures 2-4). A societal perspective increased the potential cost savings per averted case, but the overall results remained comparable with the HCP perspective (Supplement Figures 8, 11, 14 and 15).

1. Using the adjusted RSV-ICD-coded hospitalisations (Figure 2), ‘no intervention’ was cost-effective up to a WTP value of €150 000 per QALY gained for the HCP in Denmark and the Netherlands. In Finland, the 75y+ would be preferred at WTP values exceeding €124 000.
2. Using the RSV-confirmed hospitalisations (Figure 3), the finding was consistent for Denmark that ‘no intervention’ is cost-effective, whereas in Finland the 75y+ strategy became preferred at lower WTP values (≥95 000) versus using adjusted RSV-ICD-coded hospitalisations. In Spain-Valencia, the 75y+ and 65y+ strategies became cost-effective at WTP values above €55 000 and €116 000, respectively.
3. Using the TSM estimates (Figure 4), the 75y+ strategy was preferred if the WTP value exceeded €45 000 in Denmark, €101 000 in Finland, €41 000 in the Netherlands, and €114 000 in Spain-Valencia. The 65y+ strategy became the preferred strategy at higher WTP values of €65 000 in Denmark, and €70 000 in the Netherlands. In Finland and Spain-Valencia, 65y+ strategy was not preferred up to a WTP value of €150 000.

**Figure 2:**
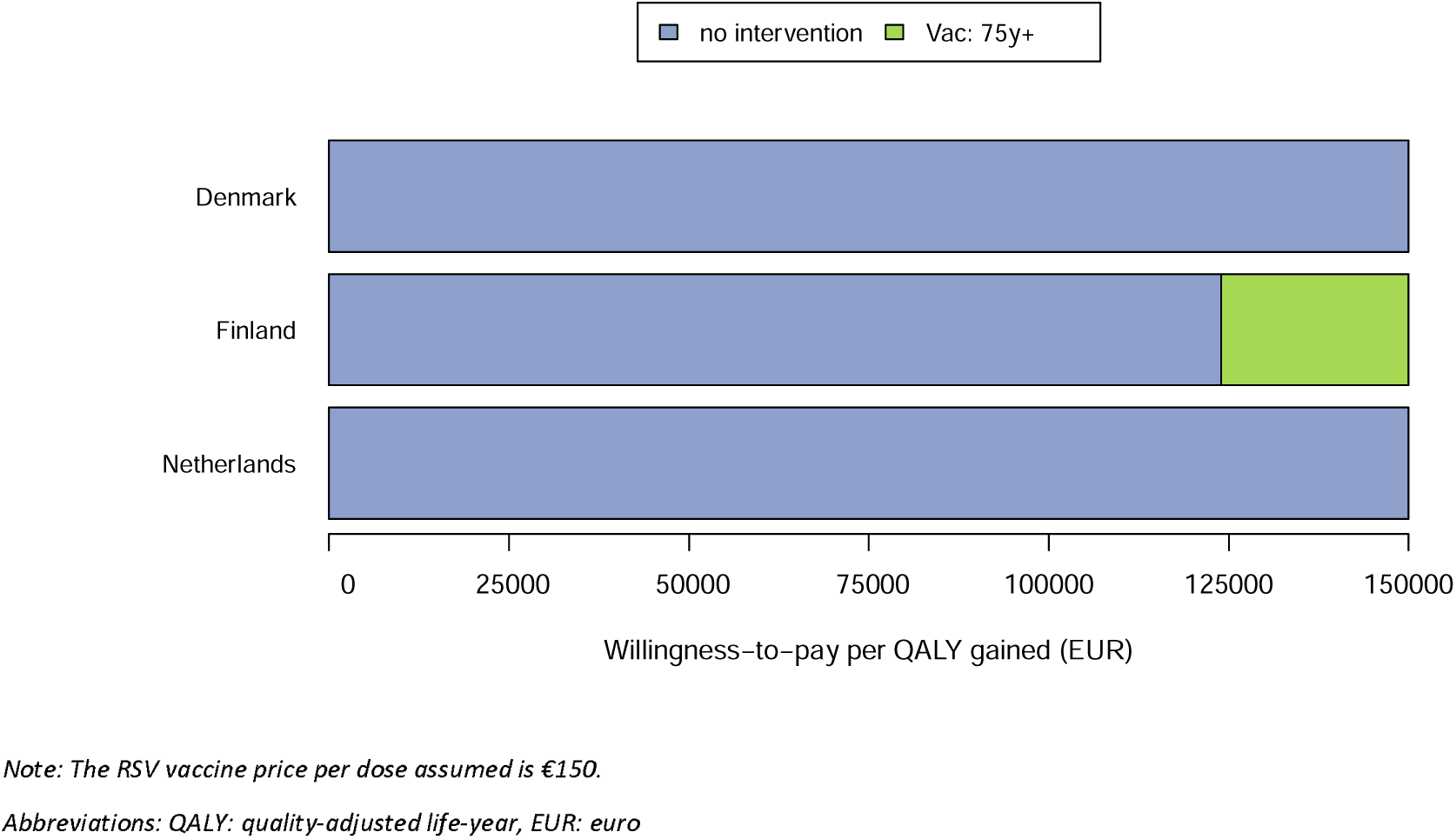
Using adjusted RSV-ICD-coded disease burden: preferred strategy based on cost-effectiveness analysis from the healthcare payers’ perspective.

**Figure 3:**
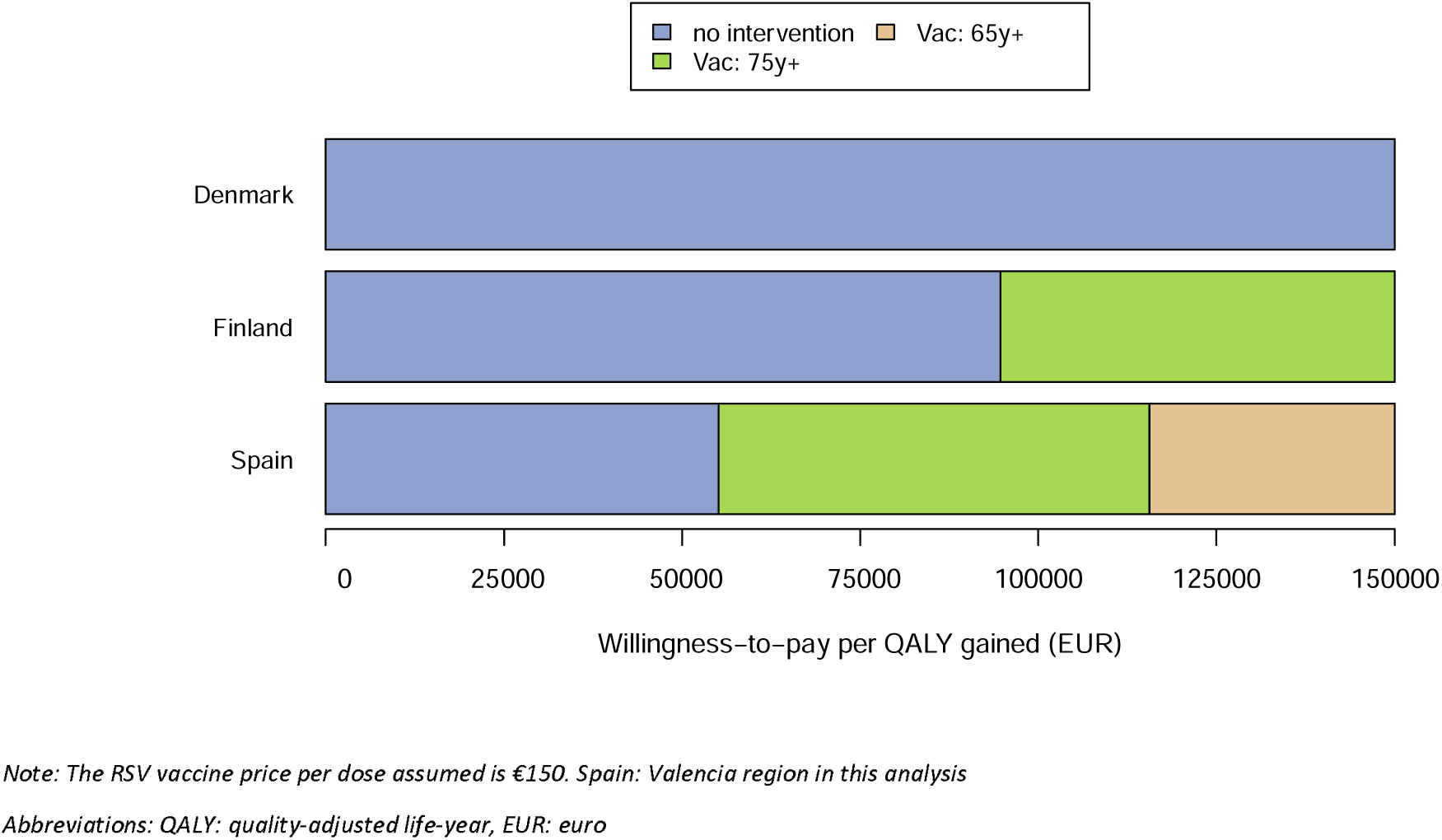
Using adjusted RSV-confirmed disease burden: preferred strategy based on cost-effectiveness analysis from the healthcare payers’ perspective.

**Figure 4:**
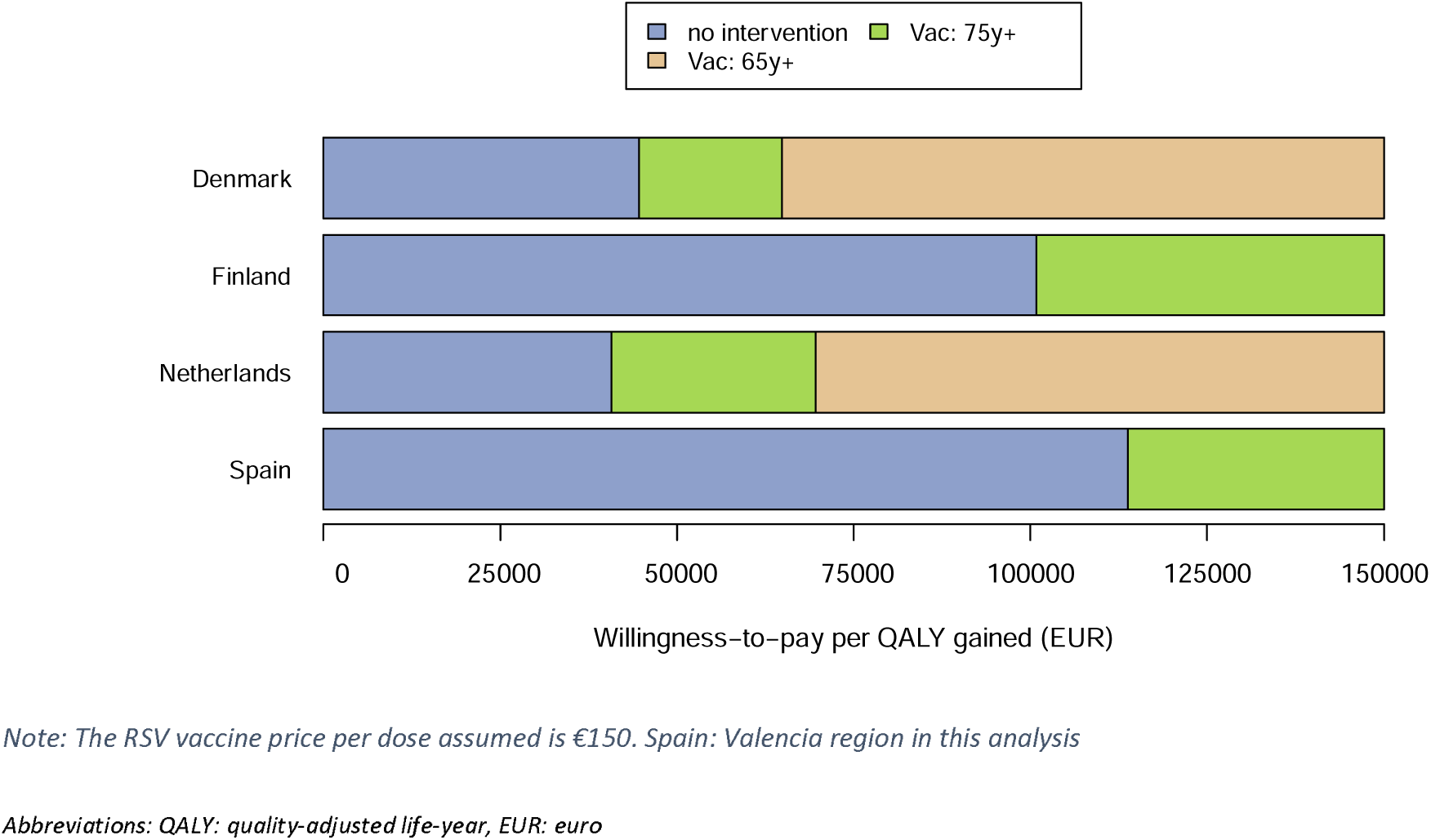
Using time-series-modelling RSV estimates: preferred strategy based on cost-effectiveness analysis from the healthcare payers’ perspective.

In all four countries and using all hospitalisation data, the 60y+ strategy was not preferred up to a WTP value of €150 000.

### Influence of price

Variations in the vaccine price from €50 to €250 per dose changed the preferred strategy over a wide range of WTP levels from the HCP perspective. (1) Using the adjusted RSV-ICD-coded dataset (Supplement Figure 17), ‘no intervention’ was cost-effective in Denmark irrespective of WTP per QALY. However, at €50 per dose, in Finland the 75y+ strategy was cost-effective at a WTP value of €40 000, whereas in the Netherlands the 75y+ strategy could be cost-effective at a WTP value of €80 000. (2) Using the adjusted RSV-confirmed dataset (Supplement Figure 18), findings are consistent in Denmark and comparable in Finland versus adjusted ICD-coded hospitalisation. In Spain-Valencia, at €50 per dose, the 75y+ strategy would be cost-effective at a WTP value of €20 000 per QALY gained. (3) Using TSM estimates (Figure 5) and a price of €50 per dose, the 75y+ strategy would be cost-effective at a WTP of €20 000 per QALY gained in Denmark, €30 000 in Finland, €10 000 in the Netherlands, and €50 000 in Spain-Valencia. The 65y+ strategy would become cost-effective at higher WTP ranges per QALY gained: €30 000 in Denmark and the Netherlands, €80 000 in Finland, and €100 000 in Spain-Valencia. The 60y+ strategy might become cost-effective in Denmark and the Netherlands at WTP values of €100 000 and €120 000 per QALY gained, respectively. The price reductions led to sharp decreases in the WTP level at which strategies became cost-effective for Denmark and the Netherlands. This effect was less pronounced for Finland and Spain-Valencia. Overall, decision uncertainty was highest nearby the WTP values at which the cost-effective strategy switched.

**Figure 5:**
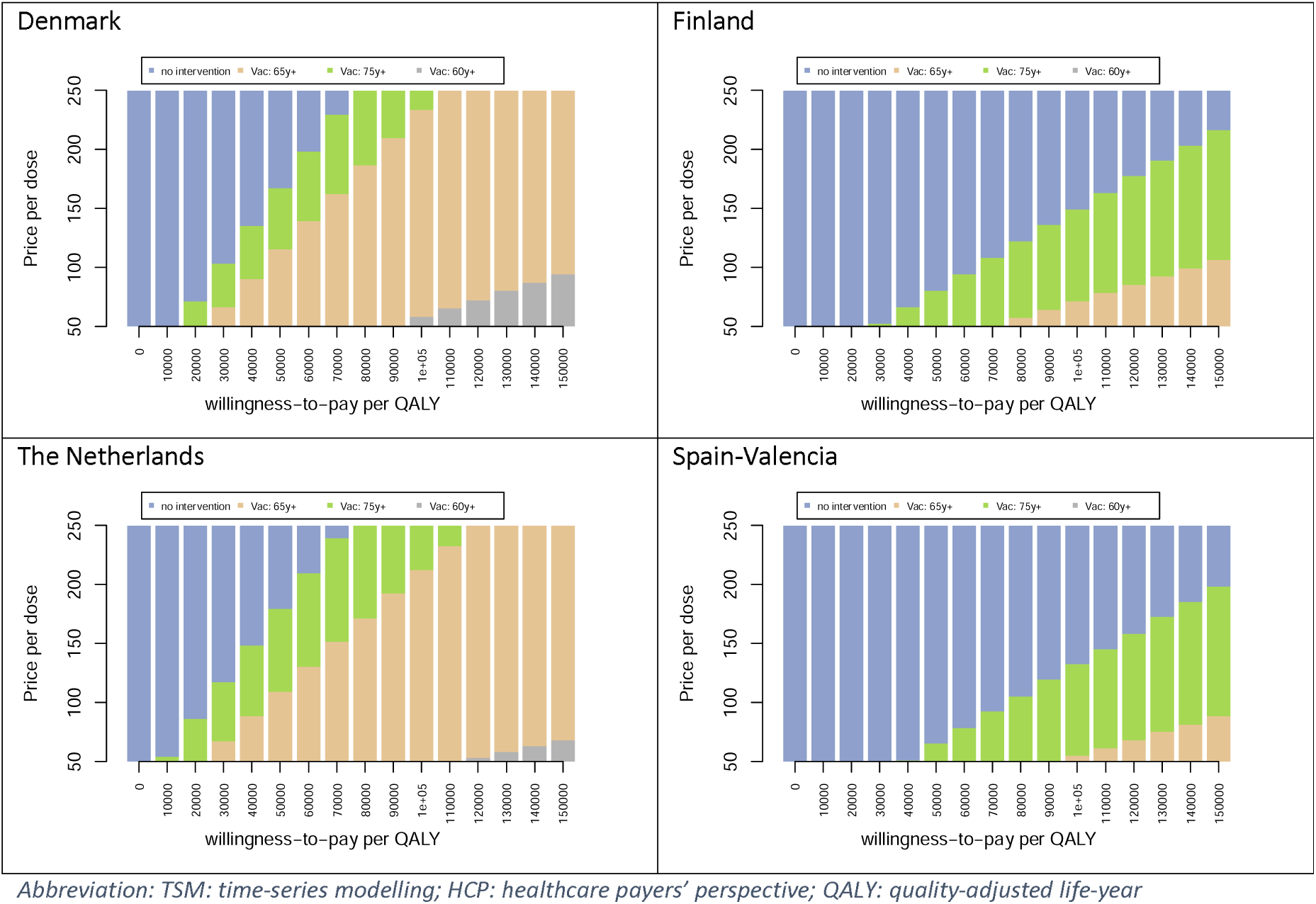
Using TSM estimates: Price threshold analysis showing the cost-effective strategy at different WTP thresholds from HCP’s perspective.

### Highly influential factors on the optimal strategy (other than price)

#### Choice of datasets

The choice of optimal strategy at a given WTP level and price depended in all countries heavily on the choice of dataset to estimate the incidence of RSV hospitalisations (RSV-ICD-coded, RSV-confirmed and RSV-attributable).

#### Hospital case fatality ratio (non-age specific and age-specific)

The hCFR was identified as another key driver. The EVPPI graphs demonstrated that the uncertainty around the non-age-specific hCFR caused the most decision uncertainty for all countries across all three datasets (Figure 6 and Supplement Figures 10, 13 and 16).

**Figure 6:**
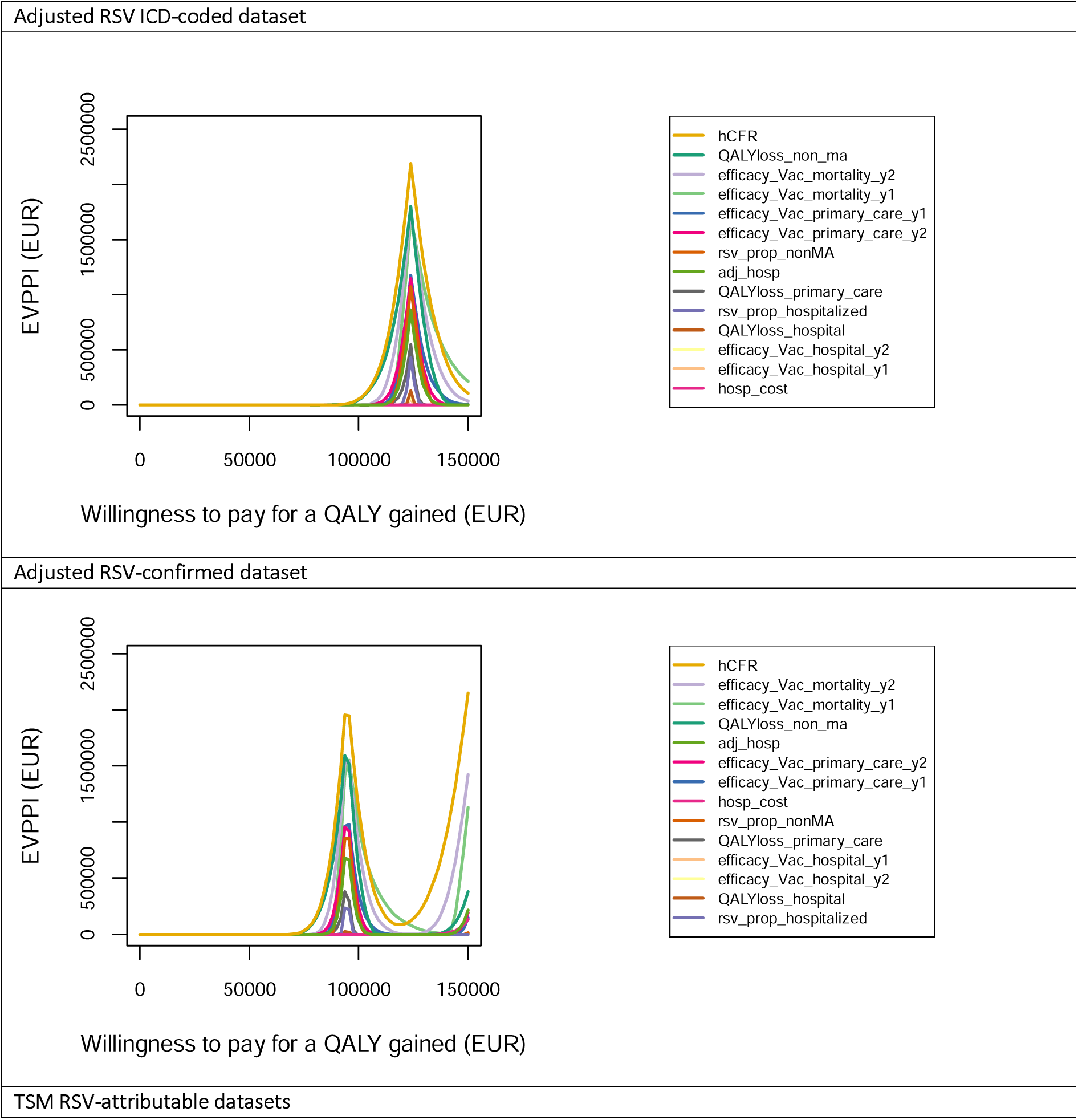
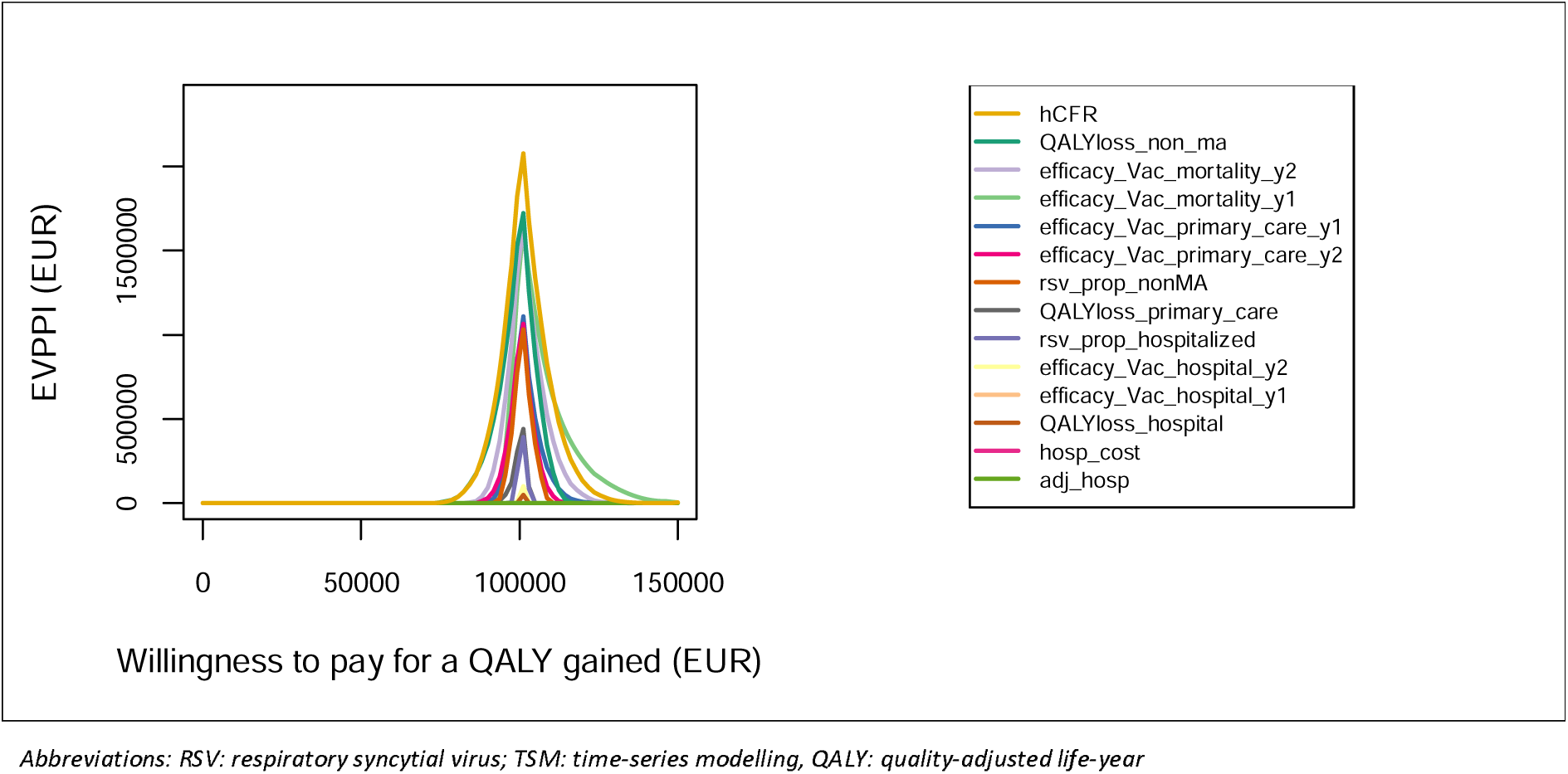
Expected value of partial perfect information for Finland, using three different hospitalisation datasets.

Compared with assuming non-age-specific hCFR for adults 60y+, using relatively higher hCFRs among adults 75y+ made the 75y+ strategy preferred at lower WTP values, and lower hCFRs in the 60-64y and 65-74y age groups made the 65y+ strategy only preferable over the 75y+ strategy at much higher WTP values (Supplement Figure 20).

#### RSV vaccine efficacy values against mortality

As illustrated by the EVPPI graphs, the uncertainty around first-and second-year RSV vaccine efficacy values against mortality were the top-ranking influential drivers (Figure 6, Supplement Figures 10, 13 and 16), because ca. 80% of the QALY gains came from the RSV-related deaths averted across all four countries.

#### QALY losses due to non-medically attended cases

The large number of non-MA cases make the estimate of the number of non-MA episodes per primary care visit over its relatively wide uncertainty interval (2.27 [uniform distribution: 1.14-3.41]) influential for the costs per QALY gained estimates (Figure 6, Supplement Figures 10, 13 and 16).

#### Adjustment factor for diagnostic under-ascertainment

The adjustment factor of 2.2 for diagnostic under-ascertainment of hospitalised cases was also influential when using the RSV-ICD-coded and RSV-confirmed datasets (Supplement Figures 20). Its parametric uncertainty was influential when using the adjusted RSV-ICD-coded and adjusted RSV-confirmed datasets (Figure 6 and Supplement Figures 10 and 13).

### Other influential factors on the optimal strategy

The duration of protective efficacy showed more impact than the type of waning curve assumed (Supplement Figure 19). Scenarios considering a seasonal shift of RSV as observed during the COVID-19 pandemic, administering RSV vaccine prior to a “mild” season and applying lower vaccine efficacy in adults 80y+ all yielded negative impacts on the cost-effectiveness results compared to the reference analysis. Scenarios administering RSV vaccine prior to a “severe” season, using higher QALY losses associated with RSV illness and a higher ratio of hospital to primary care, led to more expansive cost-effective strategies (Supplement Figure 15). Nevertheless, RSV vaccine coverage increases in the 60-64y age group had limited impact (Supplement Figure 15), in the context of comparing multiple age-based RSV strategies using a single RSV vaccine at price of €150 per dose.

## Discussion

We performed a full incremental analysis comparing no intervention and three vaccination strategies in adults aged 60, 65 and 75 years and above to each other using a hypothetical RSV vaccine which bridged phase 3 trial efficacy data available up to June 2024. Our analyses used three hospitalisation datasets in four countries to explore the influential drivers of the cost-effectiveness of RSV vaccination in European older adults. Although there are substantial differences observed among countries in term of cost-effectiveness, the influential drivers are consistent across countries. We found that apart from the vaccine price, the key driver of decision uncertainty regarding which strategy is cost-effective in all countries is the choice of the RSV hospital admissions dataset to use. The top-ranking uncertainty drivers are the non-age-specific hCFR, the vaccine efficacy values against mortality and the adjustment for diagnostic under-ascertainment of hospitalised cases, when using the adjusted RSV-ICD-coded and RSV-confirmed datasets. The uncertainty around the ratio of non-MA cases per primary care visit was also particularly influential.

Since RSV clinical symptoms are indistinguishable from those of many other respiratory virus infections and laboratory testing for RSV is relatively rare in older European adults, RSV-ICD-coded hospitalisation rates can substantially underestimate the disease burden, especially in countries without routine RSV testing [26, 46]. We, therefore, investigated the impact of using RSV-ICD-coded (except Spain-Valencia), RSV-confirmed (except the Netherlands), or TSM model-based RSV-attributable hospitalisations to inform RSV hospitalisation rates. We showed this impact to be very high. For instance, after applying an adjustment factor of 2.2 to the RSV-ICD-coded hospitalisations and RSV-confirmed, no intervention was cost-effective up to the WTP value of €150 000 per QALY gained in Denmark and the Netherlands, and up to €124 000 per QALY gained in Finland. No intervention was cost-effective up to a WTP value of €55 000, in Spain-Valencia when using adjusted RSV-confirmed hospitalisations. However, model-based analyses of TSM data yielded increased age-specific RSV attributability, and when using the resulting hospitalisation rates, the 75y+ strategy was shown to be cost-effective at much lower WTP values per QALY gained in Denmark and the Netherlands.

The TSM-based approach used in this analysis is a method commonly used to estimate the burden of many other diseases such as influenza [47]. In our analysis, we used laboratory test data series on four pathogens (influenza A, influenza B, SARS-CoV-2 and RSV) that cause RTIs (in Spain-Valencia, ILI was used as a proxy for RTI). Although these are four highly important pathogens for RTIs, this is not an exhaustive inclusion of potential causative pathogens and, therefore, the attributable fractions estimated for each or some of these pathogens to RTI hospitalisation may be overestimations, although this was partially adjusted for by including a constant term. On the other hand, RSV-ICD-coded hospital admission data are likely to substantially underestimate RSV-related hospital admissions [46, 48]. RSV diagnostic tests were also not systematically performed except in Spain-Valencia.

When adjusting RSV-ICD-coded and RSV-confirmed datasets with the diagnostic under-ascertainment factor of 2.2, the findings were substantially different than those using TSM estimates in Denmark. The large differences were also observed in the Netherlands when comparing the adjusted RSV-ICD-coded hospitalisation with the TSM estimates. However, the results of using three datasets were comparable in Finland. In Spain-Valencia, the adjusted-RSV-confirmed dataset showed more favourable results for RSV vaccination. This could be explained by the fact that testing was already systematic in all included in patients in Spain-Valencia. However, the use of ILI as inclusion criteria (in Spain-Valencia) is known to underestimate the real RSV burden in the surveillance networks compared to other case definitions an under-ascertainment that has not been corrected for here [26, 49, 50]. Hence, the adjustment factor and its uncertainty need to be interpreted with caution, taking into account country-specific coding and testing practices.

Moreover, the hCFR and the specification of its age dependency were influential (Supplement Figures 10, 13, 15 and 20). Despite the efforts to link the RSV-confirmed deaths with hospital and mortality registries of multiple countries, we were only able to use the age-specific hCFR for Finland, which had a sufficient sample size and was compliant with the applicable data privacy law. Importantly, we demonstrated that acknowledging the age-dependent nature of the hCFR is important when selecting optimal age-based RSV vaccination strategies in older adults.

Our price threshold analysis showed that different “vaccine price per dose” thresholds exist for different “WTP per QALY” levels, implying different optimal choices of strategies. When considering vaccination strategies among 60y+, 65y+ and 75y+, the strategy protecting the oldest (75y+) age group is first selected from the HCP perspective. With increasing WTP values or decreases in price, the 65y+ and 60y+ strategies might become the optimal strategy. Embedded in our calculations is the budget impact. Clearly, the smaller older-age cohorts have a budgetary advantage over larger younger-age ones. In many countries, budget-impact considerations may determine the choice of preferred strategy as much as cost-effectiveness considerations do. For instance, Hodgson and colleagues assumed one year protection and estimated the threshold price at which the 75y+ strategy can be cost-effective at a much higher price (£20.71) than the price (£3.63) at which it would be considered affordable, given a UK-set budget constraint for an individual programme of £20 million annually during the first three years of implementation [51].

By exploring many different waning efficacy profiles, we found that duration of protection is more influential than the type of waning curve to achieve a cost-effective result. However, this is for a single vaccine analysis and is likely to become more nuanced in head-to-head comparisons of different vaccine brands, especially so when vaccines are marketed to compete more on marginal differences in age-specific protective efficacy over time than on more influential differences in price-setting. Given the trial-based information available, with different case definitions of the clinical endpoints of the licensed vaccines, such a head-to-head comparison is beyond the scope of this paper.

We assumed the RSV vaccines would be administered prior to the expected RSV season in Europe in October (with instant protection) in order to maximise the effectiveness. However, depending on the country, influenza, pneumococcal and COVID-19 vaccines are also typically administered in the last quarter of the calendar year in the same target populations, which can lead to implementation challenges for RSV vaccines due to crowded schedules. Given that RSV vaccines protect longer than one season, a year-round programme might be considered, possibly together with the pneumococcal or herpes zoster vaccine among older adults. However, given the clear seasonal pattern of RSV in Europe, a year-round programme would be less effective than a seasonal programme due to waning protection. Moreover, the coverage of pneumococcal vaccines is generally lower than the coverage of influenza vaccines among adults 65 years and above in Europe. Joint advice for RSV, influenza and COVID-19 vaccine before the winter might simplify the communication and, hence, achieve higher coverage.

This is the first analysis that assessed the cost-effectiveness of three age-based RSV vaccination strategies among older adults in more than two countries and identified the key evidence gaps that were most influential to cost-effectiveness. Previous analyses on a single country or two countries did not explore the sensitivity to different data sources, the influence of a shift in seasonality and the severity of a season [30, 31, 41, 51, 52]. Our results are comparable with two cost-effectiveness analyses published in 2023, which applied vaccine protection over two RSV seasons. Moghadas and colleagues evaluated both marketed RSV vaccines in the US among adults aged 60y+ from a societal perspective. They concluded that both vaccines can be cost-effective at a price of $235 (∼€219) and $245 (∼€228) per dose given a WTP value of $95 000 (∼€88 000) per QALY gained, with sensitivity analyses on WTP values of $80 000 (∼€75 000) and $120 000 (∼110 000) per QALY gained [30]. Using the time-series estimates, we find that from the HCP’s perspective and at an RSV vaccine price of €200 per dose, the 65y+ strategy would be cost-effective only at high WTP values of €90 000 and €100 000 per QALY gained in Denmark and the Netherlands, respectively. We used age-specific hospitalisation rates, which led to much higher estimates for the 85y+ age group (∼366 per 100 000 in the Netherlands and 533 per 100 000 in Denmark) in comparison to Moghadas et al (214 per 100 000 for any adult aged 60y+, with a 54.8% proportion in 85y+) [30]. However, Moghadas et al included both market and non-market productivity losses due to RSV premature deaths based on US guidelines. Using a cost-utility framework as prescribed by the pharmacoeconomic guidelines for the countries in our study, we did not include such monetised productivity costs. Wang and colleagues conducted a cost-effectiveness analysis in Hong Kong and used adjusted hospitalisation rates ranging from 23.0 to 221.4 per 100 000 in the 60-64y and 75y+ age groups, respectively [31]. By adjusting ICD-coded hospitalisations, our analysis estimated a lower rate in Denmark (4.2 and 17.8 per 100 000), the Netherlands (0.4 and 2.2 per 100 000) and Spain (2.6 and 13.1 per 100 000), but a higher rate in Finland (37.1 and 168.0 per 100 000). The Hong Kong analysis also generally highlighted the price and disease burden as influential drivers for the cost-effectiveness analysis, as did previous analyses, which assumed protection over one RSV season [41, 51, 52].

Unlike in the UK, where the WTP thresholds of £20 000 (∼€24 000) and £30 000 (∼€35 000) per QALY gained from the National Health Service (NHS) perspective are clearly indicated, the four European countries in our analysis do not have an explicit WTP reference value [53]. In Denmark, a WTP value of DKK 88000 (∼€12 000) per QALY gained was estimated via a population survey in 2001, which might need updating [54]. In Finland, infant varicella, pneumococcal and rotavirus vaccination programmes were considered to be cost-effective at WTP values ranging from €15 000 to €25 000 per QALY gained from HCP perspective [55]. In the Netherlands, the reference values of €20 000, €50 000 and €80 000 per QALY gained depended on the disease severity and perspective [56]. In Spain, a threshold of €30 000 from the NHS perspective is commonly cited [57]. Instead of using one or multiple arbitrary threshold values, our analysis presented findings over a range of WTP values (€0-150 000 per QALY gained) to assess the influence of the WTP value on the optimal choice.

Our study has some additional strengths. It used the most recent country-specific data to populate the model according to country-specific pharmacoeconomic guidelines. Considerable differences in the RSV health and economic burden exist between countries, and policy makers should reflect on these when making decisions. Most strikingly, RSV seasonality changed during the COVID-19 pandemic in many countries. Therefore, we simulated an off-season hospital admission peak (four months earlier) observed during the pandemic and found the results would be less favourable because administering RSV vaccine in October would miss either a portion or the entirety of the seasonal peak. Furthermore, we also investigated RSV vaccine introduction in a country with a biannual seasonal pattern and found that is considerably more beneficial to introduce the vaccine first prior to a predicted “severe” season than a “mild” season. Furthermore, we explored multiple vaccine efficacy waning scenarios in combination with duration of protection based on the latest clinical data and demonstrated that the duration of protection have more impact than the type of waning curve assumed. Finally, extensive sensitivity analyses were performed using a wide range of WTP values to investigate the sensitivity of the optimal choice of strategy to the adopted WTP level.

An important limitation of our analysis is that we investigated age-based and not risk group-based strategies. We investigated one-time RSV vaccination, but did not attempt to model scenarios with repetitive vaccination of cohorts over time. Future refined information on vaccine effectiveness and durability may allow the evaluation of such strategies realistically in future. Moreover, hCFR was used to estimate RSV-related deaths in the inpatient setting. However, deaths that occurred in community setting (i.e., nursing home) were not captured due to data availability, hence, it might underestimate cost-effectiveness of the RSV vaccine.

There are a few other limitations. Due to data limitations, the hospitalisation rate of a wide age group (18-64 years) was applied to a small subgroup (60-64 years) in all four countries when using TSM estimates. However, the impact on the overall insights from this analysis was negligible. We were unable to explicitly evaluate the impact of potentially disproportionately prevented re-admissions, long-term consequences due to RSV hospitalisation (e.g., transfer to a nursing home instead of home after discharge), RSV vaccine’s potential prevention of exacerbations of chronic respiratory disease and cardiac events [58], and productivity losses of caregivers under the societal perspective, mainly because the occurrence of, and effectiveness against, these endpoints were unavailable.

## Conclusion

Large data gaps and uncertainties around the RSV burden among older adults persist in many European countries, contributing to substantial uncertainties regarding the cost-effectiveness of intervention programs. Given that these countries need to make urgent decisions about RSV vaccination, more refined age-, risk group-, and country-specific RSV burden data are crucial. Evidence-based vaccine introduction decisions will require greater investment in enhanced RSV surveillance and better data linkage systems to enable accurate assessments of the age-and country-specific RSV burden, especially in tertiary and primary care settings, such as those established in Finland. Without this, costly and potentially suboptimal decisions risk being made, leading to substantial opportunity costs due to the large target populations to be vaccinated.

## Data Availability

Almost all the data analysed and generated during this study are included in this published article and its supplementary files. Formal requests for additional data can be made to the corresponding author (XL) or the senior author (PB).

## List of abbreviation

ACIP: Advisory Committee on Immunization Practices
CI: confidence interval
COVID-19: coronavirus disease 2019
CPI: consumer price index
EVPPI: expected value of partial perfect information
hCFR: in-hospital case fatality ratio
HCP: healthcare payer
ICD: International Classification of Diseases
ILI: influenza-like-illness
JCVI: Joint Committee on Vaccination and Immunisation
MA: medical attendance
MCMARCELA: Multi-Country Model Application for RSV Cost-Effectiveness policy for Adults
PCR: polymerase chain reaction
PROMISE: Preparing for RSV Immunisation and Surveillance in Europe
QALY: quality-adjusted life-years
RSV: respiratory syncytial virus
SARS-CoV-2: severe acute respiratory syndrome coronavirus 2
TSM: time-series modelling
UK: United Kingdom
US: United States
WTP: willingness-to-pay
Y: year

## Declarations

Ethics approval and consent to participate

Not applicable

## Consent for publication

Not applicable

## Funding

This project has received funding from the Innovative Medicines Initiative 2 Joint Undertaking (JU) under grant agreement number 101034339. The JU receives support from the European Union’s Horizon 2020 research and innovation program and European Federation of Pharmaceutical Industries Associations. MJ received funding from the NIHR Health Protection Research Units in Modelling and Health Economics (NIHR200908) and Immunisation (NIHR200929).

The funders of the study had no role in study design, data collection, data analysis, data interpretation, or writing of the report.

## Competing interests

Outside the submitted work, PB declares funding received by his institute from Merck for research on varicella-zoster and Pfizer for research on pneumococcus vaccine, XL declares funding received by her institute from Icosavax, but they have not received any personal fees or other personal benefits. CKJ reports a research grant from Nordsjællands Hospital, travel grants from the University of Copenhagen, William Demants Fond in Denmark, and the European Society of Clinical Virology and expert consultation fees from Sanofi, outside of the submitted work. Outside the submitted work, HN declares consulting fees received from WHO, Pfizer, Bill and Medlinda Bill and Melinda Gates Foundation and Sanofi, honoraria for lectures/presentations from Astra Zeneca, GSK and Pfizer paid to his institution, travel grants from Pfizer and Sanofi, and participation on a Data Safety Monitoring Board or Advisory Board of GSK, Sanofi, Merck, Icosavax, Pfizer (paid to his institution), as well as WHO and ResViNET (unpaid). AUF declares grants/contracts from Moderna and VAC4EU paid to her institution, outside submitted work. AOS and JDD declares grants/contracts from Moderna, MSD and GSK paid to their institution, outside submitted work. AOS also declares consultancy fees from GSK and Moderna, outside submitted work. JDD declares consultancy fees and honoraria from GSK, Pfizer and MSD, and travel grants from Sanofi and GSK, outside the submitted work. HS declares attending Adult Immunization Board meetings twice a year, the meetings (and travel) are funded by an unrestricted grant from Vaccines Europe, outside submitted work. LW declares grants received from Research Foundation Flanders (FWO), Antwerp University Research Council and Horizon Europe 2.1 – Health paid to his institute, outside the submitted work. MJ declares research grants from NIHR, RCUK, BMGF, WHO, Gavi, Wellcome Trust, European Commission, InnoHK, TFGH, CDC, Gates Foundation paid to his institution, outside the submitted work.

Other authors declared no competing interests.

## Authors’ contributions

PB and HN initiated and supervised the study. XL, LW, JB and PB conceptualised the study. XL analysed input data, performed literature searches and auxiliary data collection. XL, LW and JB wrote the cost-effectiveness model codes. XL performed the cost-effectiveness analyses. CKJ, AUF, and LT conducted national/regional database analyses, and collected and validated data in Denmark, Spain-Valencia, and Finland, respectively. CKJ, ROY, and TL led and estimated the time-series model-based hospitalisations. HS, AOS and JDD provided implementation, resource use and cost data for Finland and Spain. HN, MJ, JB and PB advised on model parameters, intervention characteristics and scenario analyses. XL, LW, JB and PB wrote the initial manuscript draft. All authors critically reviewed the manuscript and provided final approval of the manuscript.

## Acknowledgements

PROMISE investigators: Harish Nair and Harry Campbell (University of Edinburgh, Edinburgh, UK); Louis Bont (University Medical Centre, Utrecht, the Netherlands); Philippe Beutels (University of Antwerp, Antwerp, Belgium), Peter Openshaw (Imperial College, London, UK), Andrew Pollard (University of Oxford, Oxford, UK), Hanna Nohynek (Finnish Institute for Health and Welfare, Helsinki, Finland), John Paget (Netherlands Institute for Health Services Research, Utrecht, Netherlands), Eva Molero (Teamit Research S.L., Barcelona, Spain), Javier Díez-Domingo (Vaccine Research Department, FISABIO-Public Health and CIBERESP, ISCIII, Valencia, Spain), Rolf Kramer (Sanofi Pasteur, Lyon, France), Jim Janimak (GlaxoSmithKline, Wavre, Belgium), Veena Kumar (Novavax, Gaithersburg, Maryland, United States), Elizabeth Begier (Pfizer Limited, New York City, New York, United States), Jenny Hendri (Janssen, Beerse, Belgium).

The authors would like to thank You Li, Valerie Sankatsing and Matthieu Beuvelet for their critical review of our study report and valuable comments.

## Notes

### Summary of Updates

The first author identified a coding error in the previous preprint. As a result, a second modeller conducted a thorough, independent review of the entire code. We have since updated the preprint accordingly. The key message that the source of hospitalization is the most influential driver remains unchanged. We revised Figure 1-6 Supplement file: section 2

