## Supplement for "Influential drivers of the cost-effectiveness of respiratory syncytial virus vaccination in European older adults: A multi-country analysis"

Affiliation:

Xiao Li

Address: Centre for Health Economics Research & Modelling Infectious Diseases,

Vaccine & Infectious Disease Institute, Campus Drie Eiken (D.R.212), Universiteitsplein 1

University of Antwerp, Belgium

### 1 SUPPLEMENT METHODS

Our cost-effectiveness analyses of respiratory syncytial virus (RSV) vaccination in older adults were performed in four European countries, namely, Denmark, Finland, the Netherlands and Spain-Valencia region.

#### 1.1 Input parameters

Country-specific input parameters were collected accounting for healthcare system characteristics. The primary sources of input parameters in this analysis included: national patient registries [1], active surveillance data [1], the time-series modelling (TSM) [2], national tariffs and literature. S. Table 6 summarises common and country-specific input parameters.

##### 1.1.1 RSV-related hospital admissions

We explored the impact on the cost-effectiveness results in scenario analyses by using the following sets of assumptions to estimate hospital admissions: (1) RSV- International Classification of Diseases (ICD)-coded hospitalisations with and without an adjustment factor (except Spain-Valencia), (2) RSV-confirmed hospitalisations (except the Netherlands) with and without an adjustment factor, and (3) RSV-attributable hospitalisations (see details below).

All weekly data were converted to calendar months using the Lubridate R package [3]. The monthly age-specific number of RSV hospital admissions was obtained by averaging over three or four seasons prior to the coronavirus disease 2019 (COVID-19) pandemic (2016/2017 to 2019/2020 seasons). The age-specific RSV hospitalisation rates were calculated based on average population size (or catchment population for Spain-Valencia) over the same period. Lastly, the latest (2023) population size estimates were used to estimate the number of RSV hospital admissions per age group in our forward model projections.

###### 1.1.1.1 RSV-ICD-coded hospital admissions

The RSV-ICD-coded hospital admission data are often used to estimate the in-hospital burden of RSV infections. Based on retrospective national hospital registries analyses, the RSV-ICD-coded data are presented by calendar weeks in Finland and the Netherlands (S. Figure 1). The Danish RSV-ICD-coded estimates are presented in calendar months (S. Figure 2), in line with the European General Data Protection Regulation (GDPR), because of small weekly numbers of admissions raising the possibility of identification. In Valencia, a region of Spain (Spain-Valencia), the RSV hospitalisation data were collected prospectively from the Valencia Hospital Surveillance Network. However, the sample size of RSV-ICD-coded admissions was insufficient, and the active surveillance was designed to capture the pathogen-specific hospital admissions, hence we exclude Spain-Valencia in the ICD-coded analysis (more details in the section below). The list of ICD-codes used in the analyses were published in previous studies [4, 5].

In Denmark, Finland and the Netherlands, RSV-ICD-coded data were available for the age groups 65-74 years, 75-84 years and 85 years and above. For the age group below 65 years, data were available for 50-64-year-olds in Finland, however, data were only available for the 18-64 years age group in Denmark and the Netherlands. For these two countries, we used the RSV-ICD-code-based analyses of the hospitalisation rate in 18-64-year olds for the age group 60-64 years, and this might, therefore, represent an underestimation for the RSV-related hospital admissions in these two countries for those parts of the analyses.

More generally, since many adults who present to healthcare services with respiratory symptoms are not routinely tested for RSV or coded as RSV in Europe, especially prior to the COVID-19 pandemic, historic RSV-ICD-coded hospital admission data are likely to substantially underestimate the "true" number of RSV-related hospital admissions [6, 7]. Moreover, multiple studies showed that diagnostic testing underestimates the overall RSV-related disease burden. A recent systematic review in high-income countries estimated a factor of 2.2 (2.07-2.36) to adjust for diagnostic under-ascertainment of admissions [8], hence, with and without this adjustment factor were included in our analysis.

*S. Figure 1: Country- and age-specific weekly RSV-ICD-coded number of hospital admissions in Finland and the Netherlands over time. Seasons are displayed from calendar week 27 (around early July) until week 26 of the following year.*

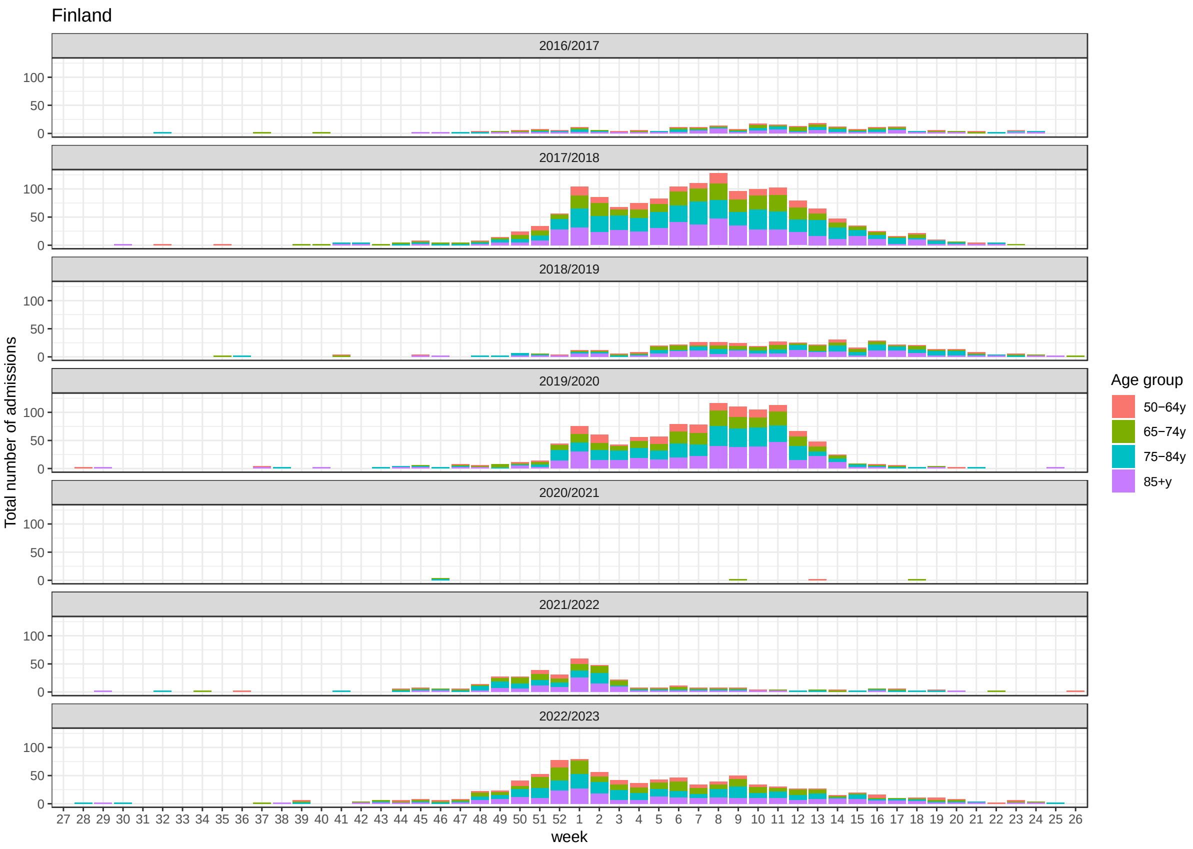

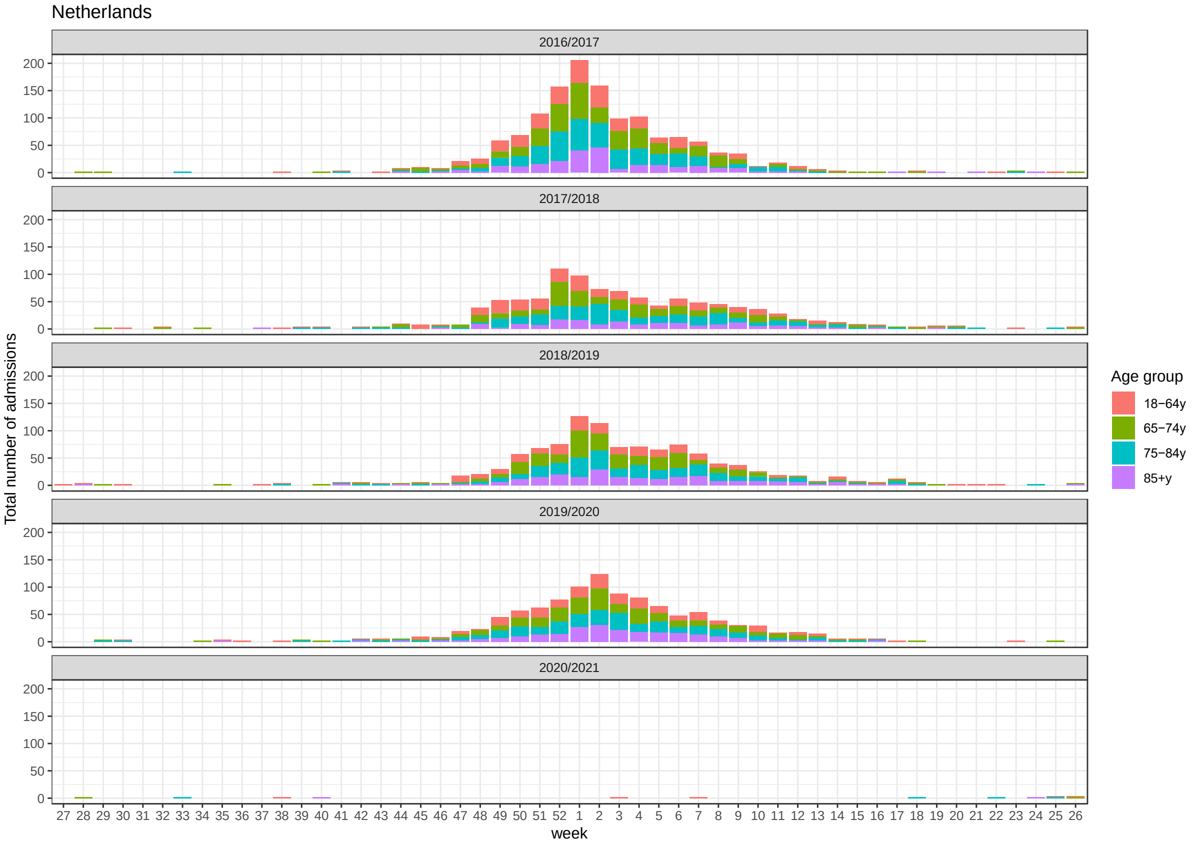
Netherlands
2016/2017
2017/2018
2018/2019
2019/2020
2020/2021
Total number of admissions
0
50
100
150
200
week
27 28 29 30 31 32 33 34 35 36 37 38 39 40 41 42 43 44 45 46 47 48 49 50 51 52 1 2 3 4 5 6 7 8 9 10 11 12 13 14 15 16 17 18 19 20 21 22 23 24 25 26
Age group
18−64y
65−74y
75−84y
85+y

*S. Figure 2: Age-specific monthly RSV-ICD-coded hospital admissions in Denmark per RSV season over time*

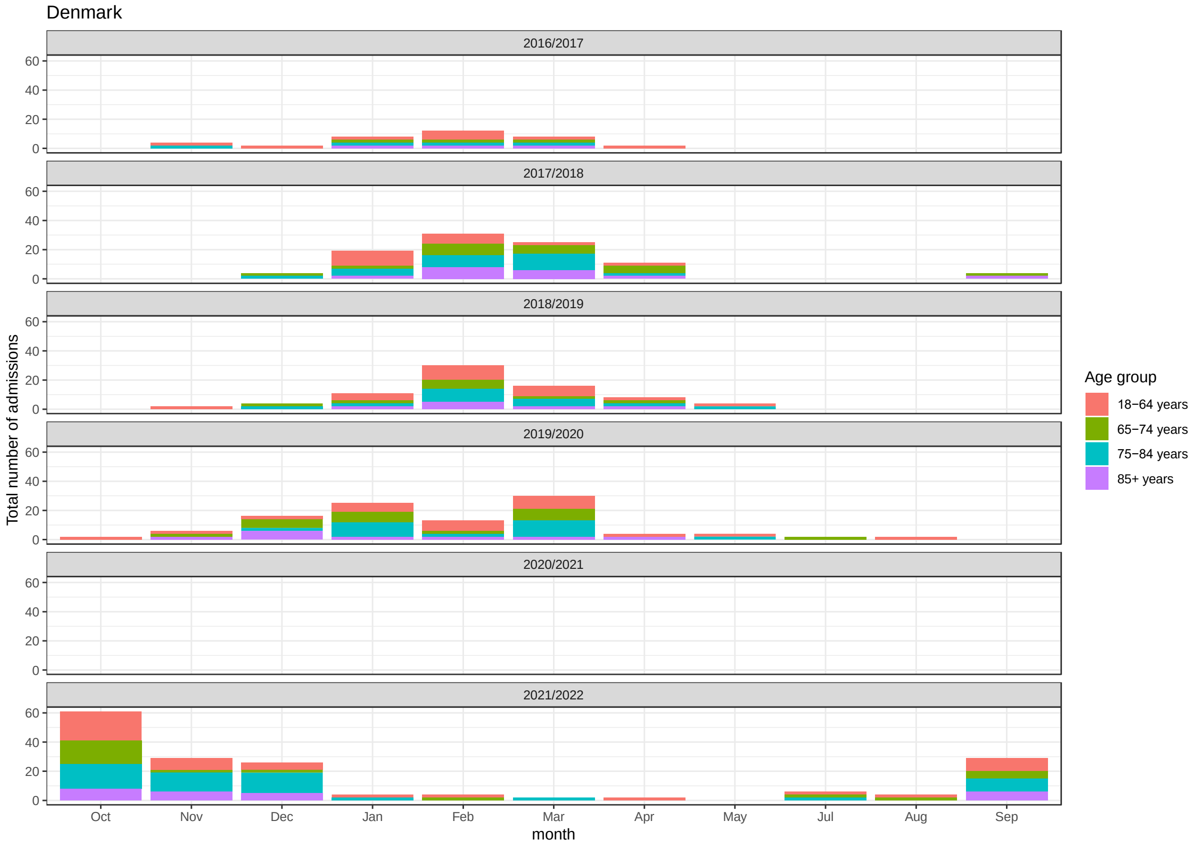

##### 1.1.1.2 RSV-confirmed admissions

RSV-confirmed hospital admission data were available for Denmark, Finland and Spain-Valencia. In Denmark, prior to the COVID-19 pandemic, only a proportion of adult patients with severe ARIs received laboratory tests, of which only approximately 50% were tested for RSV. In Finland, the majority of hospital-admitted patients with ARI had laboratory testing records. The details of the RSV-confirmed data are published elsewhere [1].

In Spain-Valencia, patient information was collected through the Valencia Hospital Surveillance Network for the Study of influenza and other Respiratory Viruses (VAHNSI), an active prospective hospital-based surveillance network. This constitutes active prospective surveillance with a catchment area that represents 21% of the overall population in Valencia

(~1 million, which in turn represents approximately 2% of the Spanish population). Patients had to fulfil the following criteria to be included in the study: being hospitalised via emergency room with a diagnosis compatible with an RTI, reside in the catchment area of one of the participating hospitals for at least 6 months, non-institutionalised, not discharged from a previous hospital admission in the last 30 days and give their (or their legally authorised representative) written consent. Patients ≥ 18 years old were included if, upon admission they met symptoms compatible with Influenza-Like-Illness (ILI) case definition, defined as the presence of at least one respiratory symptom (cough, sore throat or shortness of breath) with an onset within seven days prior to admission.

A nasopharyngeal and oropharyngeal swab was taken to all included ILI patients and tested for RSV, Influenza A and B via a multiplex-PCR. As VAHNSI is a surveillance network initially setup to cover influenza seasons, monitoring did not occur throughout the whole year and the duration of the monitoring was also different across seasons. Therefore, data was adjusted to the RSV circulation period in each of the surveillance years to allow data comparison across years with a different surveillance length. Circulation was defined as the weeks between the first of at least two consecutive weeks with two or more RSV cases and the week prior to the first of at least two consecutive weeks without RSV cases considering the PCR results of included patients from all ages. The loss of RSV confirmed cases after adjusting the data to the circulation period seasons is negligible. The duration in weeks of each surveillance year was calculated as the total number of epidemiological weeks in each RSV circulation period. The population was adjusted to the length of the RSV circulation period in years (approximated as 1 year; ~ 52.143 weeks). Due to the COVID-19 pandemic, no data was collected during season 2020/2021. During 2021/22 there were two RSV circulation periods (from W43 to W05 and from W13 to W26) and data from both circulations have been used in the study. It is worth noting that there were a few limitations when using data from this active surveillance: the use of ILI as inclusion criteria might underestimate RSV burden in active surveillances [9, 10]. The monitoring period tailored to the influenza season and this led to missing some RSV cases throughout the non-RSV season. More details are published elsewhere [1, 11].

Given RSV-confirmed data also exhibited similar issues as the RSV-ICD-coded data, we explore the impacts of using the RSV-confirmed data compared to using other hospital datasets.

*S. Figure 3: Country- and age-specific RSV-confirmed hospital admissions in Denmark (monthly), Finland (weekly) and Spain-Valencia region (weekly, active surveillance) over time.*

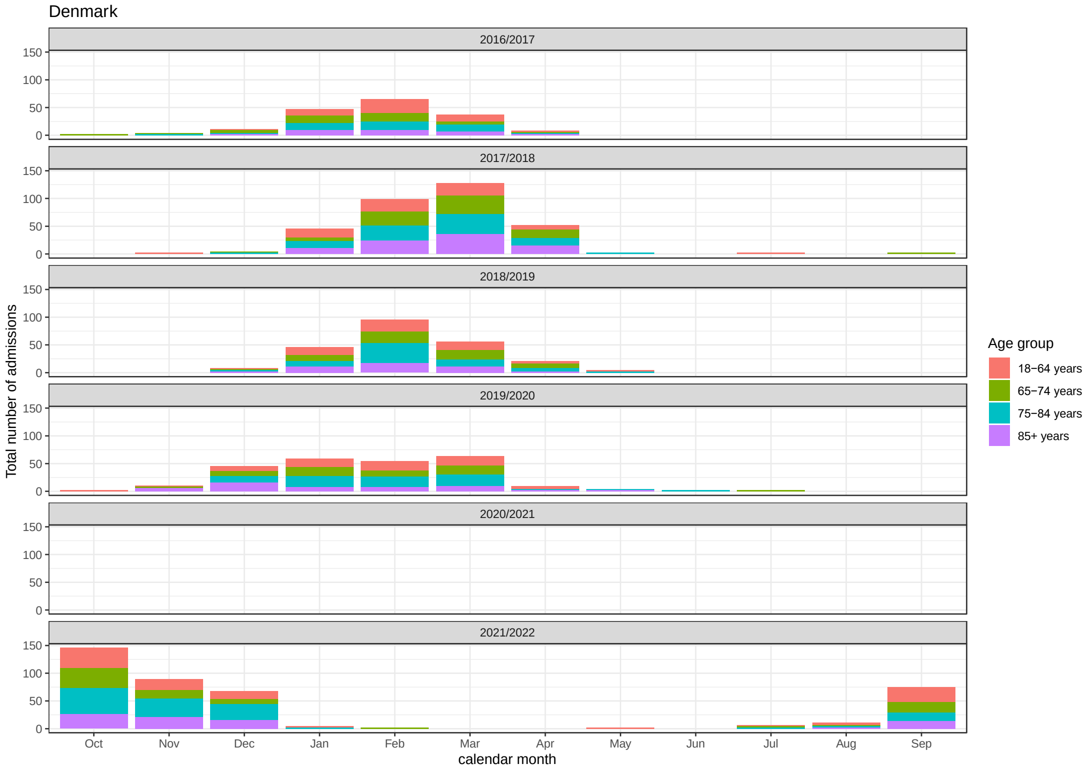

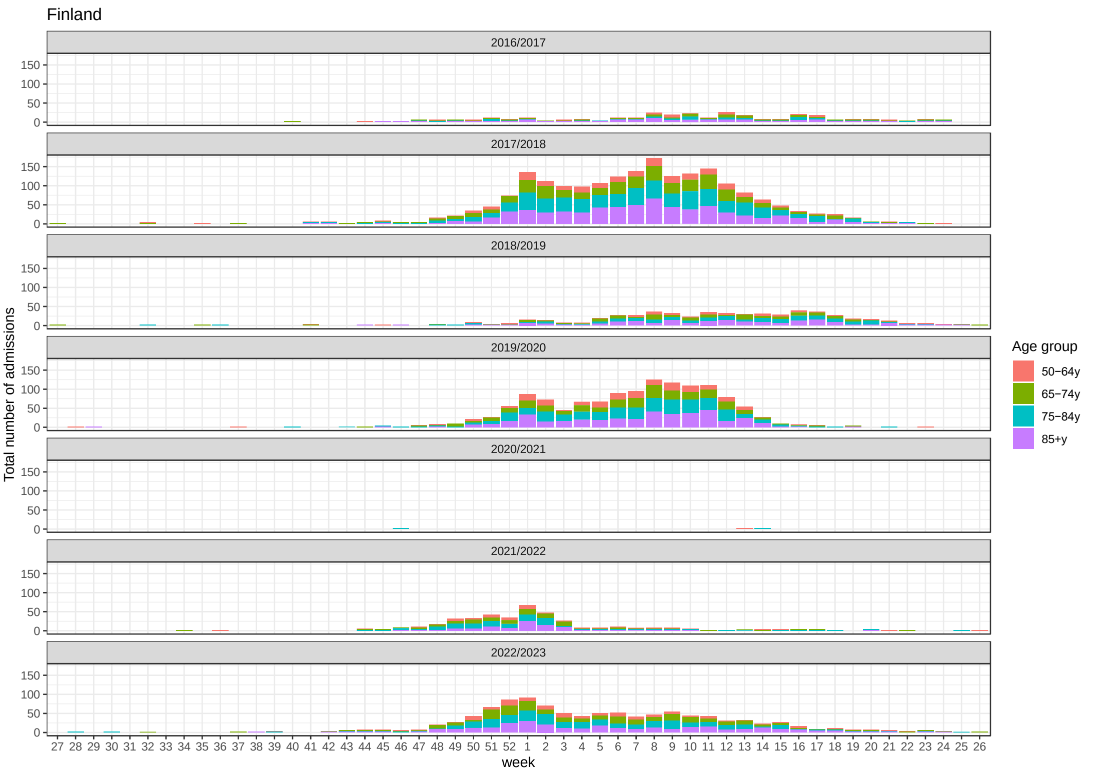

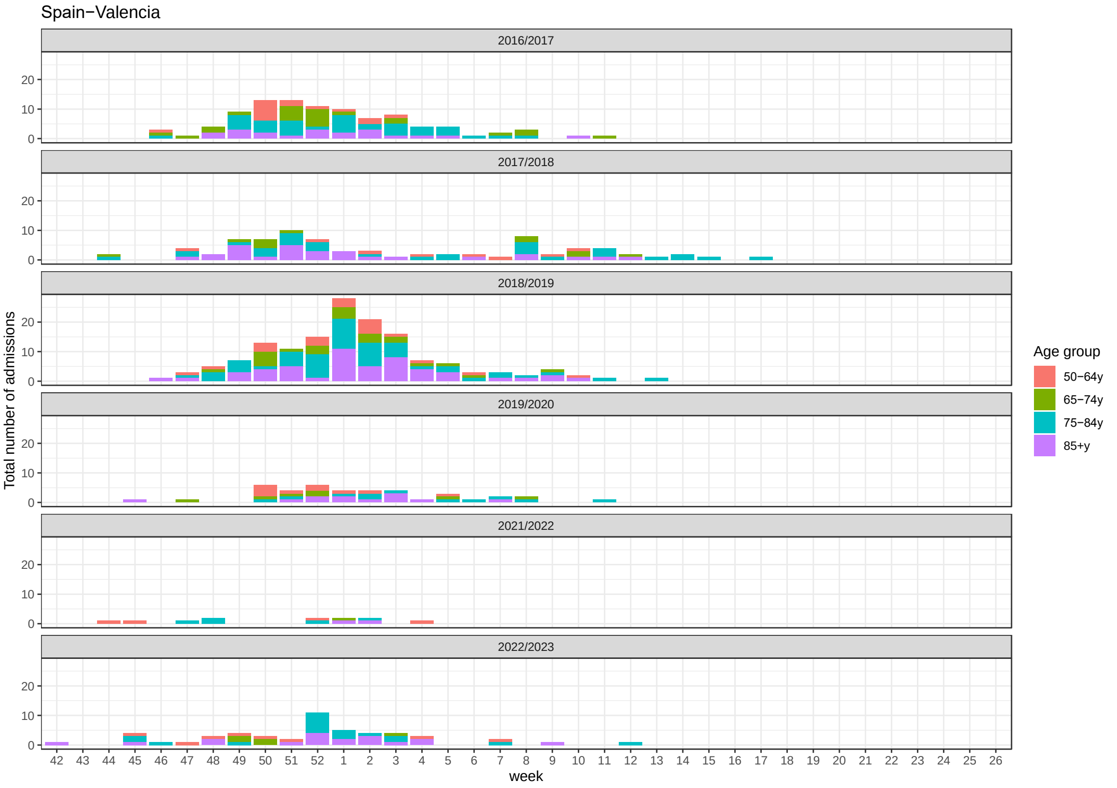

*Footnote: Denmark, data are displayed by calendar month from October to September the following year. Finland: data are displayed from calendar week 27 (around early July) until week 26 of the following year. Valencia-Spain: surveillance network initially setup to cover influenza seasons, monitoring did not occur throughout the whole year, but the duration of the influenza seasons.*

###### 1.1.1.3 RSV-attributable admissions

In addition, TSM was conducted to estimate weekly RSV-attributable hospitalisations by age group and over time. The method built on an analysis regressing influenza and RSV laboratory test frequency on respiratory tract infection (RTI) incidence over time (Johannesen et al. 2022) in Denmark, Finland and the Netherlands [12]. Briefly, RTI hospital admissions were defined as any non-scheduled/non-routine hospitalisation lasting more than 12 hours that included RTI-related ICD-10 codes, and considered a subgroup where any RSV-related ICD-10 code was present, with further subdivision into relevant age groups. Country-specific virological test data provided aggregated non-age-specific weekly numbers of positive tests, which were then used in a (Poisson) regression analysis with the age-specific number of weekly admissions serving as the outcome of interest. Thus, the number of RTI-related admissions in each country and age group was estimated by the main respiratory pathogen, distinguishing influenza A and B, RSV, and SARS-CoV-2. In Spain-Valencia, instead of RTI hospital admissions, ILI admission was used. As the active surveillance was not conducted throughout the whole year, the missing weeks were imputed for each age group using the seasonally decomposed missing value imputation from the imputTS R package in R. For further details on the TSM analysis, we refer you to the PROMISE deliverable report D1.7 [2].

Among older adults 65 years and above, the TSM estimates were substantially higher compared to the RSV-ICD-coded hospitalisations: 53-fold higher in Denmark, 2.6-fold higher in Finland, and 9-fold higher in the Netherlands. Since TSM estimates were unavailable for the 18-64-year age group in Finland, we used the RSV-confirmed data in this age group as an approximation. The TSM estimates were also higher than RSV-confirmed hospitalisations:19-fold higher in Denmark, 2.0-fold higher in Finland, and 1.3-fold higher in Spain-Valencia (after adjusted the catchment areas for active surveillance).

*S. Figure 4: Country- and age-specific weekly RSV-attributable hospital admissions in Denmark, Finland, and the Netherlands and Spain-Valencia over time. Seasons are displayed from calendar week 27 (around early July) until week 26 of the following year.*

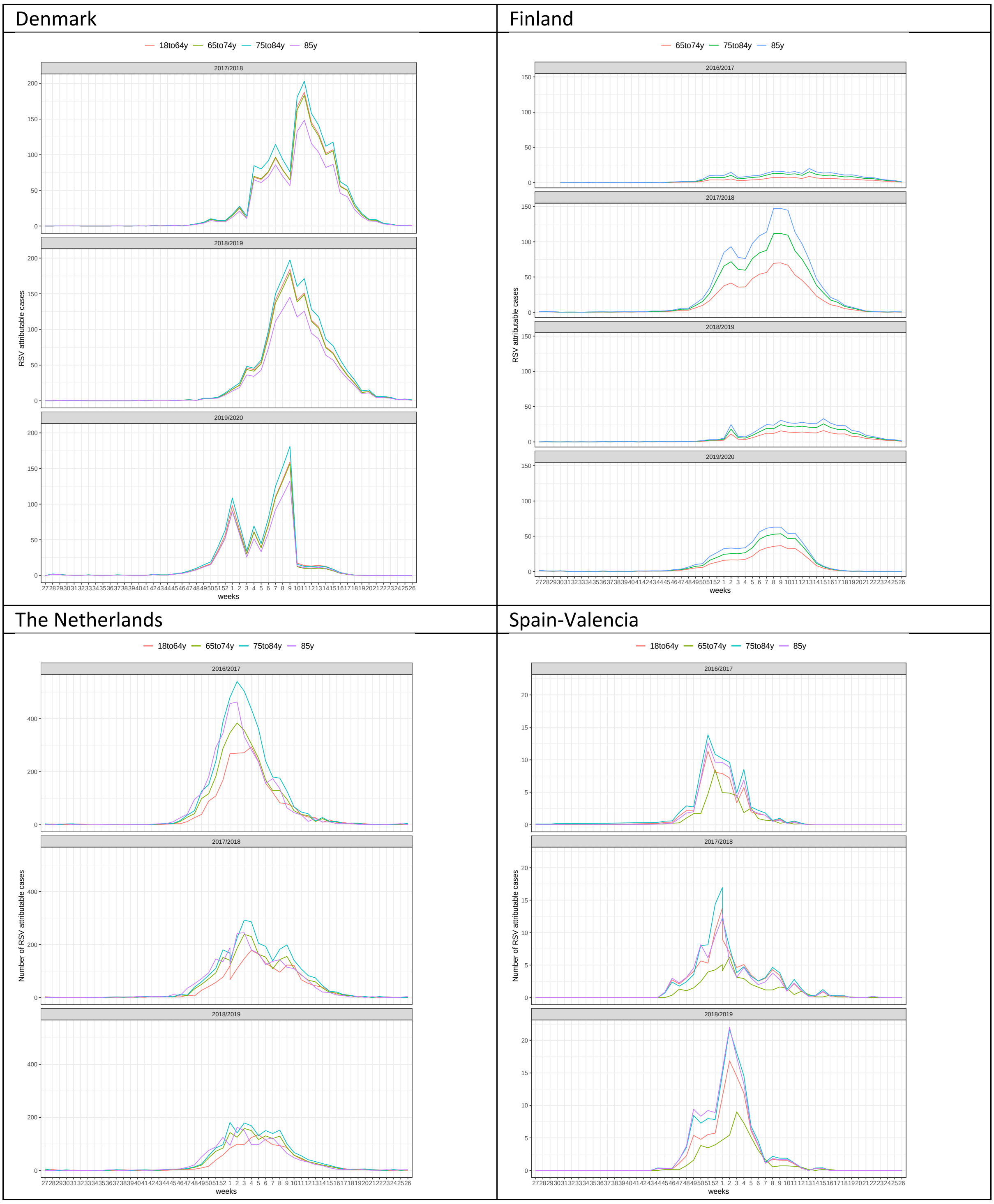

##### 1.1.2 RSV seasonality and the COVID-19 pandemic

A noticeable bi-annual pattern was observed before the COVID-19 pandemic in Finland and Spain-Valencia (RSV-confirmed data). Data from Finnish RSV-ICD-coded hospital admissions (S. Figure 1) indicated that a "severe" season has 50% more admissions than the average season, while a "weak" season has 50% fewer admissions than the average. The protection derived from vaccines diminishes over time, hence, whether vaccine uptake occurring before a severe or weak season might matter, especially if waning efficacy would be large by the time the second season peaks. To explore this, we conducted two scenario analyses. In one scenario, the RSV vaccine is introduced before a "severe" season, followed by a "weak" season; in the other scenario, the order was reversed.

Evidence suggested that RSV seasonality did change during the COVID-19 pandemic in many countries [13]. The RSV-ICD-coded hospital admission data showed almost no RSV-ICD-coded hospitalisations in the 2020/2021 season across all countries analysed (S. Figure 1). This can be attributed to social distancing as a consequence of voluntary and imposed behavioural changes through non-pharmaceutical interventions, such as school closure, lockdown, and use of face masks. RSV reappeared in the 2021/2022 season, which started earlier and was marked by an atypical peak in European countries for which data were available. For example, in Denmark, the peak was four months earlier (October) than the typical peak in February-March during pre-COVID-19 seasons. Hence, we included a scenario to investigate the prevented burden when a hospital admission peak of a country would shift four months earlier compared to the pre-COVID-19 era, followed by a 'typical' pre-COVID-19 peak. This is illustrated in S. Figure 5, where the first season peak is shifted to October-November and the second season is again a typical season peaking in February-March.

*S. Figure 5: Scenario analysis: An example of a typical (solid line) peak in Denmark prior to the COVID-19 pandemic and an "atypical" peak (dotted line) as observed in season 2021-2022*

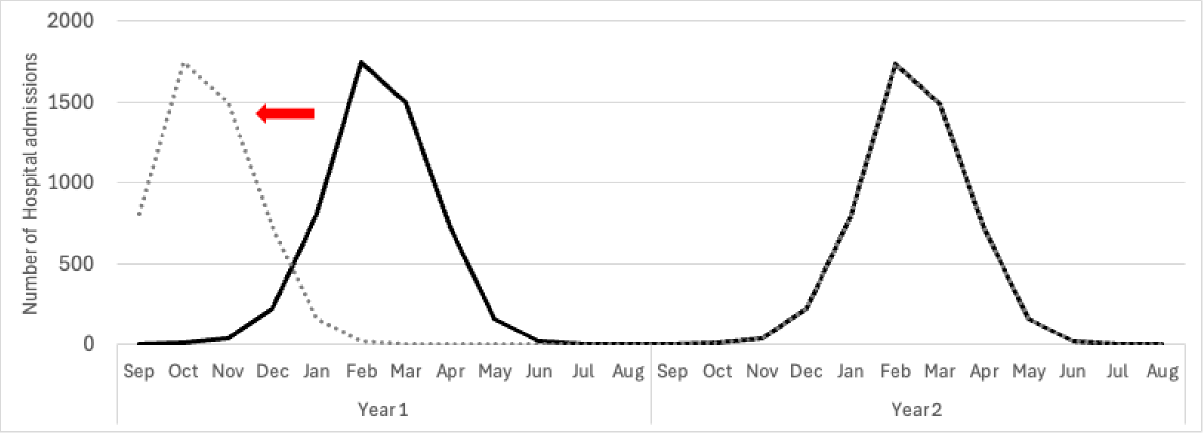

##### 1.1.3 RSV-related in-hospital deaths

Three systematic reviews and meta-analyses investigated the RSV deaths among older adults in high-income countries [8, 14, 15]. Savic and colleagues pooled nine studies in high-income countries and reported that the in-hospital case fatality ratio (hCFR) was 7.13% (95% CI: 5.4-9.36) among adults 60 years and above [15]. This estimate was higher than the hCFR of 5.3% (95% CI: 3.9-7.3) reported by Li and colleagues based on two studies [8], but lower than the case-fatality proportion estimated at 8.18% (95% CI: 5.54-11.94%) by Ngyuyen-Van-Tam and colleagues by pooling six studies [14]. The aims and selection criteria of the three systematic reviews were different. Li and colleagues aimed to estimate the case under-ascertainment with stricter selection criteria, whereas Ngyuyen-Van-Tam and colleagues also included a community-based study in addition to five studies in medically attended older adults. Therefore, for our primary analysis, we utilised the pooled non-age-specific hCFR estimate of 7.13% (95% CI: 5.4-9.36), of Savic and colleagues [15].

RSV laboratory-confirmed in-hospital deaths data were available in Denmark, Finland and Spain-Valencia. Due to the small sample size and GDPR, we could not report the age-specific hCFR in Denmark and Spain-Valencia. In Finland, there were 20, 42, 99 and 172 RSV laboratory-confirmed RSV deaths (in hospital) registered in adults 50-64 years, 65-74 years, 75-84 years and 85+ years, respectively, aggregated over four seasons (2016-2017 to 2019-2020).

We estimated the age-specific hCFR using a Beta (α,β) distribution where α represents the number of RSV laboratory-confirmed deaths and β the number of RSV laboratory-confirmed hospitalisations minus the number of RSV laboratory-confirmed deaths. S. Figure 6 illustrates the age-specific hCFR. Given the uncertainty around hCFR, we also performed a scenario analysis using age-specific hCFR estimates from Finland in all countries.

*S. Figure 6: The estimated age-specific in-hospital case fatality ratios based on Finnish RSV laboratory-confirmed data from 2016/2017 to 2019/2020. The red dotted line represents the average 7.13% hCFR, which is used in the reference case.*

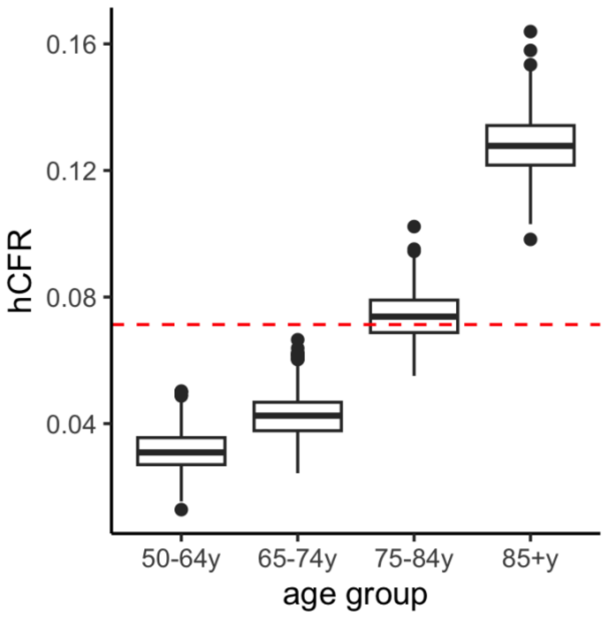

##### 1.1.4 RSV-related primary care and non-medically attended episodes

RSV-related primary care data were limited in most countries considered due to limited laboratory testing in the European primary care setting [16]. Therefore, we assumed the ratio of 8.5 (range from 7.0 to 9.5) for primary care episodes per RSV hospitalisation in adults aged 60 years and above, based on a meta-analysis of US studies conducted by McLaughlin and colleagues [17]. In the scenario analysis, the ratio was set to be 12.5 (range from 4.5-20.5), roughly based on a UK modelling study conducted by Fleming and colleagues [18]. The uncertainty around these estimates was sampled from a uniform distribution.

RSV-related non-medically attended (non-MA) episodes were estimated based on a multi-country prospective cohort study among community-dwelling older adults in Europe (Belgium, the UK and the Netherlands; sample size N=1040) [10, 19]. The study reported that out of 36 RSV patients, 25 patients had non-MA episodes and 11 patients had outpatient episodes, hence, a ratio of 2.27 was used to estimate non-MA episodes based on primary care episodes. We specified a ±50% uniform distribution, from which we sampled values to acknowledge this ratio's uncertainty.

##### 1.1.5 Vaccine efficacy, duration of protection and waning curve

Both RSV vaccines, Arexvy® and Abrysvo®, have different case definitions and primary end-points defined in their phase 3 trials (Table 1 main text), which makes the comparison very challenging [20, 21]. Additionally, the full second season vaccine efficacy data of Arexvy® have been published, reporting a median follow-up time of 18 months [22], whereas the second season efficacy data of Abrysvo® have been press-released with a median follow-up time of 16.4 months [23] [24, 25], but a full report or a scientific publication was not available at the time of analysis. Given these challenges to align product-specific characteristics using scant data, we specified the vaccine in our economic evaluation as if it were a hypothetical RSV vaccine for older adults for exploratory purposes only. Performing an economic evaluation on one product, when there are two or more competing products for the same target group and disease, would substantially limit the policy implications the analysis yields for decision makers, even if all information on this single product and the disease burden estimates would be available. However, it helps understanding which data gaps and uncertainties are most influential. The aim of our analysis was, therefore, to identify and explore key assumptions and sources of uncertainty that would affect the evaluation of any RSV vaccine for this target group. As more data on various products emerge, enabling accurate direct comparisons, our insights into critical assumptions and uncertainties will expedite future analyses.

We found several assumptions for the protection of RSV vaccines over two years in different cost-effectiveness analyses of RSV vaccines in older adults [24, 26-28]. During the June 2023 United States Advisory Committee on Immunization Practices (ACIP) meeting, a stepwise function in combination with linear waning was used in the Centers for Disease Control and Prevention (CDC) model and Pfizer’s model, however, in the GSK model, the vaccine wanes linearly [24, 26, 29]. In another cost-effectiveness analysis in the US, both continuous temporal (sigmoid) and periodically constant (stepwise in combination with linear) waning were investigated [27]. Therefore, we explored various waning scenarios with multiple types of waning accommodating a range for the duration of protection from 24 months to 48 months (S. Table 1 in main text and S. Figure 7).

Bearing in mind our hypothetical vaccine, in line with data available at the time of study, is defined by only two data points per clinical endpoint to fit to, we fitted the efficacy over time using a linear function and truncating negative values at 0 and values exceeding 1 at 1. In the reference scenario 0, we conservatively assumed linear waning up to 24 months and no efficacy after 24 months. However, in the scenario analysis, we also assumed linear waning up to 36 months and 48 months (scenario 1 and 2). We also explored exponential waning applying a similar approach while truncating values above 1 (scenario 3-5). One minus exponential waning was investigated by constraining vaccine efficacy to attain 0 by 24 months (scenario 6) or 36 months (scenario 7), and effectively creating a third data point at 0. We then fitted the three data points using one minus an exponential function. In addition, we examined the stepwise waning in two scenarios. Scenario 8 assumed the season one efficacy would protect throughout the first year, and the season two efficacy would protect throughout the second year, whereas scenario 9 assumed the season one efficacy would protect eight months and season two efficacy would protect for 9-18 months, based on the median follow-up of the efficacy data. Finally, we evaluated the stepwise waning in combination with linear waning, assuming after 18 months, the efficacy would decrease linearly to 0 by 24 months, 36 months and 48 months (scenarios 10-12). The uncertainty ranges of the season one and season two efficacy points were first obtained by sampling from a beta (ranked) distribution, and then the uncertainty ranges of the waning curves were estimated by fitting the efficacy data points to the corresponding curves (i.e., linear, exponential or stepwise).

*S. Table 1: List of waning scenarios*

| # | Scenarios | Linear fitting (truncated) | Exponential fitting | 1-Exponential decay | Stepwise |
|---|---|---|---|---|---|
| Reference scenario 0 | Linear waning (truncated): only capture 24 months | Fit a linear line based on the 2 seasons' efficacy data | | | |
| Scenario 1 | Linear waning (truncated): only capture 36 months | | | | |
| Scenario 2 | Linear waning (truncated): only capture 48 months | | | | |
| Scenario 3 | Exponential waning: only capture 24 months | | Fit an exponential curve based on the 2 seasons efficacy data | | |
| Scenario 4 | Exponential waning: only capture 36 months | | | | |
| Scenario 5 | Exponential waning: only capture 48 months | | | | |

| | | | | | |
|---|---|---|---|---|---|
| Scenario 6 | 1-exponential decay wane to 0 by 24 months | | | Use 3 data points to fit 1-exponential curve: the two available data points and 0 as the 3rd data point | |
| Scenario 7 | 1-exponential decay wane to 0 by 36 months | | | | |
| Scenario 8 | Stepwise: full protection over 2 years | | | | Season 1: 1-12 months<br>Season 2: 13-24 months |
| Scenario 9 | Stepwise: protection to data availability (18 months) | | | | Season 1: 1-8 months<br>Season 2: 9-18 months |
| Scenario 10 | Stepwise + linear waning up to 24 months | | | | Linear waning to 0 as from the last data point |
| Scenario 11 | Stepwise + linear waning up to 36 months | | | | |
| Scenario 12 | Stepwise + linear waning up to 48 months | | | | |

*S. Figure 7: Graphic presentation of the waning scenarios*

Linear waning (truncated, reference scenario 0 and scenario 1-2) & exponential waning (scenario 3-5)

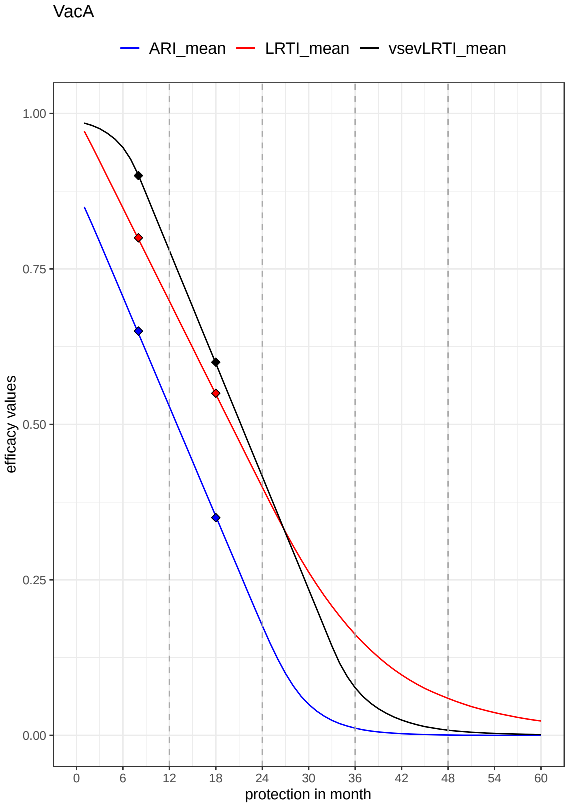

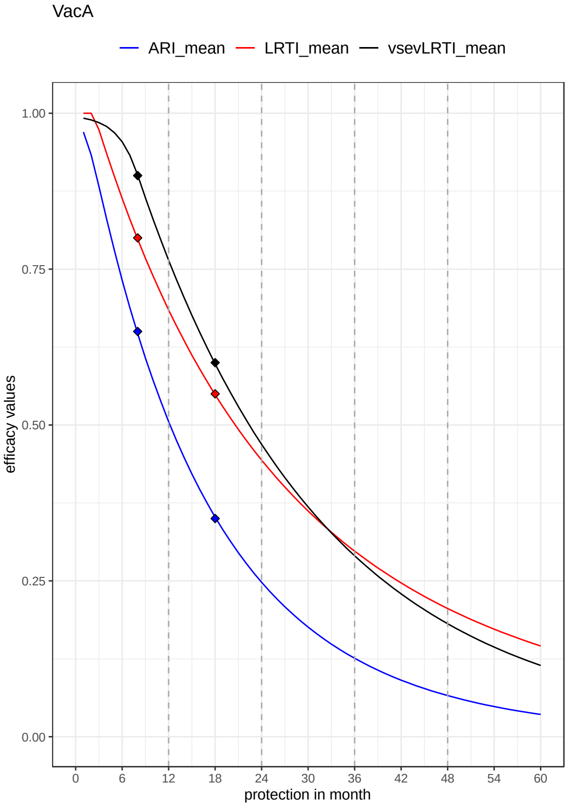

One minus exponential waning (scenario 6 and 7)

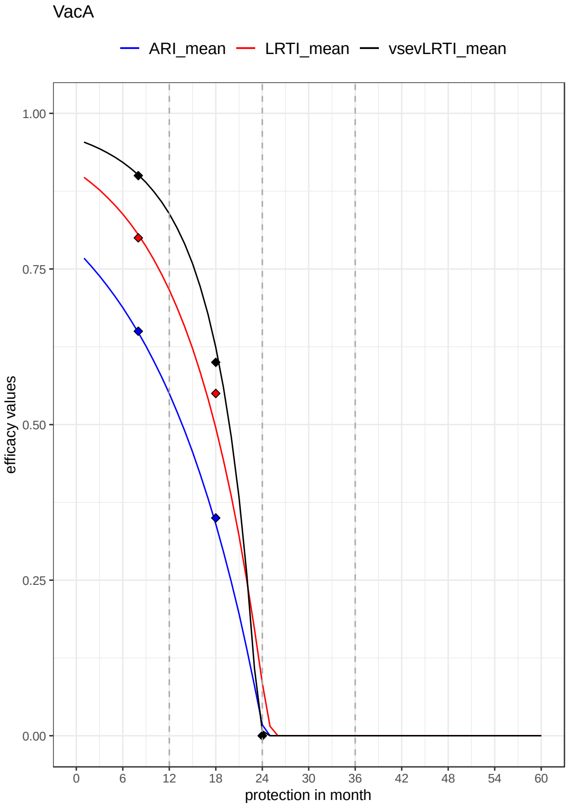

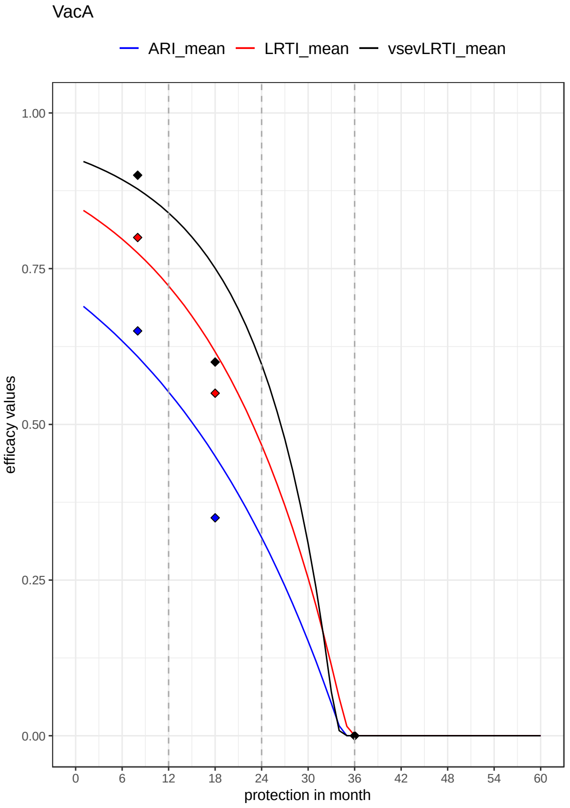

Stepwise waning (scenario 8 and 9)

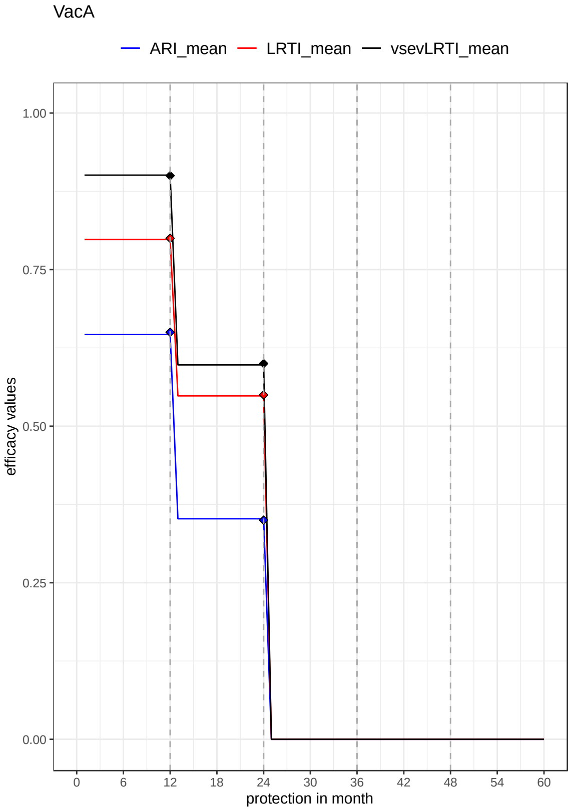

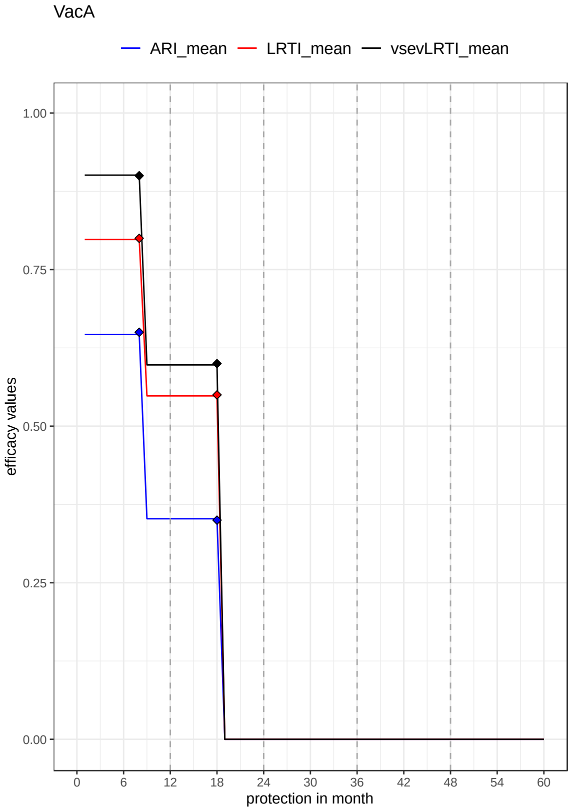

#### Stepwise plus linear waning (scenario 10 to 12)

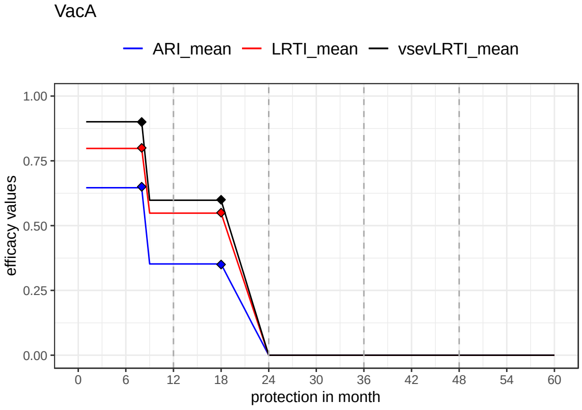

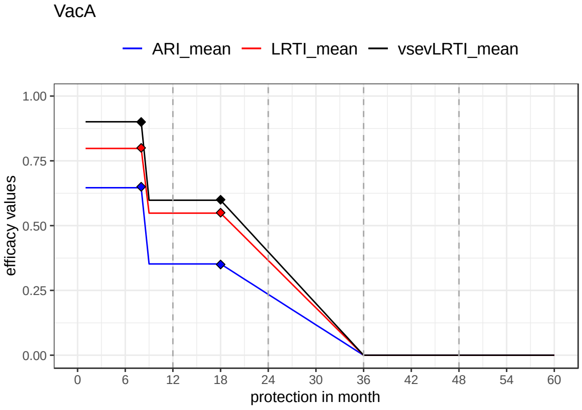

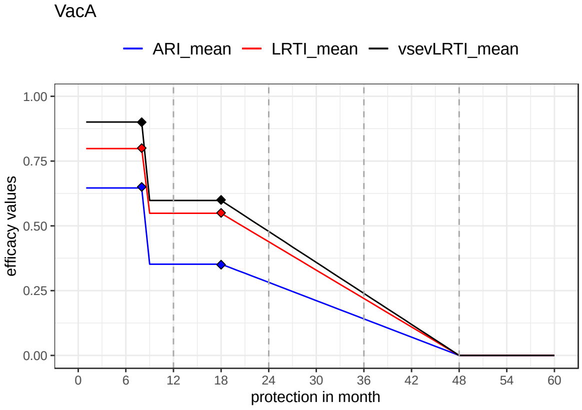

##### 1.1.6 Efficacy in older adults by age

The efficacy data reported for both RSV vaccines by age group are shown in S. Table 2 [20-22]. The confidence intervals for vaccine efficacy in the 80 years and above age group are extremely large, implying these data provide no evidence as to whether RSV vaccine efficacy would be lower or higher in adults aged over 80 years of age compared to the younger adult age groups. Clinical and immunological research indicates a reduction in immune responses among older adults to many vaccines, including influenza vaccination [30, 31], but more recently developed vaccines are trying to overcome this through a variety of approaches [32]. We assumed the efficacy values would be constant for all age groups (main text Table 1), but we conducted a scenario analysis assuming vaccine efficacy values were halved in the 80 years and above age group.

*S. Table 2: Phase 3 vaccine efficacy data for Arexvy® and Abrysvo® by open-ended and specific 10-year age group.*

| | 60y + | 70y+ | 80y+ | 60-69 | 70-79 |
|---|---|---|---|---|---|
| Arexvy (GSK) | | | | | |
| Season 1 | | | | | |
| Sample | V N=12467<br>P N=12499 | V N=5504<br>P N=5519 | V N= 1017<br>P N=1028 | V N=6963<br>P N=6980 | V N=4487<br>P N=6427 |
| Mean age | 69.5+/- 6.5 | | | | |
| Efficacy LRTD* | 82.6%<br>(57.9-94.1) | 84.4% | 33.8 %<br>(-477.7-94.5%) | 81%<br>(43.6-95.3) | 93.8%<br>(60.2-99.9) |
| Season 2 only | | | | | |
| Sample | V N=6228<br>P N=12503 | V: N = 2748<br>P: N= 5527 | V N=504<br>P N=1028 | V N=3480<br>P N=6982 | V N=6982<br>P N=4493 |
| Mean age | 69.5 (6.4) | | | | |
| Efficacy LRTD* | 56.1<br>(28.2-74.4) | 66.2<br>(18.9-88.3) | 41.0<br>(-209.9–94.0) | 50.9<br>(6.1-76.3) | 66.2<br>(18.9–88.3) |
| Abrysvo (Pfizer) | | | | | |
| Season 1 | | | | | |
| Sample | V N= 17215<br>P N=17069 | V N=6758<br>P N=6389 | V N=970<br>P N=958 | V N=10757<br>P N=10680 | V N=5788<br>P N=5431 |
| Mean age | 68.3 | | | | |
| Efficacy LRTD ≥2 symptoms | 66.7<br>(28.8-85.8) | | 80%<br>(-104.3-99.7) | 57.9<br>(-7.4-85.3) | 77.8%<br>(-18.7-98.1) |
| Efficacy LRTD ≥3 symptoms | 85.7<br>(32-98.7) | | 100%<br>(-191.2-100) | 77.8%<br>(-18.7-98.1) | 100%<br>(-573.8-100) |

** Efficacy LRTD: LRTD ≥2 symptoms including ≥1 sign or ≥3 symptoms*

*Abbreviation: V: vaccine, P: placebo, LRTD: lower respiratory tract disease*

##### 1.1.7 Direct and indirect costs

All costs were inflated to their 2023 value using the country-specific consumer price index (CPI) of all sectors [33], and those reported in local currency were converted to euro (€) using the annual exchange rates of 2023. In our analysis, direct costs include costs per non-MA episode, per primary care episode, per hospitalisation, per vaccine dose and per administration of a vaccine dose from the HCP perspective.

The costs per primary care episode for each country was based on the cost of one general practitioner (GP) consultation (S. Table 6).

Regarding the intervention cost per dose, we assumed €150 per dose for an RSV vaccine based on list price of Arexvy® in the UK (£150) [34], but varied this assumption in a price sensitivity analysis between €50 and €250. We also assumed that the RSV vaccine would be delivered in the beginning of October (with instant protection), hence, we used country-specific coverage and the administration cost of influenza vaccine as proxies for the RSV vaccine.

From a societal perspective, we also considered the indirect costs associated with productivity losses due to RSV illness using the human capital approach. Productivity costs per day were estimated by mean income per year divided by 12 months with 22 working days per month and adjusted by country-specific employment rate per age group. In line with standard practice and guidelines for cost-utility analysis, when using the human capital approach (not the friction cost approach) under a societal perspective, productivity losses are valued as costs in the numerator of the cost-utility ratio through the number of illness days lost, but the future loss of work days due to mortality is not monetised, because it is already included in the QALY gains (the denominator of the ICER), and the policy maker's WTP threshold value should be set accordingly [35]. Therefore, productivity losses due to RSV-associated premature deaths were not included, when using human capital approach in order to avoid double counting.

Details on how the country-specific cost parameters were obtained/estimated are explained in the following paragraphs.

###### *1.1.7.1 Denmark*

###### Cost per primary care consultation, RSV-related hospitalisation per episode and length of stay

The average cost per primary care consultation is based on the latest Danish national tariff [36]. It was assumed to be a standard consultation plus a common procedure cost for ARI patients (CRP, code 7120). The hospitalisation cost per average admitted RSV episode is €3398 (2022 value) in Denmark, which includes intensive care unit (ICU) stays. This cost was extracted from a previous publication that analysed the diagnosis-related group (DRG) rate of upper respiratory tract infections among older adults [37]. The mean length of stay with RSV illness in Denmark was unavailable, hence, we used the mean length of stay data from Finland (see section below).

###### Cost of RSV vaccine administration

The influenza vaccine in older adults is mainly delivered by GPs in Denmark, hence, a standard GP consultation cost was assumed for the administration cost of RSV vaccine.

##### 1.1.7.2 Finland

###### Cost per primary care consultation, RSV-related hospitalisation per episode and length of stay

The unit costs for healthcare resource use in Finland were obtained from a national pricelist (Mäklin and Kokko 2020 [38]). The cost per RTI-related primary care consultation is €73.65. S. Table 3 shows the mean cost per RSV-confirmed hospitalisation, which includes the costs of ICU stays. The mean hospital length of stay with RSV illness was 9.16 days in the 65-74 years age group.

*S. Table 3: Age-specific RSV-confirmed hospitalisation cost (including intensive care unit costs) in 2023 value*

| Age group | Mean cost | Standard deviation |
|---|---|---|
| 18-64 years | €2928 | 2065 |
| 65-74 years | €3440 | 2457 |
| 75-84 years | €3628 | 2500 |
| 85+ years | €2928 | 2427 |

###### Cost of RSV vaccine administration

In Finland, wellbeing services counties are responsible for the practical arrangements for vaccinations included in the national vaccination programme. Most influenza vaccinations have been administered during organised vaccination events by nurses. Public health nurses are responsible for vaccinating people of all ages. We assumed RSV vaccines in older adults would be administered jointly with influenza vaccines, implying the labour costs can be allocated based on the time associated with administering each vaccine. Based on a Finnish time-and-motion study [39], the administration of a vaccine takes 5–10 minutes (includes preparation, administration and documentation), hence, the marginal RSV vaccine administration costs were estimated as €4.74 (10 minutes), based on the salary of a public health nurse.

##### 1.1.7.3 The Netherlands

###### Cost per primary care consultation, RSV-related hospitalisation per episode and length of stay

The primary care consultation cost, hospitalisation cost and ICU admission cost per day was extracted from the latest version of the Dutch costing manual [40] valued at the 2021 price level, which updated to the 2023 price level using CPI as shown in S. Table 6 [33]. The RSV-related hospital costs per episode including ICU stays by age group are illustrated in S. Table 4. We used the length of stay with RSV illness in patients aged 50-64 years to estimate the productivity losses for hospitalised patients.

*S. Table 4: Mean length of stay data from 2016/2017 season to 2019-2020 season and estimated cost per hospitalisation episode including intensive care unit (ICU) admission (2023 € value)*

| Age | Mean hospitalisation LOS (including ICU) days | %ICU | Cost per RSV hospitalisation episode |
|---|---|---|---|
| 50-64 years | 8.49 | 11% | € 8529 |
| 65-74 years | 8.19 | 10% | € 7994 |
| 75-84 years | 8.37 | 6% | € 7370 |
| 85+ years | 7.94 | 2% | € 6166 |

###### Cost of RSV vaccine administration

In the Netherlands, GPs receive reimbursement of €14.01 or €21.97 for vaccinating the indicated patients in the context of the National Flu Prevention Program or for the National Pneumococcal Vaccination Program for Adults, respectively [41]. Since we assumed the RSV vaccine would be given together with influenza vaccine prior to the influenza/RSV season, we applied the reimbursement rate of influenza vaccine.

We also performed a specific scenario analysis (only for the Netherlands) that accounted for the unrelated medical costs in life-years gained offered by RSV vaccination programmes, according to the Dutch pharmacoeconomic guideline. Using the Practical Application to Include Disease Costs (PAID) tool, we estimated the discounted age-specific healthcare cost unrelated to ‘influenza or pneumonia’ using the remaining life-expectancy at age of death for each death averted by vaccination [42].

###### 1.1.7.4 Spain-Valencia

###### Cost per primary care consultation, RSV-related hospitalisation per episode and length of stay

Based on the official regional bulletins from 2023, the cost per primary care consultation is €63 for the first visit [43]. S. Table 5 illustrates the cost per RSV-related hospitalisation by age group based on the Valencia active hospital surveillance data. These costs per average admitted RSV episode (including ICU stays) are comparable with a retrospective hospital database analysis [44]. Utilising active surveillance data from Valencia, the average length of stay with RSV illness for patients aged 65 years and above was estimated to be 5 days before the COVID-19 era.

*S. Table 5: Age-specific RSV-confirmed hospitalisation cost per episode (including intensive care unit costs) in 2023 value*

| Age group | Mean cost per episode | Standard deviation |
|---|---|---|
| 0-64 years | € 5245 | 10785 |
| 65-79 years | € 4019 | 2695 |
| 80y+ years | € 3815 | 1605 |

###### Cost of RSV vaccine administration

The cost of the RSV vaccine administration was assumed to be the same as the cost of administering influenza vaccine. Based on a recent publication in 2021, this cost is €14.7 (2020 value) in adults aged 65 years and above [45].

##### 1.1.8 Health-related quality of life

The EQ-5D index population norms by age group were derived from a cross-country analysis of population surveys for 20 countries (table: country-specific TTO and VAS value sets)

[46]. The baseline utility values were categorised by age group: 55-64 years, 65-74 years and 75 years and above. We fitted a linear spline using mid-point age to estimate the baseline utility value over a one-year age interval. The country-specific baseline utility values are listed in S. Table 6.

##### 1.1.9 List of parameters used in this analysis

All the parameters used in this analysis are listed in S. Table 6.

*S. Table 6: Input parameters*

| Common input parameters (for all 4 countries) | | |
|---|---|---|
| Parameter | Value [range] | Reference /sources /distribution (if any) |
| RSV disease burden | | |
| RSV-related non-medically attended (non-MA) cases | Non-MA episode per primary care visit: 2.27 [1.14-3.41] | RESCEU prospective study [10, 19]<br>Uniform distribution |
| RSV-related primary care visits | Primary care visits per hospitalisation<br>Base case: 8.5 [7.0-9.5]<br>Scenario analysis:<br>12.5 [4.5-20.5] | Base case: McLaughlin 2020 [17]<br>Scenario analysis: Fleming 2015 [18]<br>Uniform distribution |
| Hospital case fatality ratio | Base case: 60+y:7.13% (95% CI: 5.4-9.36)<br>Sensitivity analysis: age-specific rate based on Finnish data | Base case: Savic 2022 [15]<br>Lognormal distribution<br>Scenario analysis: details in section 1.2.1<br>Beta distribution |
| Quality-adjusted life-year | | |
| QALY loss per non-MA case | 0.004 [0.001-0.01] | Base case: Mao and Li 2022 [19]<br>Gamma distribution |
| QALY loss per primary care visit | Base case: 0.004 [0.001-0.01]<br>Sensitivity analysis: 0.0193 (0.0095-0.0316) | Base case: Mao and Li 2022 [19]<br>Scenario analysis: US ACIP meeting [24]<br>Gamma distribution |
| QALY loss per hospital admission | Base case: 0.01023 [0.0089 – 0.0117]<br>Sensitivity analysis: 0.0185 (0.0053-0.0347) | Base case: Mao 2023, hospital admission value based on the utility value of RSV infant patients' parents [47].<br>Scenario analysis: US ACIP meeting [24]<br>Gamma distribution |
| Unit Costs (€ 2023 value) | | |
| Intervention cost per dose | €150<br>range: €50 - €250 | Assumption based on the UK list price of £150 per dose [34], varied in probabilistic price threshold analysis and scenario analysis |
| Cost per non-MA episode | €4.06 | Cost of over-the-counter medication. Mao and Li et al. 2022 [19] |
| Productivity losses (days) | | |
| Productivity losses due to non-MA episode | 1 day | RESCEU prospective study [10, 19] |
| Productivity losses due to primary care episode | 2 days | RESCEU prospective study [10, 19] |

| | | |
|---|---|---|
| Productivity losses due to hospitalisation | Hospital length of stay +3 day | Assumption |
| Intervention characteristics | | |
| Efficacy against non-medically attended cases | Season 1: 65% (35-85%)<br>Season 2: 35% (15-60%) | Details see section 1.2.4 |
| Efficacy against primary care visit | Season 1: 65% (35-85%)<br>Season 2: 35% (15-60%) | |
| Efficacy against hospital admission | Season 1: 80% (35-95%)<br>Season 2: 55% (25-75%) | |
| Efficacy against deaths | Season 1: 90% (60-100%)<br>Season 2: 60% (55-90%) | |
| Duration of protection | Base case: 24 months<br>Sensitivity analysis: 36 and 48 months | Details see section 1.2.4 |
| Waning curve | Base case: linear curve<br>Scenario analysis: exponential, 1-exponential, stepwise, stepwise plus linear | Details see section 1.2.4 |
| Country specific input parameters | | |
| Denmark | | |
| Parameter | Values | Reference |
| Target population | 60-64y: 358,338<br>65-69y: 325,352<br>70-74y: 298,794<br>75-79y: 284,851<br>80-84y: 169,568<br>85y+: 134 764 | Statistics Denmark [48] |
| Life expectancy and general probability of deaths (age-specific: (1-year interval) | Life expectancy by age: range from 23.65 (60y) to 1.98 (99y)<br>Probability of death by age range from 0.0076 (60y) to 0.3695 (99y) | Average life expectancy 2018:2022<br>Life table, Statistics Denmark [49] |
| Age-specific baseline utility values | 55-64y: 0.870<br>65-74y: 0.847<br>75y+: 0.794 | EQ-5D index value population norms by age group [46] |
| RSV disease burden | | |
| RSV hospital admissions | - Age-specific RSV-attributable admissions by calendar month estimated with TSM<br>- Age-specific RSV-coded hospital admissions by calendar month | Danish Patient Registry 2016-2020, four seasons average prior to COVID-19 era (Details in section 1.4.1) [1, 2] |
| Cost (2023 value) | | |
| Exchange rate Danish Krone to Euro | 7.44 | Annual exchange rates 2022 Eurostat [50] |
| Cost of hospitalisation per episode | €3 545 | Average cost of upper respiratory infection [37] |
| Cost per primary care consultation | €30.25 (Danish Krone 153.61+71.48) | HONORARTABEL (tariff) 2022 [36] regular GP consultation (code 0101) and a procedure CRP (code 7120) |
| Average hospital length of stay | Same as Finland | Assumption |
| Mean income per day for age 60 years and above | €110.4 | Eurostat [23] |

| | | |
|---|---|---|
| Employment rate by age | 60-64y: 60%<br>65-69y: 19% | Eurostat [51] |
| Vaccine administration costs | €21 (Danish Krone 153.61) | Regular GP consultation (code 0101) |
| Coverage | 7-64y: 28.5%<br>65y: 78% | Influenza vaccine coverage [52] |
| Discounting | | |
| Discount rate | 3.5% both for effects and costs | Danish pharmacoeconomic guideline [53] |
| Finland | | |
| Parameter | Value | Reference/source |
| Target population | 60-64y: 354 703<br>65-69y: 349 928<br>70-74y: 343 733<br>75-79y: 273 662<br>80-84y: 168 074<br>85-89y: 100 950<br>90-94y: 46 307<br>95-99y: 10 755 | Population 2022, Statistics Finland [54] |
| Life expectancy and general probability of deaths (age-specific: (1-year interval) | Life expectancy by age: range from 23.88 (60y) to 1.98 (99y)<br>Probability of death by age range from 0.00654 (60y) to 0.3657 (99y) | Life table, Statistics Finland [55] |
| Age-specific baseline utility values | 55-64y: 0.81<br>65-74y: 0.79<br>75y+: 0.68 | Finnish study EQ-5D index value population norms [56] |
| Disease burden | | |
| RSV hospital admissions | - Age-specific RSV-attributable admissions by calendar month estimated from TSM<br>- Age-specific RSV-coded hospital admissions by calendar month | Finland Patient Registry 2016-2020, four seasons average peri-COVID-19<br>(Details in section 1.4.1) [1, 2] |
| Cost (€ 2023 value) | | |
| Cost of hospitalisation per episode | 18-64y: €2928 (SD: 2065)<br>65-74y: €3440 (SD: 2457)<br>74-84y: €3628 (SD: 2500)<br>85y+: €2928 (SD: 2427) | Mäklin S, Kokko P. (2020). Terveyden- ja sosiaalihuollon yksikkökustannukset Suomessa vuonna 2017 [38]<br>The cost included the intensive care unit costs. (Details in section 1.2.7.1) |
| Cost per primary care visit | €73.65 | Mäklin S, Kokko P. (2020). Terveyden- ja sosiaalihuollon yksikkökustannukset Suomessa vuonna 2017 [38] |
| Mean income per day for age 60 years and above | €104 | Eurostat [23] |
| Average hospital length of stay | 9.16 days | Care Register for Health Care 2016-2020 |
| Employment rate by age | 60-64y: 62.7%<br>65-69y: 19.1% | Statistics Finland, labor force survey [54] |
| Vaccine | | |
| Coverage | 50-64: 35%<br>65y+: 60% | Influenza vaccine coverage as proxy [54] |
| Administration costs of vaccine | €4.74 | 10 minutes public nurse time, Nieminen et al., 2022 [39]. |

| | | |
|---|---|---|
| Discounting | | |
| Discount rate | 3% both for effects and costs | Pharmacoeconomic guideline [57] |
| The Netherlands | | |
| Parameter | Value | Reference/Source |
| Target population | 60-64y: 1 181 421<br>65-69y: 1 030 621<br>70-74y: 927 219<br>75-79y: 770 710<br>80-84y: 470 814<br>85+y: 401 803 | Eurostat [58] |
| Life expectancy and general probability of deaths (age-specific: (1-year interval) | Life expectancy by age: range from 24.40 (60y) to 1.93 (99y)<br>Probability of death by age range from 0.00536 (60y) to 0.3657 (99y) | Statistics Netherlands [59] [60] |
| Age-specific baseline utility values | 55-64y: 0.890<br>65-74y: 0.886<br>75y+: 0.83 | EQ-5D index value population norms by age group [46] |
| RSV disease burden | | |
| RSV hospital admissions | - Age-specific RSV-attributable admissions by calendar month estimated with TSM<br>- Age-specific RSV-coded hospital admissions by calendar month | Netherlands Patient Registry 2016-2020 (Details in section 1.4.1), prior to COVID-19 pandemic. [1, 2] |
| Costs (€ 2023 value) | | |
| Cost of hospitalisation per day | €721 per hospital day | Hakkaart-van Roijen et al. 2024 [40] (Details on costs of hospitalisation in section 1.4.4.6) |
| Costs of ICU per day | €3 054 per day | |
| Cost per primary care consultation | €34.57 | |
| Hospital length of stay (days) | 50-64y: 8.49<br>65-74y: 8.19<br>75-84: 8.37<br>85y+: 7.94 | Netherlands Patient Registry 2012-2020 (Details in section 1.4.1), prior to COVID-19 pandemic |
| Average productivity cost per hour per day # | €46.64 per hour * 7.5 hour per day | Hakkaart-van Roijen et al. 2024 [40] |
| Employment rate by age | 60-64y: 58.09%<br>65-69y: 14.11% | Eurostat [51] |
| Vaccine administration costs | €14.01 | Based on administration cost of vaccination influenza vaccine among adults 60 years and above in the Netherlands [41] |
| Influenza vaccine coverage | 60-64y: 40.1%<br>65y: 68.4% | Vaccine Coverage Dutch National Influenza Prevention Program 2022 [51] |
| Discounting | | |
| Discount rate | 1.5% for effects and 3% for costs | Hakkaart-van Roijen et al. 2024 [40] |
| Spain-Valencia | | |
| Target population (Valencia) | 60-64y: 69 452<br>65-69y: 59 391<br>70-74y: 50 839<br>75-79y: 44 167<br>80-84y: 29 675<br>85-89y: 19 509 | This is the population of the active surveillance in Valencia, it represents ~21% of the population in Valencia.[1, 2] |

| | | |
|---|---|---|
| | 90-94y: 9 421<br>95+y: 2 572 | |
| Life expectancy and general probability of deaths (age-specific: (1-year interval) | Life expectancy by age:<br>50y: 34<br>60y: 25<br>70y: 17<br>80y: 10<br>Probability of death by age (nqx): 0.006 (60y) to 0.30 (99y) | Spanish National Statistics Institute [61] |
| Age-specific baseline utility values (Spain) | 55-64y: 0.901<br>65-74y: 0.891<br>75y+: 0.781 | EQ-5D index value population norms by age group [46] |
| Cost (2023 value) | | |
| Cost of hospitalisation per episode | 60-69y: €5 245 (SD: 10785)<br>70-79y: €4 019 (SD: 2695)<br>80+y: €3 815 (SD: 1605) | Valencia hospital active surveillance database |
| Cost of primary care visit | €63 | Official regional bulletins, first GP consultation cost [43] |
| Hospital length of stay | 5 days | Valencia Patient Registry 2016-2020 (Details in section 1.4.1) |
| Average productivity cost per hour per day # | €75 per day | Eurostat [23] |
| Employment rate by age | 60-64y: 58%<br>65-69y:17% | Eurostat [51] |
| Vaccine administration costs | €16.5 | Assuming same administration cost as influenza vaccine [45] |
| Vaccine coverage | 60-64y: 51.9%<br>65y: 67.2% | Influenza vaccination coverage in Valencia general population [62] |
| Discounting | | |
| Discount rate | 3% both for effects and costs | Spanish Recommendations on Economic Evaluation of Health Technologies (2010) [63] |

#### 1.2 Cost-effectiveness analysis

Our study applied the concept of dominance and extended dominance based on the incremental cost-effectiveness ratio (ICER), and the concept of expected net loss curves to determine the cost-effective strategy for a given willingness-to-pay (WTP) value [64]. The expected net loss was calculated in the model over a range of WTP threshold values (€0 to €150 000) and the results for each alternative program were compared through the expected net loss curves (ENLCs). The preferred strategy was the strategy with the lowest expected net loss (equal to the program with the highest expected incremental net benefit). The expected net loss for the cost-effective strategy is the same as the expected value of perfect information (EVPI) when the preferred (cost-effective) strategy is chosen. We used EVPI as a measure of decision uncertainty. Expected net loss (ENL) accounts for both the probability of making a non-preferred (i.e., wrong) decision and the cost consequences of making that wrong decision. Higher EVPI indicates more expected losses; hence, higher decision uncertainty.

A detailed description of state-of-the-art ways to represent uncertainties in health economic evaluation can be found in Alarid-Escudero et al. 2019 [64] and Bilcke and Beutels 2021 [65]. We also presented for each strategy a cost-effectiveness acceptability curve based on 1000 simulations for a range of WTP values which shows the probability that a program is cost-effective. The ICER was also calculated for the preferred strategy by dividing the mean incremental costs by the mean incremental QALYs gained compared to the next-best strategy. Strategies that were at least as expensive and less effective (strongly dominated strategies) and weakly dominated strategies were excluded from the cost-effectiveness analyses.

#### 1.3 Expected Value of Partial Perfect Information (EVPPI)

The expected value of partial perfect information (EVPPI) for each uncertain input parameter were obtained independently in order to identify the input parameters that contribute most to decision uncertainty. Parameters with the highest EVPPI are most influential. Note that the impact of the uncertainty of other parameters was evaluated in a list of scenario analyses (see section 2.5).

#### 1.4 Scenario analyses

A large number of scenario analyses were performed to explore the impact of key assumptions and input parameters. An overview of vaccine waning-related scenarios is shown inS. Table 1, and a list of additional scenarios unrelated to the assumed waning characteristics is summarised below in S. Table 7.

*S. Table 7: List of scenarios unrelated to the assumed waning characteristics*

| Abbreviations | Full name | Explanation |
|---|---|---|
| ISO3 codes of the country (from HCP) Time series | Country analysis using TSM hospitalisation estimates from HCP | - Vaccine protection: 2 years linear waning<br>- Hospitalisations: TSM estimates, except Spain-Valencia: RSV-confirmed data<br>- hCFR: Non-age-specific (7.13%)<br>- Seasonality: Average 3 or 4 seasons prior to COVID-19 pandemic<br>- Primary care ratio vs. hospitalisation: 8.5 (7.0 to 9.5)<br>- Price: €150 per dose<br>- QALY values: RESCEU study<br>- Vaccine efficacy: Non-age-specific<br>- Vaccine coverage: Age-specific 65 years and 60-64 years |
| Adjusted ICD-coded | ICD-coded hospital admission without adjustment factor | Hospitalisations: ICD-coded hospital admission data multiplied by the 2.2 adjustment factors |
| ICD-coded | ICD-coded hospital admission with adjustment factor | Hospitalisations: ICD-coded hospital admission data |
| Adjusted Lab-confirmed | Laboratory-confirmed hospital admission with adjustment factor | Hospitalisations: laboratory-confirmed hospital admission data multiplied by the 2.2 adjustment factors |
| Lab-confirmed | Laboratory-confirmed hospital admission without adjustment factor | Hospitalisations: laboratory-confirmed hospital admission data |
| Time series (societal) or Lab confirmed | Country analysis from a societal perspective | Perspective: societal |
| Age-specific hCFR | Age-specific hCFR | hCFR: Age-specific hCFR from Finnish data |
| Severe mild season | Introduce RSV vaccine in a severe RSV season | Seasonality: RSV vaccine introduced in a “severe” first season (50% higher than average incidence of infections), followed by a “mild” season (50% lower than average) |
| Mild severe season | Introduce RSV vaccine in a mild RSV season | Seasonality: RSV vaccine introduced in a “mild” first season (50% lower than average), followed by a “severe” season (50% higher than average) |
| PeriCOVID season | RSV seasonality changed during the COVID-19 pandemic | Seasonality: RSV peak shifted 4 months earlier than the “typical” pre-COVID-19 peak |
| Higher Primary care ratio | A higher primary care ratio vs. hospitalisation | Primary care ratio vs. hospitalisation: 12.5 (4.5-20.5) |
| Higher price (EUR 250) | Higher vaccine price | Price: €250 per dose |
| Lower price (EUR 50) | Lower vaccine price | Price: €50 per dose |

| | | |
|---|---|---|
| Higher QALY loss | Higher QALY losses per episodes | QALY values: US CDC presentation |
| Lower efficacy at 80y+ | Lower vaccine efficacy in adults 80 years and above | Vaccine efficacy: 50% lower than the overall vaccine efficacy in population 80 years and above |
| Higher coverage: 60y to 64y | Higher vaccine coverage in 60-64y | Vaccine coverage: 60-64 years have same coverage as 65 years and above |

### 2 SUPPLEMENT RESULTS

#### 2.1 Using adjusted RSV-ICD-coded hospitalisations

##### 2.1.1 Adjusted RSV-ICD-coded disease and economic burden without vaccination

*S. Table 8: Mean [95% CI] of adjusted RSV-coded disease and economic burden in adults over 60 years for each country without intervention over a 5-year time horizon. The adjusted RSV-ICD-coded estimates were used for all countries. All results are from the healthcare payers' perspective unless otherwise stated. Both costs and quality-adjusted life-years were discounted at country-specific discount rates. All costs are presented in €'000 in 2023 value.*

| Denmark | | | | | |
|---|---|---|---|---|---|
| | 60-64 years | 65-74 years | 75-84 years | 85 years + | 60 years + |
| Undiscounted outcomes | | | | | |
| Non-MA episodes | 1 614 [797 - 2564] | 6 166 [3044 - 9795] | 6 974 [3442 - 11078] | 6 696 [3305 - 10638] | 21 450 [10588 - 34076] |
| Primary care episodes | 637 [521 - 764] | 2 433 [1991 - 2920] | 2 752 [2252 - 3302] | 2 643 [2163 - 3171] | 8 465 [6927 - 10157] |
| Hospitalisations | 75 [70 - 80] | 286 [268 - 306] | 323 [303 - 346] | 310 [291 - 332] | 995 [931 - 1064] |
| Deaths | 5 [4 - 7] | 21 [15 - 27] | 23 [17 - 31] | 22 [16 - 29] | 72 [52 - 94] |
| QALY losses | 106 [80 - 139] | 303 [229 - 395] | 210 [157 - 280] | 114 [81 - 168] | 733 [551 - 955] |
| Direct medical costs ('000 €) | 291 [272 - 312] | 1 112 [1039 - 1194] | 1 258 [1176 - 1350] | 1 208 [1129 - 1296] | 3 869 [3616 - 4152] |
| Costs of productivity loss ('000 €) | 296 [222 - 381] | 166 [125 - 214] | 0 [0 - 0] | 0 [0 - 0] | 463 [346 - 595] |
| Total costs from societal perspective ('000 €) | 587 [499 - 683] | 1 278 [1176 - 1391] | 1 258 [1176 - 1350] | 1 208 [1129 - 1296] | 4 331 [3990 - 4702] |
| Discounted costs and QALYs | | | | | |
| QALY losses | 79 [60 - 103] | 244 [184 - 317] | 186 [139 - 252] | 109 [77 - 162] | 618 [463 - 829] |
| Direct medical costs ('000 €) | 290 [271 - 311] | 1 105 [1033 - 1186] | 1 249 [1168 - 1341] | 1 198 [1120 - 1286] | 3 842 [3591 - 4124] |
| Costs of productivity loss ('000 €) | 295 [221 - 379] | 165 [124 - 213] | 0 [0 - 0] | 0 [0 - 0] | 460 [344 - 592] |
| Total costs from societal perspective ('000 €) | 585 [497 - 679] | 1 270 [1169 - 1382] | 1 249 [1168 - 1341] | 1 198 [1120 - 1286] | 4 302 [3964 - 4671] |
| Finland | | | | | |
| | 60-64 years | 65-74 years | 75-84 years | 85 years + | 60 years + |
| Undiscounted outcomes | | | | | |
| Non-MA episodes | 14 205 [7012 - 22566] | 52 412 [25871 - 83261] | 67 448 [33293 - 107146] | 41 282 [20377 - 65580] | 175 347 [86554 - 278554] |
| Primary care episodes | 5 606 [4587 - 6726] | 20 685 [16926 - 24817] | 26 618 [21782 - 31936] | 16 292 [13332 - 19547] | 69 202 [56627 - 83026] |
| Hospitalisations | 659 [617 - 705] | 2 430 [2276 - 2600] | 3 127 [2928 - 3346] | 1 914 [1792 - 2048] | 8 130 [7613 - 8699] |
| Deaths | 47 [35 - 62] | 175 [128 - 229] | 225 [165 - 295] | 138 [101 - 181] | 586 [428 - 767] |
| QALY losses | 865 [652 - 1128] | 2 286 [1726 - 2975] | 1 771 [1320 - 2403] | 616 [422 - 940] | 5 538 [4154 - 7304] |

| | | | | | |
|---|---|---|---|---|---|
| Direct medical costs ('000 €) | 2 421 [934 - 5797] | 10 243 [3658 - 28313] | 13 355 [4896 - 34345] | 6 814 [2453 - 18740] | 32 832 [17740 - 58945] |
| Costs of productivity loss ('000 €) | 1798 [1345 - 2312] | 759 [568 - 976] | 0 | 0 | 2 556 [1913 - 3288] |
| Total costs from societal perspective ('000 €) | 4 219 [2555 - 7687] | 11 001 [4365 - 29330] | 13 355 [4896 - 34345] | 6 814 [2453 - 18740] | 35 389 [19925 - 61801] |
| Discounted costs and QALYs | | | | | |
| QALY losses | 672 [507 - 876] | 1 912 [1437 - 2487] | 1 604 [1181 - 2219] | 592 [400 - 911] | 4 780 [3561 - 6450] |
| Direct medical costs ('000 €) | 2 410 [929 - 5770] | 10 188 [3638 - 28162] | 13 276 [4867 - 34142] | 6 767 [2436 - 18611] | 32 640 [17638 - 58581] |
| Costs of productivity loss ('000 €) | 1 789 [1339 - 2301] | 754 [564 - 970] | 0 | 0 | 2 543 [1903 - 3272] |
| Total costs from societal perspective ('000 €) | 4 199 [2543 - 7651] | 10 942 [4342 - 29173] | 13 276 [4867 - 34142] | 6 767 [2436 - 18611] | 35 183 [19808 - 61459] |
| The Netherlands | | | | | |
| | 60-64 years | 65-74 years | 75-84 years | 85 years + | 60 years + |
| Undiscounted outcomes | | | | | |
| Non-MA episodes | 21 494 [10610 - 34145] | 86 336 [42617 - 137152] | 78 348 [38674 - 124462] | 33 695 [16632 - 53527] | 219 873 [108532 - 349287] |
| Primary care episodes | 8 483 [6941 - 10177] | 34 073 [27882 - 40880] | 30 920 [25302 - 37097] | 13 298 [10881 - 15954] | 86 774 [71006 - 104109] |
| Hospitalisations | 997 [933 - 1066] | 4 003 [3749 - 4283] | 3 633 [3402 - 3887] | 1 562 [1463 - 1672] | 10 194 [9546 - 10908] |
| Deaths | 72 [52 - 94] | 288 [211 - 378] | 262 [191 - 343] | 113 [82 - 147] | 734 [536 - 962] |
| QALY losses | 1 534 [1158 - 2002] | 4 717 [3555 - 6149] | 2 825 [2122 - 3674] | 708 [516 - 996] | 9 784 [7386 - 12748] |
| Direct medical costs ('000 €) | 8 880 [8307 - 9506] | 33 528 [31371 - 35888] | 28 159 [26358 - 30140] | 10 229 [9572 - 10964] | 80 796 [75620 - 86482] |
| Costs of productivity loss ('000 €) | 9 713 [7236 - 12531] | 5 369 [4000 - 6927] | 0 | 0 | 15 082 [11236 - 19459] |
| Total costs from societal perspective ('000 €) | 18 593 [15771 - 21631] | 38 897 [35902 - 42212] | 28 159 [26358 - 30140] | 10 229 [9572 - 10964] | 95 877 [88334 - 104365] |
| Discounted costs and QALYs | | | | | |
| QALY losses | 1 332 [1005 - 1739] | 4 240 [3206 - 5533] | 2 646 [1987 - 3450] | 687 [499 - 971] | 8 905 [6712 - 11563] |
| Direct medical costs ('000 €) | 8 845 [8274 - 9469] | 33 339 [31195 - 35686] | 27 981 [26192 - 29950] | 10 161 [9507 - 10891] | 80 326 [75181 - 85979] |
| Costs of productivity loss ('000 €) | 9 674 [7208 - 12482] | 5 337 [3976 - 6886] | 0 | 0 | 15 012 [11184 - 19368] |
| Total costs from societal perspective ('000 €) | 18 520 [15709 - 21546] | 38 676 [35699 - 41972] | 27 981 [26192 - 29950] | 10 161 [9507 - 10891] | 95 338 [87835 - 103780] |

*Abbreviations: QALY: quality-adjusted life-year, MA: medically attended*

##### 2.1.2 Effects of vaccination on adjusted RSV-ICD-coded disease and economic burden, and the associated costs

The adjusted RSV-ICD-coded disease and economic burden averted by each strategy compared to no intervention and the associated intervention costs are presented in S. Table 9 for each country. The mean and 95% credible intervals (CIs) are based on 1000 samples drawn for probabilistic sensitivity analysis. The 60y+ strategy offered the highest disease burden averted, hence, also the highest discounted QALYs gained, and the highest direct and indirect medical costs averted. However, this was the most expensive strategy. The direct medical costs averted, intervention costs and QALYs gained were similar between the 60y+ and 65y+ strategies; nevertheless, the 60y+ strategy led to the highest indirect costs averted as well as the highest intervention costs.

*S. Table 9: Using the adjusted RSV-ICD-10-coded data: mean [95% CI] discounted disease and economic burden averted over 2 years protection (truncated linear waning, 24 months protection) for vaccinating 60 years and above, 65 years and above and 75 years and above in October against RSV disease in adults compared to no intervention for each country. Both cost and quality-adjusted life-year were discounted at country-specific discount rates. All costs are presented in €'000 in 2023 value. All the results are from the healthcare payers' perspective unless otherwise stated. The assumed RSV vaccine price per dose is €150.*

| Denmark | 60 years + | 65 years + | 75 years + |
|---|---|---|---|
| non-MA cases averted | 3 587 [1396 - 6796] | 3 533 [1376 - 6692] | 2 574 [1008 - 4868] |
| Primary care episode averted | 1 412 [724 - 2148] | 1 391 [714 - 2115] | 1 013 [523 - 1537] |
| Hospitalisation averted | 217 [149 - 271] | 214 [147 - 267] | 155 [108 - 193] |
| Death averted | 17 [11 - 23] | 17 [11 - 23] | 12 [8 - 17] |
| QALY gained due to RSV non-MA episode averted | 14 [2 - 44] | 14 [2 - 43] | 10 [2 - 31] |
| QALY gained due to RSV MA episode averted | 8 [3 - 17] | 8 [3 - 17] | 6 [2 - 13] |
| QALY gained due to death averted | 105 [66 - 143] | 101 [64 - 138] | 55 [35 - 75] |
| Total QALYs gained | 127 [84 - 175] | 123 [81 - 170] | 71 [47 - 99] |
| Direct medical costs averted ('000€) | 816 [581 - 1012] | 803 [573 - 995] | 584 [418 - 721] |
| Intervention costs ('000€) | 195 469 | 178 006 | 94 757 |
| Incremental costs ('000€) | 194 654 [194458 - 194888] | 177 203 [177011 - 177433] | 94 174 [94036 - 94339] |

| | | | |
|---|---|---|---|
| Costs associated with productivity loss averted | 41 [25 - 62] | 30 [19 - 46] | 0 |
| Incremental costs from societal perspective (‘000€) | 194 613 [194405 - 194853] | 177 172 [176973 - 177407] | 94 174 [94036 - 94339] |
| Finland | 60 years + | 65 years + | 75 years + |
| non-MA cases averted | 23 384 [9225 - 43992] | 22 616 [8928 - 42541] | 16 563 [6566 - 31095] |
| Primary care episode averted | 9 206 [4791 - 13887] | 8 903 [4637 - 13425] | 6 521 [3412 - 9810] |
| Hospitalisation averted | 1 401 [981 - 1736] | 1 355 [949 - 1677] | 989 [697 - 1221] |
| Death averted | 110 [70 - 150] | 107 [68 - 145] | 78 [50 - 106] |
| QALY gained due to RSV non-MA episode averted | 93 [15 - 286] | 90 [14 - 276] | 66 [11 - 202] |
| QALY gained due to RSV MA episode averted | 51 [21 - 114] | 49 [20 - 110] | 36 [15 - 81] |
| QALY gained due to death averted | 628 [400 - 855] | 583 [371 - 794] | 317 [202 - 433] |
| Total QALYs gained | 772 [516 - 1069] | 722 [483 - 1001] | 419 [277 - 591] |
| Direct medical costs averted (‘000€) | 5 344 [2542 - 10030] | 5183 [2366 - 9941] | 3 725 [1605 - 8152] |
| Intervention costs (‘000€) | 139 296 | 120 085 | 55 683 |
| Incremental costs (‘000€) | 133 951 [129266 - 136753] | 114 902 [110144 - 117719] | 51 958 [47531 - 54078] |
| Costs associated with productivity loss averted | 211 [129 - 319] | 108 [66 - 163] | 0 |
| Incremental costs from societal perspective (‘000€) | 133 740 [128995 - 136582] | 114 794 [110004 - 117626] | 51 958 [47531 - 54078] |
| The Netherlands | 60 years + | 65 years + | 75 years + |
| Non-MA cases averted | 35 267 [14274 - 65281] | 34 326 [13912 - 63481] | 21 148 [8596 - 39027] |
| Primary care episode averted | 13 885 [7418 - 20573] | 13 515 [7229 - 20005] | 8 327 [4465 - 12297] |
| Hospitalisation averted | 2 074 [1507 - 2522] | 2 017 [1468 - 2450] | 1 240 [906 - 1503] |
| Death averted | 162 [104 - 219] | 157 [102 - 213] | 97 [63 - 131] |
| QALY gained due to RSV non-MA episode averted | 141 [23 - 426] | 137 [22 - 415] | 84 [14 - 256] |
| QALY gained due to RSV MA episode averted | 77 [32 - 171] | 75 [31 - 166] | 46 [19 - 102] |
| QALY gained due to death averted | 1 532 [989 - 2078] | 1 458 [942 - 1977] | 671 [434 - 909] |
| Total QALYs gained | 1 750 [1187 - 2375] | 1 669 [1132 - 2266] | 801 [543 - 1093] |
| Direct medical costs averted (‘000€) | 15 798 [11668 - 19120] | 15 301 [11327 - 18495] | 8 932 [6645 - 10760] |
| Intervention costs (‘000€) | 481 683 | 403 983 | 184 347 |
| Incremental costs (‘000€) | 465 885 [462562 - 470015] | 388 682 [385488 - 392656] | 175 415 [173587 - 177702] |
| Costs associated with productivity loss averted | 1 393 [851 - 2084] | 945 [581 - 1399] | 0 |
| Incremental costs from societal perspective (‘000€) | 464 492 [460833 - 468778] | 387 738 [384399 - 391816] | 175 415 [173587 - 177702] |

*Abbreviations: QALY: quality-adjusted life-year, MA: medically attended*

##### 2.1.3 Cost-effectiveness based on RSV-ICD-10-coded hospitalisation estimates

###### *2.1.3.1 Incremental cost-effectiveness plane*

The cost-effectiveness plane per country is shown in S. Figure 8, presenting the cost-effectiveness frontier and dominated strategies (i.e., strategies not lying on the cost-effectiveness frontier) when using the HCP and the societal perspective. From the HCP perspective, S. Figure 14 shows findings consistent with Figure 2 in the main text. From a societal perspective, the 60y+ and 65y+ strategies offer slightly lower incremental costs when accounted for productivity losses in all four countries (S. Table 9), but the overall findings are consistent with the HCP perspective.

*S. Figure 8: Incremental cost-effectiveness plane from healthcare payers' (left) and societal (right) perspectives. Adjusted RSV-ICD-coded data were used for all countries. The RSV vaccine price per dose assumed is €150.*

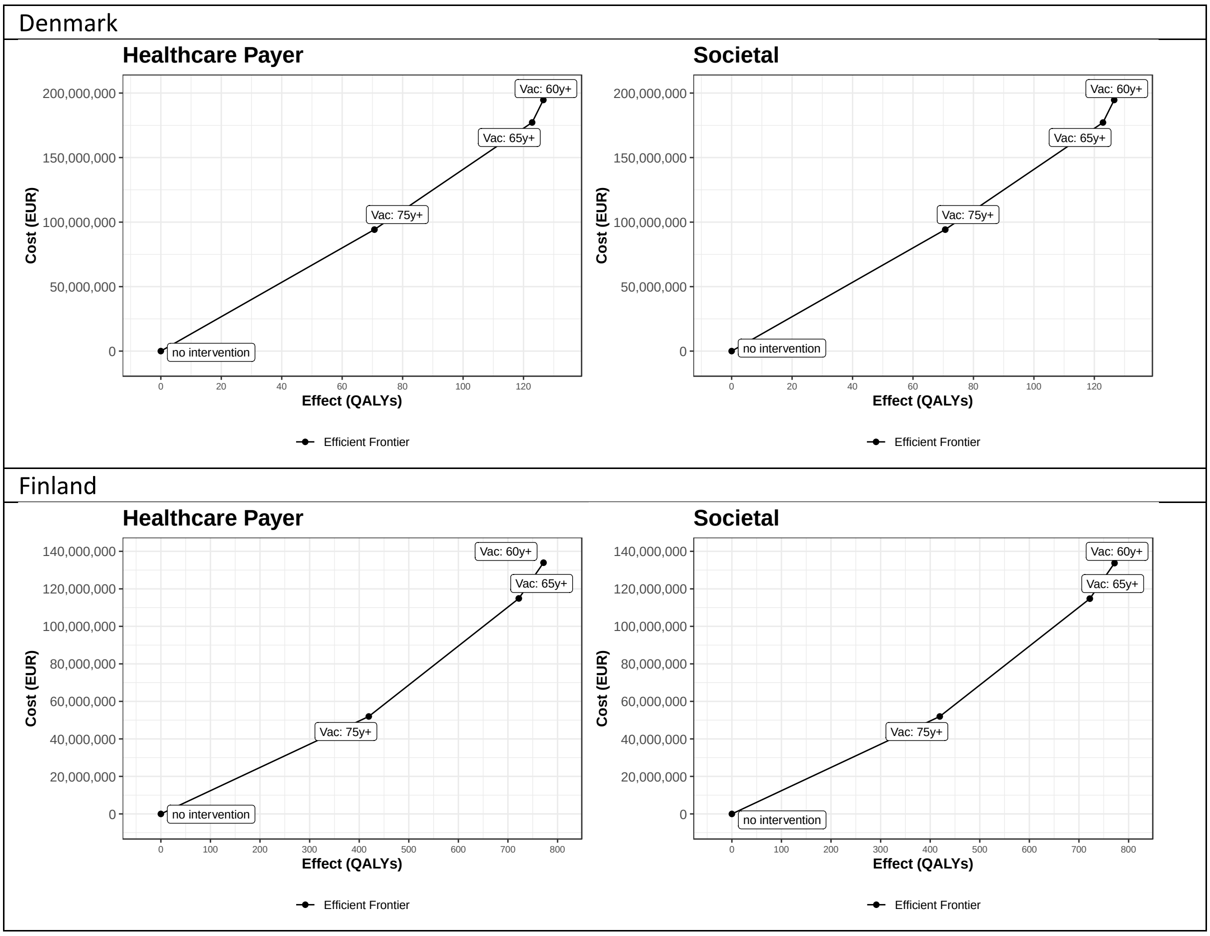

The Netherlands

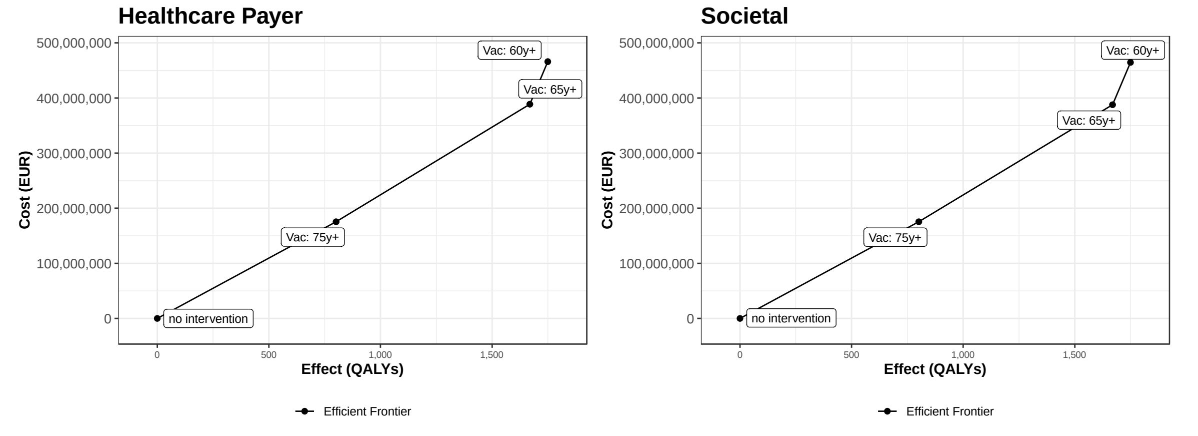

*Abbreviations: QALY: quality-adjusted life-year, EUR: euro*

##### 2.1.3.2 Cost-effectiveness acceptability curves (CEACs) and expected net loss curves (ENLCs)

CEACs and ENLCs inform the uncertainty surrounding the cost-effectiveness results. For each strategy, the probability to be cost-effective is shown using CEACs (S. Figure 9 left plots). For Denmark and the Netherlands, there is no decision uncertainty up to WTP values of €150,000 per QALY gained: no intervention is cost-effective with 100% probability (left plots) and there is no expected net loss of choosing no intervention (right plots). In Finland, for WTP values below €125,000 per QALY gained no intervention is the preferred strategy with 100% certainty (i.e., probability to be cost-effective is 100%), whereas for WTP values above €150,000 per QALY gained, the 75y+ strategy is preferred with 80% certainty.

ENLCs show the expected net loss (i.e., the expected cost of uncertainty) for each strategy. Expected net loss was highest for WTP values for which the choice between strategies was most uncertain. For Finland, the expected net loss reaches €4 million.

ENLCs also allow identification of the cost-effective program for a given WTP threshold. In the right-side panels of S. Figure 9 the preferred strategy for each WTP value is the strategy with the lowest expected net loss for that WTP value. The findings are consistent with Figure 2 in the main text and S. Figure 8S. Figure 14, but also emphasise the very high costs associated with choosing a non-preferred strategy within relatively broad ranges of WTP values in Finland.

*S. Figure 9: Using the adjusted RSV-ICD-10-coded data: Cost-effectiveness acceptability curves (left plots) and expected net loss curves (right plots) for each country comparing 3 strategies against RSV diseases in older adults: 60 years and above, 65 years and above and 75 years and above. All strategies were compared to no intervention and to each other. RSV-ICD-coded hospitalisation with adjustment factor were used for all countries. The assumed RSV vaccine price per dose is €150. The results are from healthcare payers' perspective.*

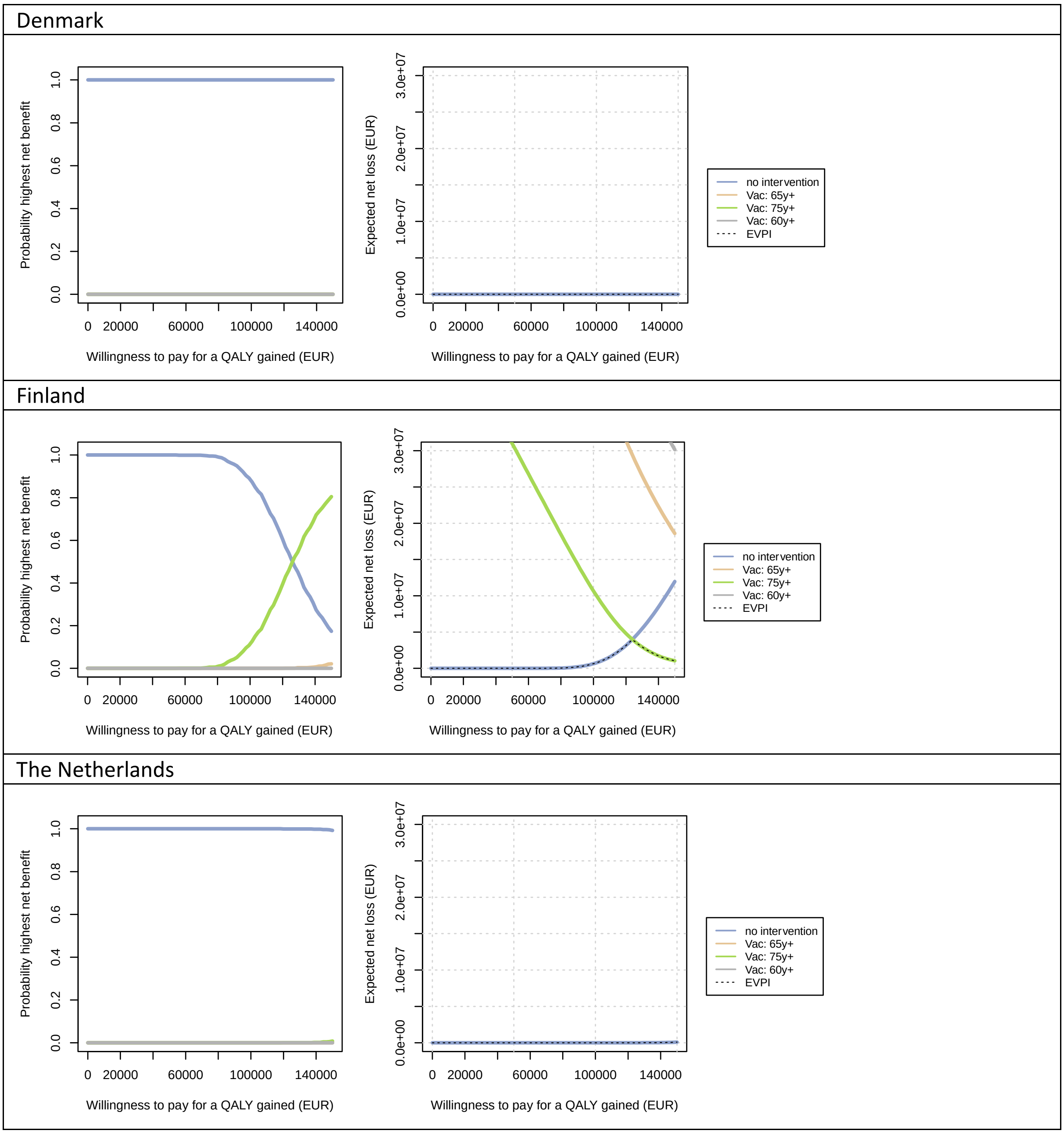

*Abbreviations: QALY: quality-adjusted life-year, EUR: euro*

##### 2.1.3.3 Expected value of partial perfect information (EVPPI)

For each country, the EVPPI was calculated for each uncertain input parameter across a range of WTP values to identify input parameters that contribute the most to decision uncertainty (i.e., uncertainty about the preferred strategy based on cost-effectiveness): the higher the EVPPI, the more influential. It is worth noting that the price is fixed at €150 per dose when calculating EVPPI, because price threshold analysis is conducted separately (see main text Figure 4). The EVPPI curves reach their peak at the WTP value at which the preferred strategy is most uncertain (i.e., preferred strategy changes).

Using adjusted RSV-ICD-coded data, no intervention is cost-effective in Denmark and the Netherlands up to a WTP value of €150,000 per QALY gained with 100% certainty, hence, the EVPPI graphs are flat (no decision uncertainty caused by uncertain input parameters). In Finland, the uncertainty around non-age-specific hCFR caused most decision uncertainty (i.e., with highest EVPPI; S. Figure 10). In addition to the uncertainty around age-specific hCFR, the uncertainty around the QALY losses due to non-MA, vaccine efficacy against mortality and primary care, proportion of non-MA visits and the adjustment factors became highly influential on which strategy is preferred for a limited range of WTP values (S. Figure 10 and main text Figure 6).

*S. Figure 10: Using adjusted RSV-ICD-coded data: the expected value of partial perfect information for each country (Finland's graph is reported in the main text). The assumed RSV vaccine price per dose is €150. The results are from healthcare payers' perspective.*

| Denmark |
|---|

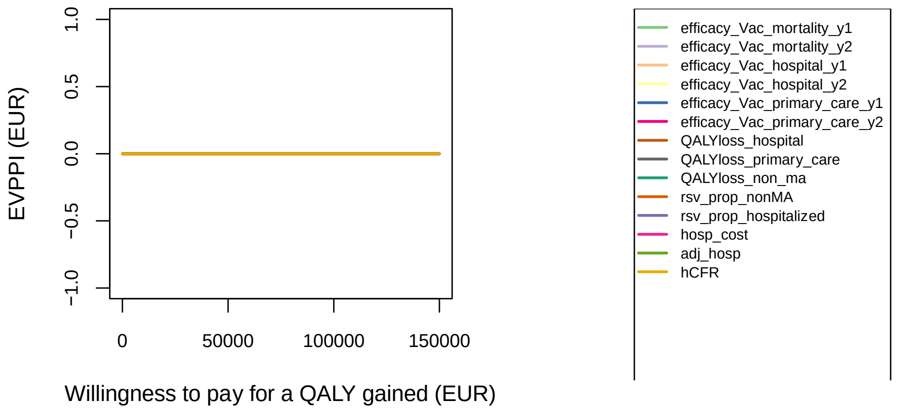

The Netherlands

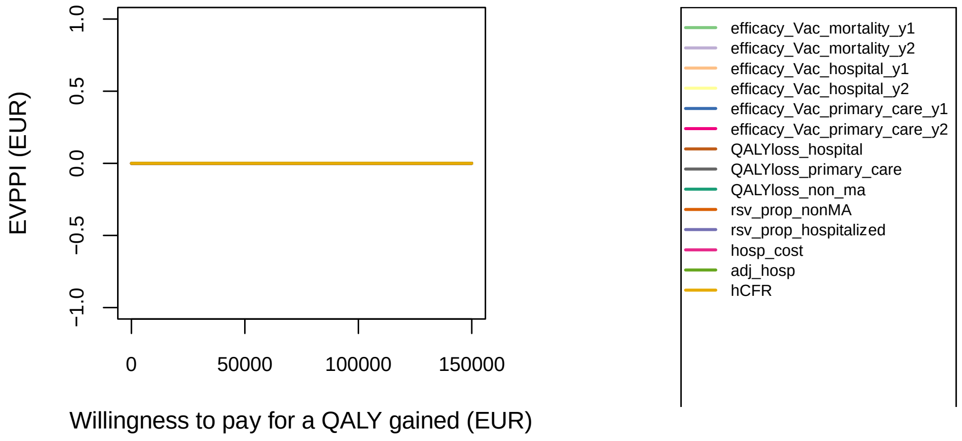

*Abbreviations: QALY: quality-adjusted life-year, EUR: euro, hCFR: in-hospital case fatality ratio, adj: adjustment factor, hosp: hospitalisation, RSV: respiratory syncytial virus, ma: medically attended, y: year*

#### 2.2 Using adjusted RSV-confirmed hospitalisations

##### 2.2.1 Adjusted RSV-confirmed disease and economic burden without vaccination

*S. Table 10: Mean [95% CI] of adjusted RSV-coded disease and economic burden in adults over 60 years for each country without intervention over a 5-year time horizon. The adjusted RSV-confirmed estimates were used for all countries. All results are from the healthcare payers' perspective unless otherwise stated. Both costs and quality-adjusted life-years were discounted at country-specific discount rates. All costs are presented in €'000 in 2023 value.*

| | | | | | |
|---|---|---|---|---|---|
| Denmark | | | | | |
| | 60-64 years | 65-74 years | 75-84 years | 85 years + | 60 years + |
| Undiscounted outcomes | | | | | |
| Non-MA episodes | 3 878 [1914 - 6160] | 16 328 [8060 - 25939] | 20 514 [10126 - 32588] | 18 164 [8966 - 28855] | 58 883 [29066 - 93541] |
| Primary care episodes | 1 530 [1252 - 1836] | 6 444 [5273 - 7731] | 8 096 [6625 - 9713] | 7169 [5866 - 8601] | 23 239 [19016 - 27881] |
| Hospitalisations | 180 [168 - 192] | 757 [709 - 810] | 951 [891 - 1018] | 842 [789 - 901] | 2 730 [2557 - 2921] |
| Deaths | 13 [9 - 17] | 55 [40 - 71] | 68 [50 - 90] | 61 [44 - 79] | 197 [144 - 258] |
| QALY losses | 254 [192 - 333] | 795 [600 - 1036] | 620 [465 - 827] | 310 [219 - 455] | 1 980 [1486 - 2582] |
| Direct medical costs ('000 €) | 699 [654 - 751] | 2 945 [2753 - 3160] | 3 700 [3458 - 3971] | 3276 [3062 - 3516] | 10 620 [9926 - 11397] |
| Costs of productivity loss ('000 €) | 712 [533 - 915] | 416 [311 - 535] | 0 | 0 | 1 128 [844 - 1451] |
| Total costs from societal perspective ('000 €) | 1 411 [1199 - 1640] | 3 361 [3094 - 3653] | 3 700 [3458 - 3971] | 3 276 [3062 - 3516] | 11 748 [10845 - 12720] |
| Discounted costs and QALYs | | | | | |
| QALY losses | 190 [143 - 247] | 642 [484 - 834] | 550 [410 - 743] | 296 [208 - 439] | 1 677 [1254 - 2257] |
| Direct medical costs ('000 €) | 696 [651 - 747] | 2 926 [2735 - 3141] | 3 675 [3435 - 3944] | 3 251 [3038 - 3489] | 10 548 [9859 - 11320] |
| Costs of productivity loss ('000 €) | 708 [530 - 911] | 413 [309 - 531] | 0 | 0 | 1 121 [839- 1442] |
| Total costs from societal perspective ('000 €) | 1 404 [1193 - 1632] | 3 339 [3075 - 3629] | 3 675 [3435 - 3944] | 3 251 [3038 - 3489] | 11 669 [10773 - 12635] |
| Finland | | | | | |
| | 60-64 years | 65-74 years | 75-84 years | 85 years + | 60 years + |
| Undiscounted outcomes | | | | | |
| Non-MA episodes | 18 733 [9247 - 29759] | 69 081 [34099 - 109741] | 86 902 [42896 - 138050] | 51 901 [25619 - 82449] | 226 616 [111861-359999] |
| Primary care episodes | 7 393 [6050 - 8870] | 27 263 [22309 - 32710] | 34 296 [28064 - 41148] | 20 483 [16761 - 24575] | 89 435 [73184 - 107302] |
| Hospitalisations | 869 [813 - 929] | 3 203 [2999 - 3427] | 4 029 [3773 - 4311] | 2 406 [2253 - 2575] | 10 507 [9839 - 11243] |
| Deaths | 63 [46 - 82] | 231 [168 - 302] | 290 [212 - 380] | 173 [127 - 227] | 757 [553 - 991] |
| QALY losses | 1140 [860 - 1487] | 3 017 [2277 - 3926] | 2 286 [1704 - 3102] | 775 [530 - 1181] | 7 217 [5414 - 9496] |

| | | | | | |
|---|---|---|---|---|---|
| Direct medical costs ('000 €) | 3 193 [1231 - 7645] | 13 500 [4821 - 37317] | 17 206 [6308 - 44251] | 8 566 [3084 - 23560] | 42 466 [22849 - 75620] |
| Costs of productivity loss ('000 €) | 2 371 [1774 - 3049] | 1 008 [754 - 1296] | 0 | 0 | 3 378 [2528 - 4346] |
| Total costs from societal perspective ('000 €) | 5 564 [3369 - 10137] | 14 508 [5760 - 38669] | 17 206 [6308 - 44251] | 8 566 [3084 - 23560] | 45 844 [25774 - 79937] |
| Discounted costs and QALYs | | | | | |
| QALY losses | 886 [669 - 1155] | 2 523 [1896 - 3281] | 2 070 [1524 - 2862] | 744 [503 - 1145] | 6 223 [4633 - 8387] |
| Direct medical costs ('000 €) | 3 178 [1226 - 7609] | 13 427 [4795 - 37117] | 17 104 [6271 - 43988] | 8 507 [3063 - 23397] | 42 217 [22710 - 75153] |
| Costs of productivity loss ('000 €) | 2 359 [1766 - 3035] | 1 002 [750 - 1289] | 0 | 0 | 3 361 [2515 - 4324] |
| Total costs from societal perspective ('000 €) | 5 538 [3353 - 10089] | 14 429 [5729 - 38460] | 17 104 [6271 - 43988] | 8 507 [3063 - 23397] | 45 579 [25623 - 79449] |
| Spain-Valencia | | | | | |
| | 60-64 years | 65-74 years | 75-84 years | 85 years + | 60 years + |
| Undiscounted outcomes | | | | | |
| Non-MA episodes | 3 817 [1884 - 6064] | 13 949 [6885 - 22159] | 22 016 [10868 - 34975] | 11 628 [5740 - 18472] | 51 410 [25377 - 81670] |
| Primary care episodes | 1 506 [1233 - 1807] | 5 505 [4505 - 6605] | 8 689 [7110 - 10425] | 4 589 [3755 - 5506] | 20 289 [16603 - 24343] |
| Hospitalisations | 177 [166 - 189] | 647 [606 - 692] | 1021 [956 - 1092] | 539 [505 - 577] | 2 384 [2232 - 2551] |
| Deaths | 13 [9 - 17] | 47 [34 - 61] | 74 [54 - 96] | 39 [28 - 51] | 172 [125 - 225] |
| QALY losses | 274 [207 - 357] | 715 [540 - 937] | 700 [525 - 921] | 204 [145 - 297] | 1 893 [1423 - 2461] |
| Direct medical costs ('000 €) | 1 037 [144 - 4753] | 3 042 [1055 - 8268] | 4 570 [2011 - 9671] | 2 372 [1205 - 4510] | 11 022 [6357 - 18804] |
| Costs of productivity loss ('000 €) | 242 [176 - 318] | 53 [38 - 69] | 0 [0 - 0] | 0 [0 - 0] | 295 [214 - 387] |
| Total costs from societal perspective ('000 €) | 1 280 [365 - 4978] | 3 095 [1111 - 8340] | 4 570 [2011 - 9671] | 2 372 [1205 - 4510] | 11 316 [6601 - 19189] |
| Discounted costs and QALYs | | | | | |
| QALY losses | 208 [157 - 271] | 589 [443 - 765] | 624 [466 - 841] | 194 [137 - 286] | 1 615 [1211 - 2129] |
| Direct medical costs ('000 €) | 1 032 [144 - 4730] | 3 027 [1049 - 8225] | 4 542 [1999 - 9612] | 2 356 [1197 - 4479] | 10 957 [6319 - 18701] |
| Costs of productivity loss ('000 €) | 241 [175 - 316] | 52 [38 - 69] | 0 | 0 | 293 [213 - 385] |
| Total costs from societal perspective ('000 €) | 1 273 [363 - 4954] | 3 079 [1106 - 8297] | 4 542 [1999 - 9612] | 2 356 [1197 - 4479] | 11 250 [6562 - 19084] |

*Abbreviations: QALY: quality-adjusted life-year, MA: medically attended*

##### 2.2.2 Effects of vaccination on adjusted RSV-confirmed disease and economic burden, and the associated costs

The adjusted RSV-confirmed disease and economic burden averted by each strategy compared to no intervention and the associated intervention costs are presented in S. Table 9 for each country. Given the differences in baseline disease burden estimates, the adjusted RSV-confirmed disease burdens averted were higher than the RSV-ICD-coded disease burdens averted for all three strategies in Denmark. It is worth noting that the intervention costs were the same (given that the target population remains the same), leading to overall less incremental direct and indirect costs compared to using adjusted ICD-coded hospitalisation. In Finland, the disease burdens are comparable between using adjusted RSV-ICD-coded and using adjusted RSV-confirmed hospitalisations. In Spain-Valencia, the findings are consistent with other countries that the 60y+ strategy offered the highest disease burden averted, highest discounted QALYs gained, the highest direct and indirect medical costs averted, and highest intervention costs.

*S. Table 11: Using the adjusted RSV-confirmed data: mean [95% CI] discounted disease and economic burden averted over 2 years protection (truncated linear waning, 24 months protection) for vaccinating 60 years and above, 65 years and above and 75 years and above in October against RSV disease in adults compared to no intervention for each country. Both cost and quality-adjusted life-year were discounted at country-specific discount rates. All costs are presented in €'000 in 2023 value. All the results are from the healthcare payers' perspective unless otherwise stated. The assumed RSV vaccine price per dose is €150.*

| Denmark | 60 years + | 65 years + | 75 years + |
|---|---|---|---|
| non-MA cases averted | 10 064 [3955 - 18973] | 9 940 [3909 - 18730] | 7 451 [2940 - 14017] |
| Primary care episode averted | 3 962 [2053 - 5993] | 3 913 [2029 - 5916] | 2 933 [1527 - 4425] |
| Hospitalisation averted | 605 [421 - 752] | 597 [416 - 742] | 446 [313 - 553] |
| Death averted | 48 [30 - 65] | 47 [30 - 64] | 35 [22 - 48] |
| QALY gained due to RSV non-MA episode averted | 40 [6 - 123] | 39 [6 - 121] | 30 [5 - 91] |
| QALY gained due to RSV MA episode averted | 22 [9 - 49] | 22 [9 - 48] | 16 [7 - 36] |
| QALY gained due to death averted | 287 [183 - 392] | 280 [178 - 381] | 161 [102 - 219] |

| | | | |
|---|---|---|---|
| Total QALYs gained | 349 [232 - 483] | 341 [227 - 472] | 206 [138 - 289] |
| Direct medical costs averted ('000€) | 2 274 [1638 - 2805] | 2 246 [1619 - 2769] | 1 680 [1217 - 2067] |
| Intervention costs ('000€) | 195 469 | 178 006 | 94 757 |
| Incremental costs ('000€) | 193 195 [192664 - 193831] | 175 760 [175237 - 176387] | 93 077 [92690 - 93540] |
| Costs associated with productivity loss averted | 102 [62 - 155] | 78 [48 - 118] | 0 |
| Incremental costs from societal perspective ('000€) | 193 093 [192536 - 193740] | 175 682 [175143 - 176317] | 93 077 [92690 - 93540] |
| Finland | 60 years + | 65 years + | 75 years + |
| non-MA cases averted | 30 196 [11909 - 56811] | 29 194 [11522 - 54917] | 21 149 [8378 - 39724] |
| Primary care episode averted | 11 887 [6184 - 17936] | 11 493 [5984 - 17332] | 8 326 [4352 - 12530] |
| Hospitalisation averted | 1 810 [1267 - 2242] | 1 749 [1225 - 2165] | 1 264 [889 - 1560] |
| Death averted | 142 [91 - 194] | 138 [88 - 187] | 99 [63 - 135] |
| QALY gained due to RSV non-MA episode averted | 120 [19 - 369] | 116 [19 - 356] | 84 [14 - 258] |
| QALY gained due to RSV MA episode averted | 66 [27 - 147] | 63 [26 - 142] | 46 [19 - 103] |
| QALY gained due to death averted | 820 [522 - 1116] | 761 [484 - 1036] | 408 [260 - 556] |
| Total QALYs gained | 1 005 [671 - 1393] | 940 [629 - 1302] | 538 [356 - 758] |
| Direct medical costs averted ('000€) | 6912 [3276 - 12997] | 6702 [3065 - 12896] | 4766 [2047 - 10537] |
| Intervention costs ('000€) | 139 296 | 120 085 | 55 683 |
| Incremental costs ('000€) | 132 383 [126298 - 136019] | 113 383 [107189 - 117021] | 50 917 [45146 - 53637] |
| Costs associated with productivity loss averted | 279 [170 - 420] | 144 [88 - 216] | 0 |
| Incremental costs from societal perspective ('000€) | 132 105 [125941 - 135788] | 113 239 [107018 - 116903] | 50 917 [45146 - 53637] |
| Spain-Valencia | 60 years + | 65 years + | 75 years + |
| Non-MA cases averted | 8 328 [3393 - 15345] | 7 976 [3251 - 14692] | 6 173 [2523 - 11351] |
| Primary care episode averted | 3 279 [1762 - 4835] | 3 141 [1688 - 4629] | 2431 [1310 - 3578] |
| Hospitalisation averted | 487 [358 - 590] | 467 [343 - 564] | 360 [266 - 435] |
| Death averted | 38 [25 - 51] | 36 [24 - 49] | 28 [18 - 38] |
| QALY gained due to RSV non-MA episode averted | 33 [5 - 100] | 32 [5 - 96] | 25 [4 - 75] |
| QALY gained due to RSV MA episode averted | 18 [7 - 40] | 17 [7 - 38] | 13 [6 - 30] |
| QALY gained due to death averted | 263 [170 - 356] | 240 [155 - 325] | 148 [96 - 200] |
| Total QALYs gained | 314 [213 - 428] | 288 [196 - 394] | 186 [126 - 258] |
| Direct medical costs averted ('000€) | 2 136 [1192 - 3563] | 2 019 [1107 - 3412] | 1 541 [831 - 2812] |
| Intervention costs ('000€) | 30 122 | 24 120 | 11 787 |
| Incremental costs ('000€) | 27 986 [26559 - 28930] | 22 101 [20708 - 23013] | 10 246 [8975 - 10956] |
| Costs associated with productivity loss averted | 32 [19 - 49] | 9 [5 - 14] | 0 |
| Incremental costs from societal perspective ('000€) | 27 954 [26525 - 28902] | 22 092 [20703 - 23003] | 10 246 [8975 - 10956] |

*Abbreviations: QALY: quality-adjusted life-year, MA: medically attended*

#### 2.2.3 Cost-effectiveness based on RSV-ICD-10-coded hospitalisation estimates

##### *2.2.3.1 Incremental cost-effectiveness plane*

S. Figure 11 shows the cost-effectiveness plane using adjusted RSV-confirmed data. From the HCP perspective, S. Figure 11shows findings consistent with Figure 3 in the main text. From both HCP and societal perspectives, the findings are consistent in the Denmark and Finland, comparing to using adjusted RSV-ICD-coded hospitalisations. In Spain-Valencia, the 75y+, 65y+ and 60y+ strategies are efficient when compared to no intervention and to each other from both perspectives.

*S. Figure 11: Incremental cost-effectiveness plane from healthcare payers' (left) and societal (right) perspectives. Adjusted RSV-confirmed data were used for all countries. The RSV vaccine price per dose assumed is €150.*

Denmark

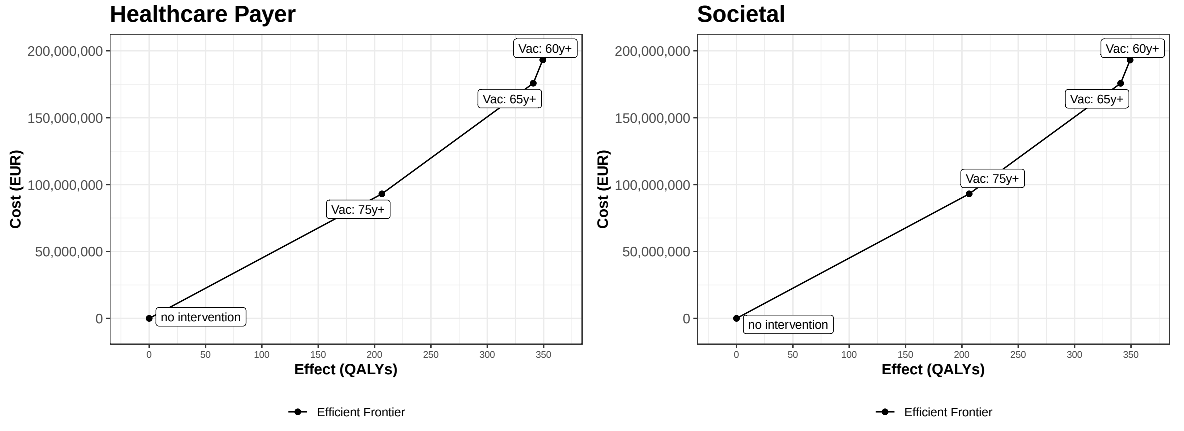

Finland

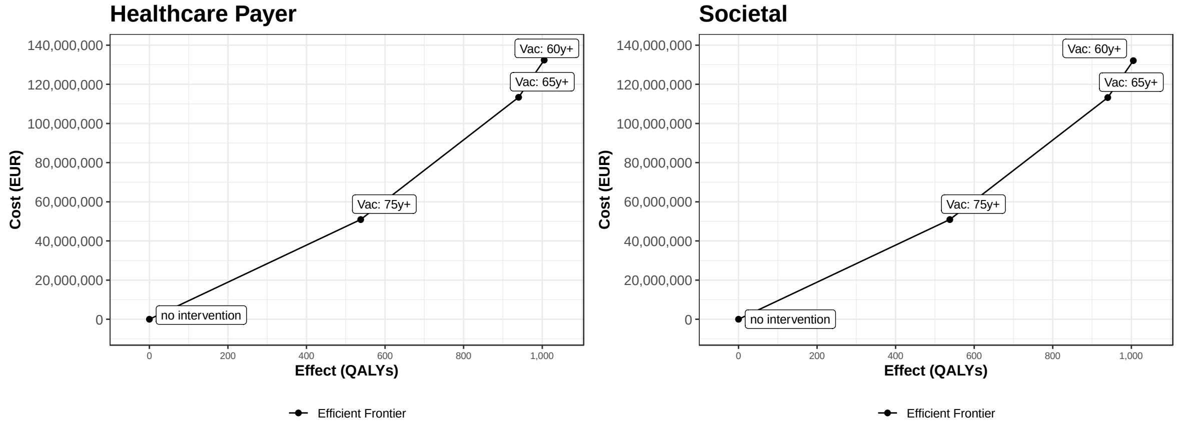

Spain-Valencia

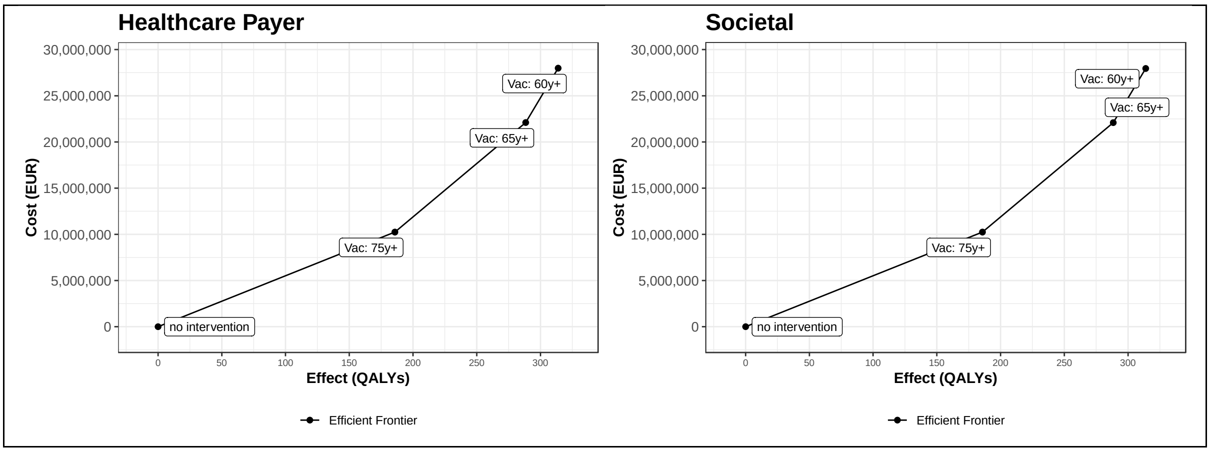

*Abbreviations: QALY: quality-adjusted life-year, EUR: euro*

##### 2.2.3.2 Cost-effectiveness acceptability curves (CEACs) and expected net loss curves (ENLCs)

For each strategy, the probability of being cost-effective is shown using CEACs (S. Figure 9 left plots). For Denmark, the results are comparable with using adjusted-ICD-coded hospitalisations. For Finland, the 75y+ strategy becomes cost-effective at lower WTP values (€95,000 per QALY gained) compared to adjusted ICD-coded analysis (€125,000). For Spain-Valencia, both 75y+ and 65y+ strategies reach probability of being 50% cost-effective. For WTP values below €40,000 per QALY gained no intervention is the preferred strategy with 100% certainty, whereas for WTP values of €80,000 per QALY gained, the 75y+ strategy is preferred with 95% certainty.

NLCs show the expected net loss (i.e., the expected cost of uncertainty) for each strategy. The expected net loss (cost of decision uncertainty) reaches peak for Finland at a lower WTP value than using adjusted RSV-ICD-coded hospitalisation. In Spain-Valencia, the expected net loss is below €0.1million.

*S. Figure 12: Using the adjusted RSV-confirmed data: Cost-effectiveness acceptability curves (left plots) and expected net loss curves (right plots) for each country comparing 3 strategies against RSV diseases in older adults: 60 years and above, 65 years and above and 75 years and above. All strategies were compared to no intervention and to each other. RSV-confirmed hospitalisation with adjustment factor were used for all countries. The assumed RSV vaccine price per dose is €150. The results are from healthcare payers' perspective.*

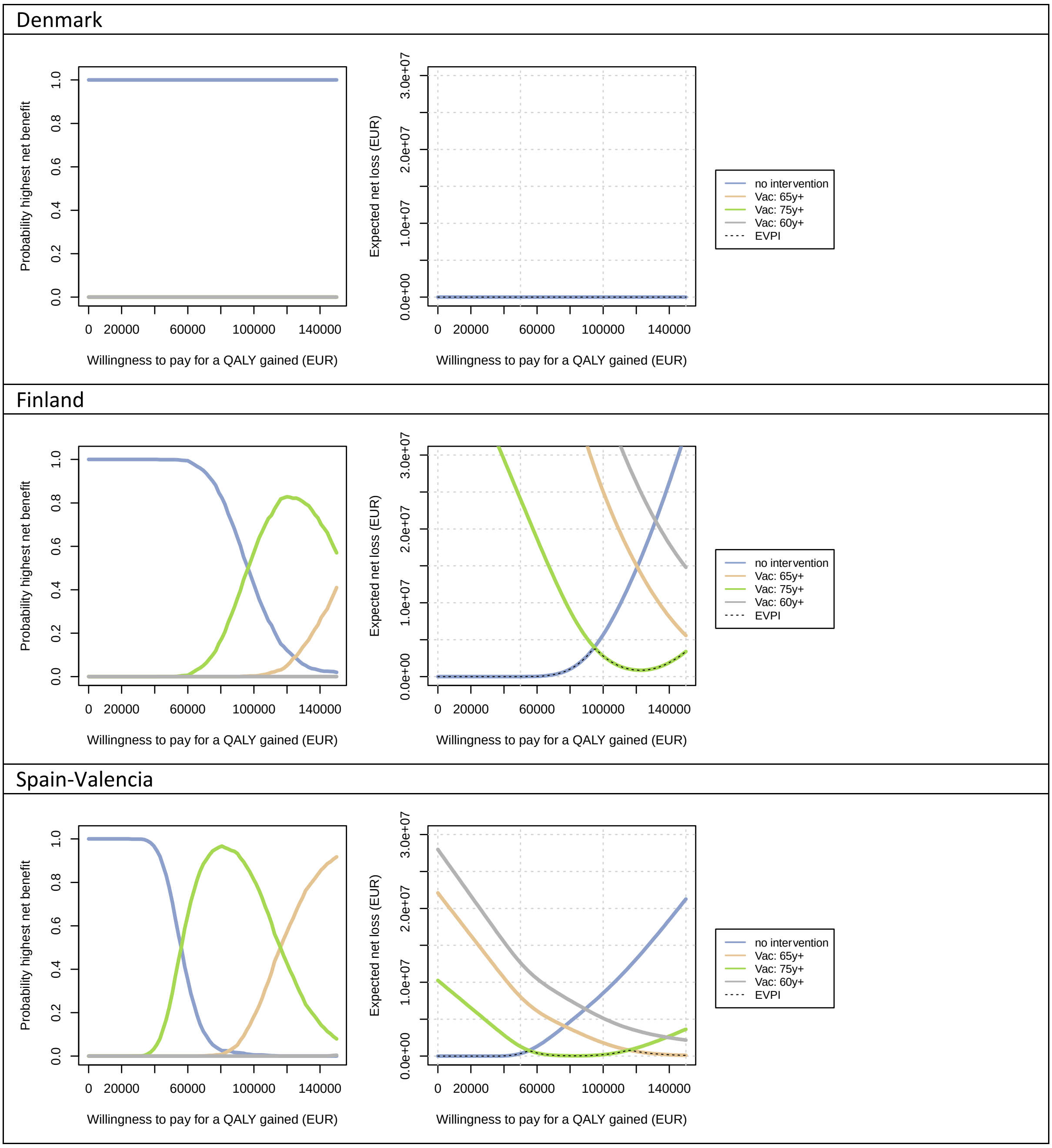

*Abbreviations: QALY: quality-adjusted life-year, EUR: euro*

##### 2.2.3.3 Expected value of partial perfect information (EVPPI)

Using adjusted RSV-confirmed data, no intervention is cost-effective in Denmark up to a WTP value of €150,000 per QALY gained with 100% certainty, hence, the EVPPI graph is still flat. In Finland and Spain-Valencia, the top five influential drivers are: uncertainty around age-specific hCFR, vaccine efficacy against mortality (both first and second year), the adjustment factors and QALY losses due to non-MA cases (S. Figure 13 and main text Figure 6).

*S. Figure 13: Using adjusted RSV-confirmed data: the expected value of partial perfect information for each country (Finland's graph is reported in the main text). The assumed RSV vaccine price per dose is €150. The results are from healthcare payers' perspective.*

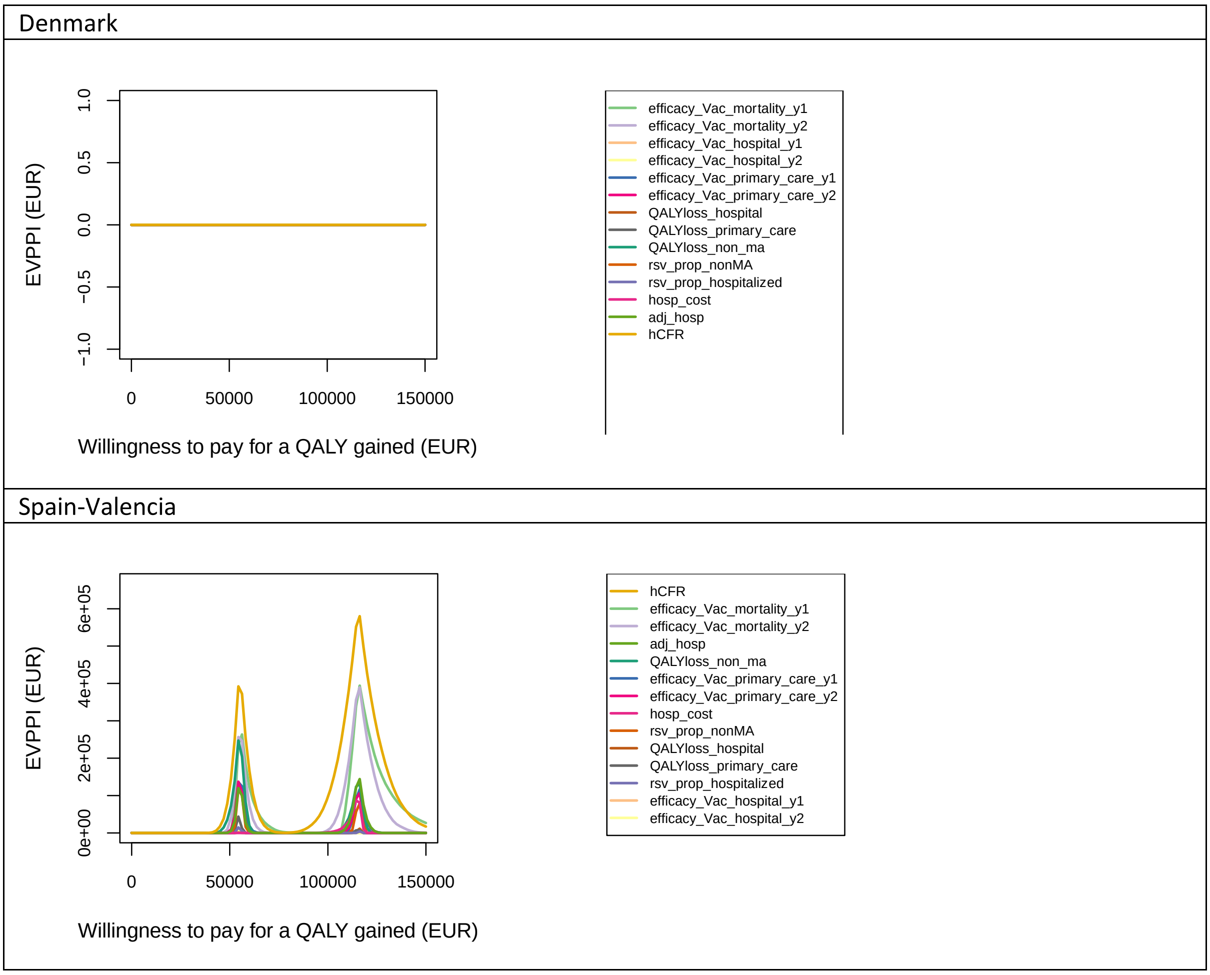

*Abbreviations: QALY: quality-adjusted life-year, EUR: euro, hCFR: in-hospital case fatality ratio, adj: adjustment factor, hosp: hospitalisation, RSV: respiratory syncytial virus, ma: medically attended, y: year*

#### 2.3 Using time-series modelled hospitalisation estimates

##### 2.3.1 RSV-attributable disease and economic burden without vaccination

*S. Table 12: Mean [95% CI] of RSV-attributable disease and economic burden in adults over 60 years for each country without intervention over a 5-year time horizon. All results are from the healthcare payers' perspective unless otherwise stated. Both costs and quality-adjusted life-years were discounted at country-specific discount rates. All costs are presented in €'000 in 2023 value.*

| Denmark | | | | | |
|---|---|---|---|---|---|
| | 60-64 years | 65-74 years | 75-84 years | 85 years + | 60 years + |
| Undiscounted outcomes | | | | | |
| Non-MA episodes | 34 834 [17182 - 54919] | 146 436 [72232 - 230866] | 180 780 [89169 - 285005] | 152 807 [75366 - 240913] | 514 858 [253949 - 811704] |
| Primary care episodes | 13 747 [11408 - 16029] | 57 789 [47959 - 67384] | 71 342 [59210 - 83195] | 60 303 [50048 - 70318] | 203 181 [168625 - 236926] |
| Hospitalisations | 1 615 [1615 - 1615] | 6 788 [6788 - 6789] | 8 381 [8380 - 8381] | 7 084 [7083 - 7085] | 23 867 [23866 - 23869] |
| Deaths | 116 [86 - 151] | 489 [361 - 636] | 604 [445 - 785] | 510 [376 - 663] | 1719 [1268 - 2235] |
| QALY losses | 2 284 [1730 - 2975] | 7 128 [5434 - 9277] | 5 482 [4140 - 7320] | 2 611 [1857 - 3800] | 17 505 [13301 - 22953] |
| Direct medical costs ('000 €) | 6 282 [6158 - 6421] | 26 408 [25887 - 26991] | 32 601 [31958 - 33321] | 27 556 [27015 - 28163] | 92 847 [91017 - 94895] |
| Costs of productivity loss ('000 €) | 6 393 [4803 - 8246] | 3 734 [2806 - 4817] | 0 | 0 [ | 10 128 [7609 - 13063] |
| Total costs from societal perspective ('000 €) | 12 675 [10960 - 14656] | 30 142 [28701 - 31782] | 32 601 [31958 - 33321] | 27 556 [27015 - 28163] | 102 974 [98663 - 107878] |
| Discounted costs and QALYs | | | | | |
| QALY losses | 1 703 [1297 - 2216] | 5 754 [4392 - 7527] | 4 862 [3639 - 6582] | 2 487 [1751 - 3659] | 14 806 [11159 - 19838] |
| Direct medical costs ('000 €) | 6 252 [6129 - 6390] | 26 241 [25724 - 26821] | 32 378 [31740 - 33093] | 27 344 [26806 - 27946] | 92 215 [90399 - 94250] |
| Costs of productivity loss ('000 €) | 6 363 [4780 - 8207] | 3 709 [2786 - 4783] | 0 [ | 0 [ | 10 072 [7567 - 12991] |
| Total costs from societal perspective ('000 €) | 12 615 [10908 - 14587] | 29 950 [28518 - 31579] | 32 378 [31740 - 33093] | 27 344 [26806 - 27946] | 10 2287 [98002 - 107161] |
| Finland | | | | | |
| | 60-64 years | 65-74 years | 75-84 years | 85years + | 60 years + |
| Undiscounted outcomes | | | | | |
| Non-MA episodes | 10 205 [5034 - 16092] | 53 419 [26353 - 84226] | 80 155 [39538 - 126368] | 57 134 [28179 - 90078] | 200 914 [99104 - 316764] |
| Primary care episodes | 4 027 [3341 - 4695] | 21 081 [17493 - 24576] | 31 632 [26253 - 36887] | 22 547 [18713 - 26291] | 79 288 [65801 - 92448] |
| Hospitalisations | 473 [473 - 473] | 2 476 [2476 - 2477] | 3 716 [3715 - 3716] | 2 649 [2648 - 2649] | 9 314 [9312 - 9315] |
| Deaths | 34 [25 - 44] | 178 [132 - 232] | 268 [197 - 348] | 191 [141 - 248] | 671 [495 - 872] |
| QALY losses | 612 [465 - 796] | 2 311 [1767 - 3014] | 2 082 [1555 - 2830] | 853 [587 - 1287] | 5 858 [4418 - 7845] |

| | | | | | |
|---|---|---|---|---|---|
| Direct medical costs ('000 €) | 1 740 [660 - 4107] | 10 446 [3725 - 29264] | 15 864 [5797 - 40380] | 9 428 [3361 - 25655] | 37 479 [19993 - 66681] |
| Costs of productivity loss ('000 €) | 1 291 [970 - 1665] | 731 [549 - 943] | 0 | 0 | 2 022 [1519 - 2608] |
| Total costs from societal perspective ('000 €) | 3 031 [1865 - 5469] | 11 177 [4415 - 29924] | 15 864 [5797 - 40380] | 9 428 [3361 - 25655] | 39 501 [21892 - 68660] |
| Discounted costs and QALYs | | | | | |
| QALY losses | 478 [365 - 623] | 1 937 [1475 - 2537] | 1 888 [1402 - 2605] | 818 [556 - 1247] | 5 122 [3825 - 6973] |
| Direct medical costs ('000 €) | 1 733 [658 - 4092] | 10 392 [3706 - 29111] | 15 773 [5763 - 40147] | 9 363 [3338 - 25478] | 37 261 [19880 - 66276] |
| Costs of productivity loss ('000 €) | 1 287 [967 - 1659] | 727 [546 - 937] | 0 | 0 | 2 013 [1513 - 2596] |
| Total costs from societal perspective ('000 €) | 3 020 [1858 - 5449] | 11 119 [4392 - 29768] | 15 773 [5763 - 40147] | 9 363 [3338 - 25478] | 39 275 [21767 - 68261] |
| The Netherlands | | | | | |
| | 60-64 years | 65-74 years | 75-84 years | 85years + | 60 years + |
| Undiscounted outcomes | | | | | |
| Non-MA episodes | 68 630 [33851 - 108198] | 303 132 [149519 -477898] | 340 092 [167747 -536159] | 175 059 [86343 - 275987] | 886 913 [437460 -1398242] |
| Primary care episodes | 27 084 [22477 - 31582] | 119 626 [99280 - 139496] | 134 212 [111389 -156514] | 69 084 [57336 - 80562] | 350 007 [290482 - 408154] |
| Hospitalisations | 3 181 | 14 052 | 15 766 | 8 115 | 41 115 |
| Deaths | 229 [169 - 298] | 1012 [747 - 1316] | 1135 [838 - 1476] | 584 [431 - 760] | 2 961 [2185 - 3850] |
| QALY losses | 4 894 [3706 - 6388] | 16 396 [12461 - 21352] | 12 185 [9260 - 15962] | 3 678 [2688 - 5127] | 37 153 [28412 - 48559] |
| Direct medical costs ('000 €) | 28 350 [28088 - 28643] | 117 702 [116545- 118994] | 122 215 [120920- 123664] | 53 138 [52475 - 53883] | 321 405 [318024 - 325184] |
| Costs of productivity loss ('000 €) | 31 010 [23195 - 40119] | 17 493 [13084 - 22632] | 0 | 0 | 48 503 [36279 - 62751] |
| Total costs from societal perspective ('000 €) | 59 360 [51280 - 68738] | 135 195 [129664- 141543] | 122 215 [120920 -123664] | 53 138 [52475 - 53883] | 369 908 [354395 - 387713] |
| Discounted costs and QALYs | | | | | |
| QALY losses | 4 248 [3220 - 5526] | 14 756 [11246 - 19198] | 11 422 [8682 - 14978] | 3 567 [2595 - 4982] | 33 992 [25958 - 44487] |
| Direct medical costs ('000 €) | 28 241 [27980 - 28533] | 117 062 [115912- 118347] | 121 458 [120171 -122898] | 52 782 [52123 - 53521] | 319 543 [316181 - 323299] |
| Costs of productivity loss ('000 €) | 30 891 [23105 - 39965] | 17 389 [13006 - 22497] | 0 | 0 | 48 280 [36112 - 62462] |
| Total costs from societal perspective ('000 €) | 59 132 [51082 - 68474] | 134 451 [128952- 140762] | 121 458 [120171 -122898] | 52 782 [52123 - 53521] | 367 823 [352385 - 385541] |
| Spain-Valencia | | | | | |
| | 60-64 years | 65-74 years | 75-84 years | 85years + | 60 years + |
| Undiscounted outcomes | | | | | |
| Non-MA episodes | 1 627 [804 - 2568] | 6 782 [3351 - 10703] | 11 231 [5541 - 17703] | 6 657 [3283 - 10499] | 26 298 [12978 - 41473] |
| Primary care episodes | 642 [533 - 748] | 2 677 [2221 - 3120] | 4 432 [3679 - 5169] | 2 627 [2179 - 3063] | 10 378 [8612 - 12096] |
| Hospitalisations | 75 [75 - 76] | 314 [314 - 315] | 521 [520 - 521] | 309 [308 - 309] | 1219 [1218 - 1220] |
| Deaths | 5 [4 - 7] | 23 [17 - 29] | 37 [28 - 49] | 22 [16 - 29] | 88 [65 - 114] |

| | | | | | |
|---|---|---|---|---|---|
| QALY losses | 116 [88 - 152] | 347 [264 - 452] | 355 [268 - 470] | 117 [84 - 169] | 935 [710 - 1223] |
| Direct medical costs ('000 €) | 442 [61 - 2029] | 1480 [510 - 4068] | 2330 [1023-4942] | 1358 [696 - 2534] | 5610 [3333 - 9342] |
| Costs of productivity loss ('000 €) | 103 [75 - 136] | 25 [18 - 33] | 0 | 0 | 128 [94 - 169] |
| Total costs from societal perspective ('000 €) | 545 [157 - 2145] | 1 505 [540 - 4091] | 2 330 [1023 - 4942] | 1 358 [696 - 2534] | 5 739 [3454 - 9505] |
| Discounted costs and QALYs | | | | | |
| QALY losses | 88 [67 - 115] | 286 [219 - 373] | 316 [238 - 425] | 111 [79 - 162] | 802 [606 - 1069] |
| Direct medical costs ('000 €) | 440 [60 - 2020] | 1 473 [508 - 4047] | 2 317 [1017 - 4913] | 1 348 [691 - 2516] | 5 578 [3313 - 9287] |
| Costs of productivity loss ('000 €) | 103 [75 - 135] | 25 [18 - 33] | 0 | 0 | 128 [93 - 168] |
| Total costs from societal perspective ('000 €) | 543 [157 - 2135] | 1 498 [538 - 4070] | 2 317 [1017 - 4913] | 1 348 [691 - 2516] | 5 706 [3433 - 9449] |

*Abbreviations: QALY: quality-adjusted life-year, MA: medically attended*

##### 2.3.2 Effects of vaccination on RSV-attributable disease and economic burden, and the associated costs

The RSV-attributable disease and economic burden averted by each strategy compared to no intervention and the associated intervention costs are presented in S. Table 13 for each country using TSM estimates. Given the differences in baseline disease burden estimates, the RSV-attributable disease burdens averted were much higher than the adjusted RSV-ICD-coded and adjusted RSV-confirmed disease burdens averted for all three strategies in Denmark and the Netherlands. In Finland, the disease burden averted were lower when using TSM estimates than using adjusted RSV-confirmed hospitalisation. In Spain-Valencia, the baseline disease burdens were lower when using TSM estimates than the adjusted RSV-confirmed data. Consequently, the direct and indirect costs averted and QALYs gained were also lower when using the TSM estimates.

*S. Table 13: Mean [95% CI] discounted disease and economic burden averted over 2 years protection (truncated linear waning, 24 months protection) for vaccinating 60 years and above, 65 years and above and 75 years and above in October against RSV disease in adults compared to no intervention for each country. TSM hospitalisation estimates were used for all countries. Both costs and quality-adjusted life-years were discounted at country-specific discount rates. All costs are presented in €'000 in 2023 value. All the results are from the healthcare payers' perspective unless otherwise stated. The RSV vaccine price per dose assumed is €150.*

| Denmark | 60 years + | 65 years + | 75 years + |
|---|---|---|---|
| non-MA cases averted | 87 403 [33969 - 163376] | 86 300 [33558 - 161256] | 64 026 [24963 - 119388] |
| Primary care episode averted | 34 408 [18397 - 52025] | 33 974 [18175 - 51343] | 25 206 [13519 - 37999] |
| Hospitalisation averted | 5 265 [3662 - 6463] | 5 197 [3617 - 6375] | 3 847 [2689 - 4709] |
| Death averted | 415 [266 - 565] | 410 [263 - 557] | 303 [194 - 412] |
| QALY gained due to RSV non-MA episode averted | 346 [54 - 1075] | 342 [53 - 1062] | 254 [40 - 787] |
| QALY gained due to RSV MA episode averted | 190 [79 - 417] | 187 [78 - 412] | 139 [58 - 306] |
| QALY gained due to death averted | 2 543 [1630 - 3458] | 2 474 [1586 - 3364] | 1 405 [900 - 1908] |
| Total QALYs gained | 3 079 [2062 - 4228] | 3 003 [2012 - 4126] | 1 797 [1200 - 2515] |
| Direct medical costs averted ('000€) | 19 799 [14274 - 24070] | 19 542 [14100 - 23746] | 14 474 [10483 - 17545] |
| Intervention costs ('000€) | 195 469 | 178 006 | 94 757 |
| Incremental costs ('000€) | 175 671 [171400 - 181196] | 158 464 [154260 - 163906] | 80 283 [77212 - 84274] |
| Costs associated with productivity loss averted | 915 [557 - 1360] | 699 [428 - 1036] | 0 |
| Incremental costs from societal perspective ('000€) | 174 756 [170287 - 180298] | 157 764 [153403 - 163227] | 80 283 [77212 - 84274] |
| Finland | 60 years + | 65 years + | 75 years + |
| non-MA cases averted | 26 731 [10427 - 49832] | 26 462 [10337 - 49279] | 20 562 [8057 - 38210] |
| Primary care episode averted | 10 524 [5647 - 15857] | 10 417 [5598 - 15676] | 8 095 [4363 - 12146] |
| Hospitalisation averted | 1 606 [1123 - 1964] | 1 588 [1113 - 1940] | 1 231 [867 - 1499] |
| Death averted | 126 [81 - 172] | 125 [80 - 170] | 97 [62 - 132] |
| QALY gained due to RSV non-MA episode averted | 106 [17 - 329] | 105 [16 - 325] | 82 [13 - 253] |
| QALY gained due to RSV MA episode averted | 58 [24 - 128] | 57 [24 - 126] | 45 [19 - 98] |
| QALY gained due to death averted | 656 [421 - 892] | 639 [410 - 869] | 381 [245 - 517] |
| Total QALYs gained | 820 [547 - 1144] | 802 [535 - 1118] | 507 [337 - 724] |
| Direct medical costs averted ('000€) | 6 077 [2897 - 11501] | 6 016 [2827 - 11448] | 4 591 [1999 - 9763] |
| Intervention costs ('000€) | 139 296 | 12 085 | 55 683 |
| Incremental costs ('000€) | 133 219 [127795 - 136398] | 114 070 [108637 - 117258] | 51 092 [45920 - 53684] |
| Costs associated with productivity loss averted | 141 [85 - 213] | 104 [64 - 154] | 0 |
| Incremental costs from societal perspective) ('000€) | 133 078 [127625 - 136295] | 113 965 [108511 - 117164] | 51 092 [45920 - 53684] |

| The Netherlands | 60 years + | 65 years + | 75 years + |
|---|---|---|---|
| non-MA cases averted | 141 518 [56436 - 259869] | 138 704 [55371 - 254527] | 95 572 [38282 - 174986] |
| Primary care episode averted | 55 721 [30562 - 82234] | 54 613 [29990 - 80520] | 37 631 [20752 - 55348] |
| Hospitalisation averted | 8 346 [6051 - 10002] | 8 173 [5936 - 9785] | 5 615 [4101 - 6701] |
| Death averted | 652 [424 - 877] | 638 [415 - 859] | 438 [286 - 589] |
| QALY gained due to RSV non-MA episode averted | 564 [90 - 1734] | 553 [88 - 1699] | 381 [61 - 1169] |
| QALY gained due to RSV MA episode averted | 308 [128 - 678] | 301 [125 - 664] | 208 [86 - 458] |
| QALY gained due to death averted | 5 769 [3752 - 7767] | 5 542 [3606 - 7458] | 2 954 [1924 - 3970] |
| Total QALYs gained | 6 640 [4478 - 8905] | 6 397 [4315 - 8580] | 3 543 [2420 - 4819] |
| Direct medical costs averted ('000€) | 62 581 [46329 - 74557] | 61 080 [45298 - 72692] | 40 129 [29919 - 47616] |
| Intervention costs ('000€) | 481 683 | 403 983 | 184 347 |
| Incremental costs ('000€) | 419 102 [407126 - 435354] | 342 903 [331291 - 358685] | 144 218 [136731 - 154428] |
| Costs associated with productivity loss averted | 4 395 [2706 - 6502] | 3 051 [1888 - 4477] | 0 |
| Incremental costs from societal perspective ('000€) | 414 707 [401778 - 431562] | 339 853 [327578 - 355904] | 144 218 [136731 - 154428] |
| Spain-Valencia | 60 years + | 65 years + | 75 years + |
| non-MA cases averted | 4 262 [1716 - 7773] | 4 134 [1666 - 7536] | 3 273 [1323 - 5952] |
| Primary care episode averted | 1 678 [930 - 2456] | 1 628 [903 - 2381] | 1 289 [717 - 1879] |
| Hospitalisation averted | 249 [184 - 296] | 242 [178 - 287] | 191 [141 - 226] |
| Death averted | 19 [13 - 26] | 19 [12 - 25] | 15 [10 - 20] |
| QALY gained due to RSV non-MA episode averted | 17 [3 - 52] | 16 [3 - 50] | 13 [2 - 40] |
| QALY gained due to RSV MA episode averted | 9 [4 - 20] | 9 [4 - 20] | 7 [3 - 16] |
| QALY gained due to death averted | 129 [84 - 173] | 120 [79 - 161] | 76 [50 - 103] |
| Total QALYs gained | 155 [106 - 211] | 146 [100 - 199] | 96 [66 - 134] |
| Direct medical costs averted ('000€) | 1 087 [612 - 1784] | 1 045 [583 - 1707] | 815 [450 - 1432] |
| Intervention costs ('000€) | 30 122 | 24 20 | 11 787 |
| Incremental costs ('000€) | 29 035 [28338 - 29510] | 23 076 [22413 - 23537] | 10 971 [10355 - 11337] |
| Costs associated with productivity loss averted | 13 [8 - 19] | 4 [3 - 7] | 0 |
| Incremental costs from societal perspective ('000€) | 29 022 [28327 - 29497] | 23 071 [22406 - 23533] | 10 971 [10355 - 11337] |

*Abbreviations: QALY: quality-adjusted life-year, MA: medically attended*

##### 2.3.3 Cost-effectiveness based on time-series modelled hospitalisation estimates

###### *2.3.3.1 Incremental cost-effectiveness plane*

From the HCP perspective, S. Figure 14 shows findings consistent with Figure 4 in the main text. From a societal perspective, the 75y+, 65y+ and 60y+ strategies are efficient when compared to no intervention and to each other in all four countries.

*S. Figure 14: Incremental cost-effectiveness plane from healthcare payers' (HCP; left) and societal (right) perspectives. TSM hospitalisation estimates were used for all countries. The assumed RSV vaccine price per dose is €150.*

Denmark

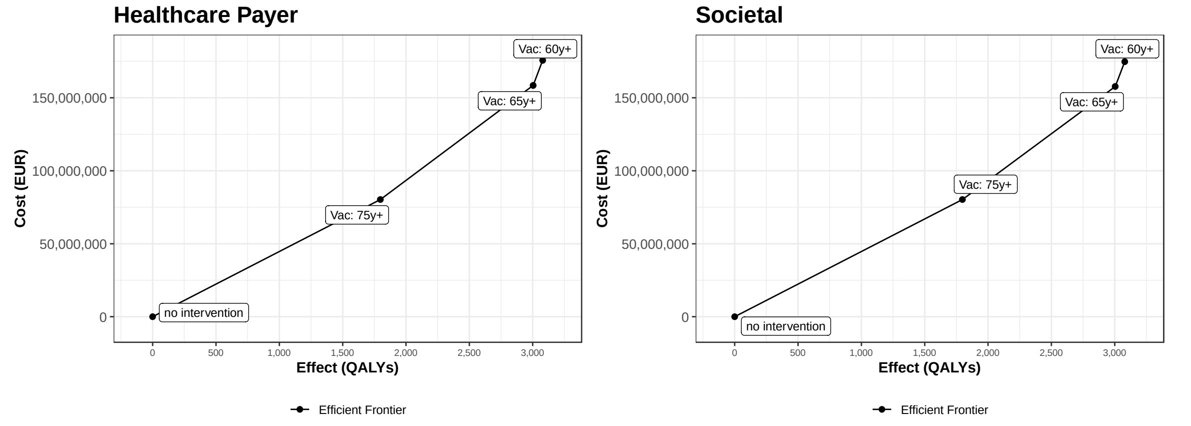

Finland

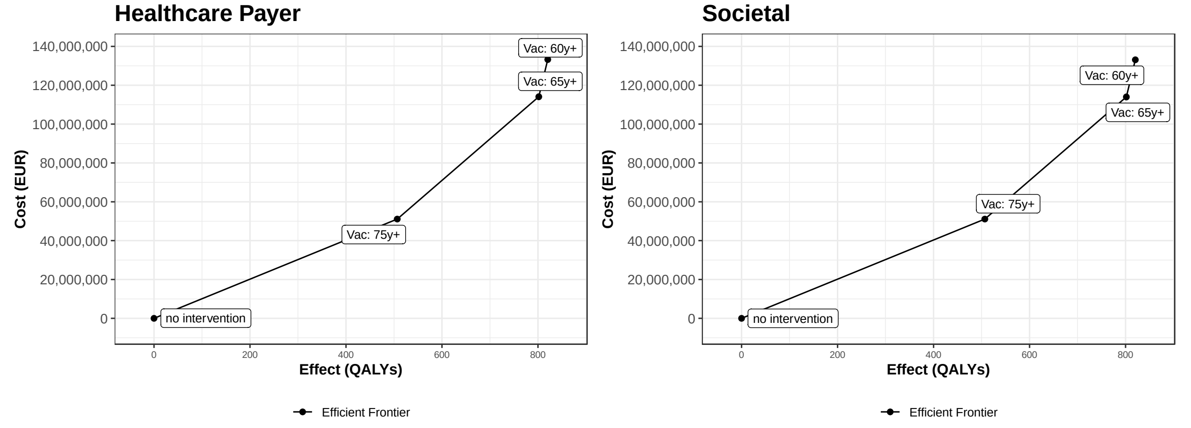

The Netherlands

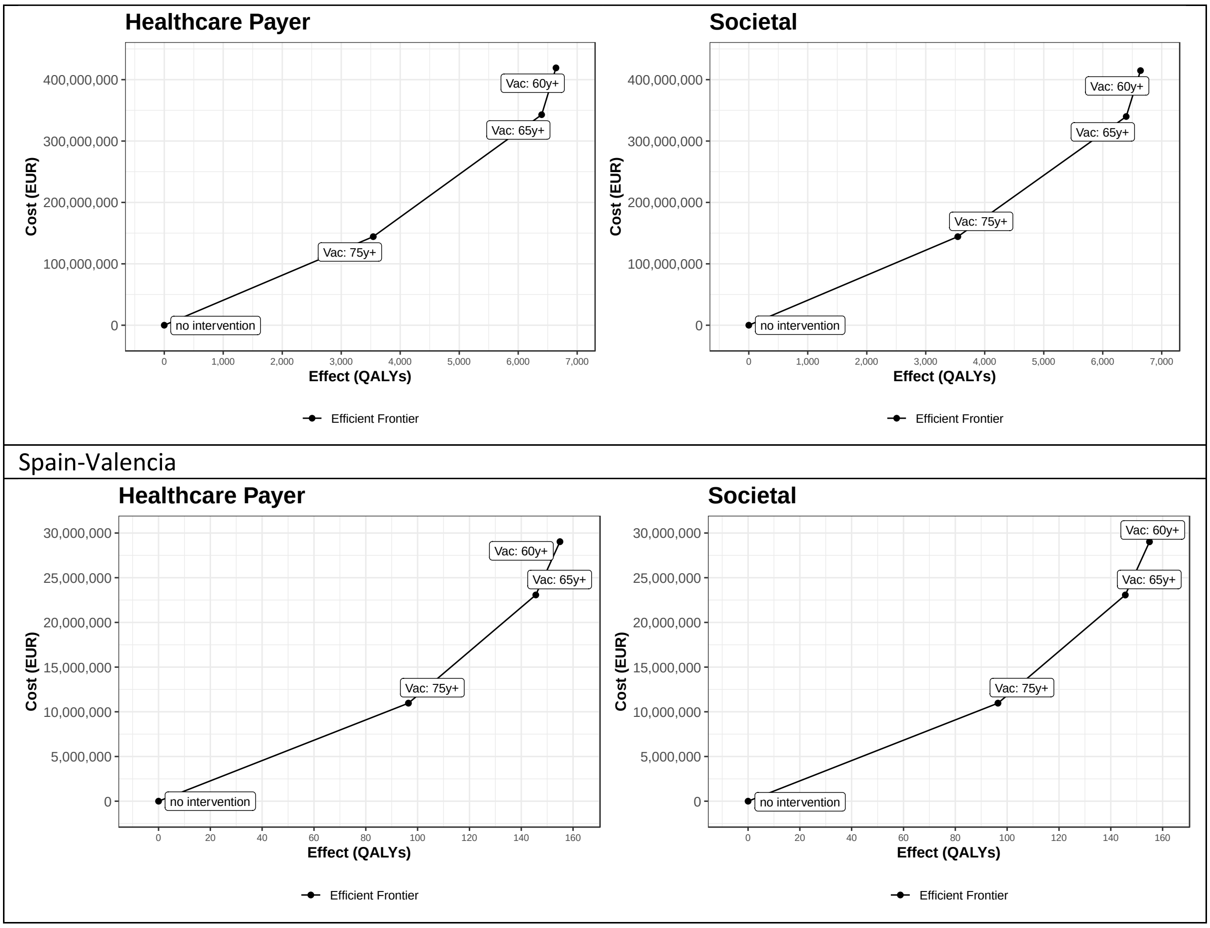

*Abbreviations: QALY: quality-adjusted life-year, EUR: euro*

##### *2.3.3.2 Cost-effectiveness acceptability curves (CEACs) and expected net loss curves (ENLCs)*

CEACs and ENLCs inform the uncertainty surrounding the cost-effectiveness results (S. Figure 15 left plots). For instance, for Denmark, when the WTP values is below €30,000 per QALY gained, “no intervention” is the preferred strategy with 100% certainty (i.e., probability to be cost-effective is 100%), whereas for WTP values above €110,000 per QALY gained, the 65y+ strategy is preferred with 100% certainty.

ENLCs show the expected net loss (i.e., the expected cost of uncertainty) for each strategy. For Spain-Valencia, the expected net loss stays well below €1 million for the entire range of WTP values considered. For Denmark, Finland and the Netherlands, the expected net loss reaches €6, €4 and €16 million, respectively. This either means that uncertain parameters are more influential for the cost-effectiveness of RSV vaccination in the Netherlands, or that generally more uncertainty was accounted for in the input data distributions that are specific to these countries.

*S. Figure 15: Using the TSM hospitalisation estimates: cost-effectiveness acceptability curves (left plots) and expected net loss curves (right plots) for each country comparing 3 strategies against RSV diseases in older adults: 60 years and above, 65 years and above and 75 years and above. All strategies were compared to no intervention and to each other. TSM estimates were used for all countries, except Spain-Valencia, where RSV-confirmed data were used based on their active surveillance database. The assumed RSV vaccine price per dose is €150. The results are from healthcare payers' perspective.*

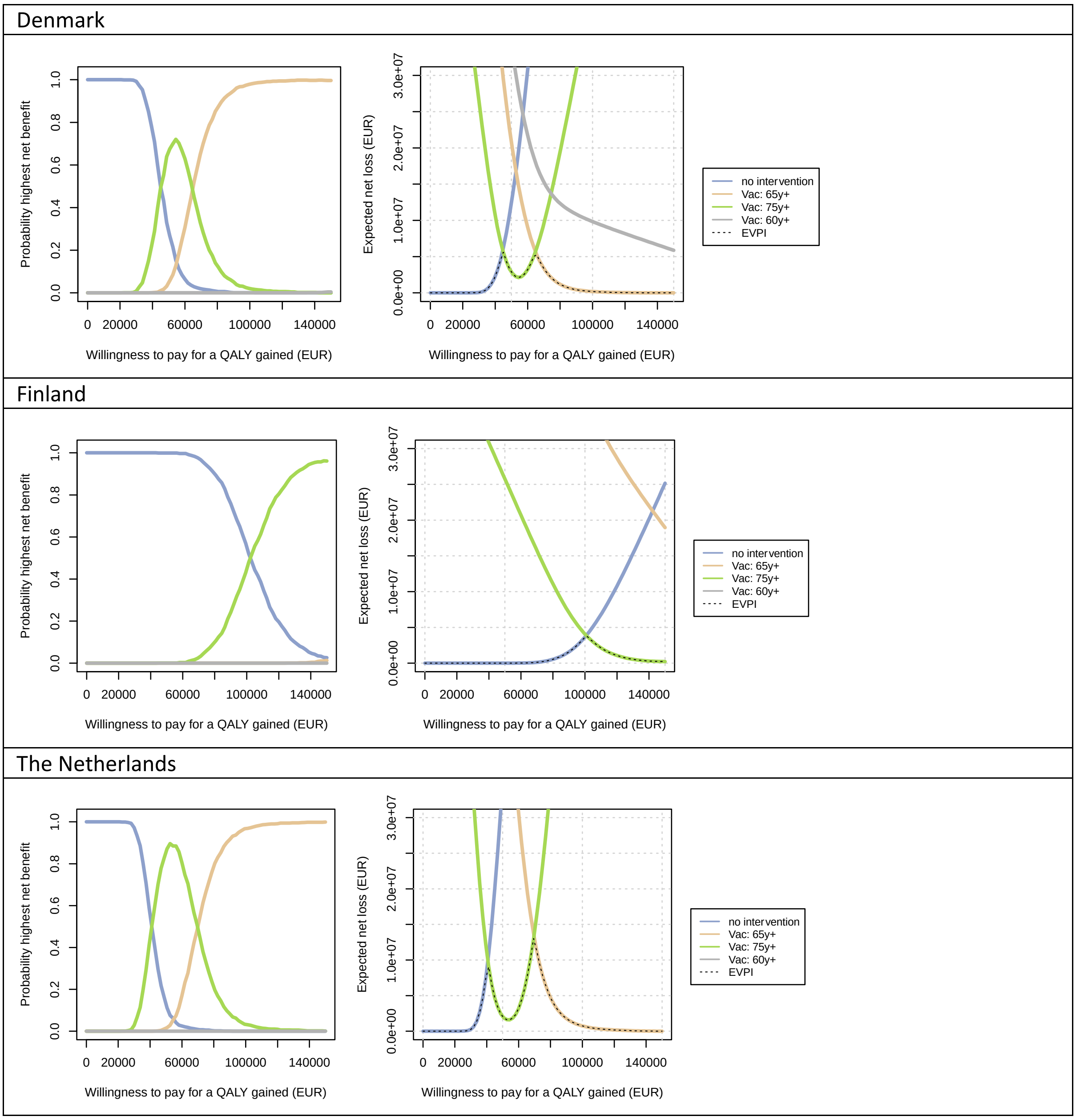

Spain-Valencia

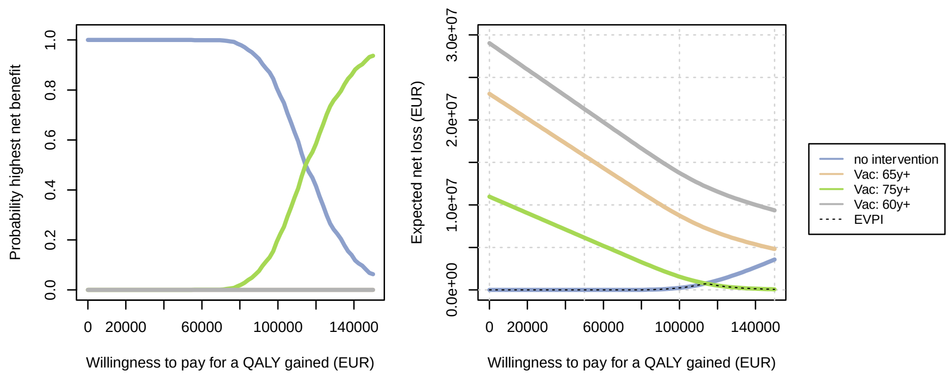

*Abbreviations: QALY: quality-adjusted life-year, EUR: euro*

##### *2.3.3.3 Expected value of partial perfect information (EVPPI)*

Using the TSM hospitalisation estimates, the uncertainty around non-age-specific hCFR caused most decision uncertainty (i.e., with highest EVPPI; S. Figure 16, main text Figure 6) for all countries. The vaccine efficacy against against mortality (both first and second year), QALY losses of non-MA cases for all countries are also the top influential drivers.

*S. Figure 16: The expected value of partial perfect information for each country (Finland's graph is reported in the main text). The assumed RSV vaccine price per dose is €150. The results are from healthcare payers' perspective.*

Denmark

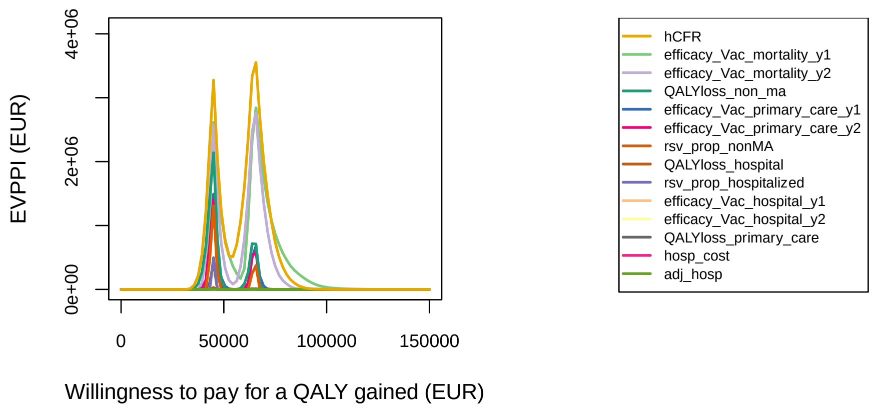

The Netherlands

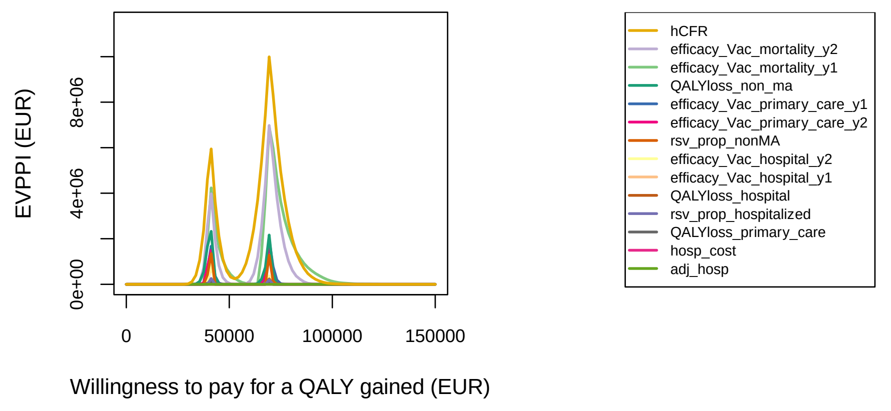

Spain-Valencia

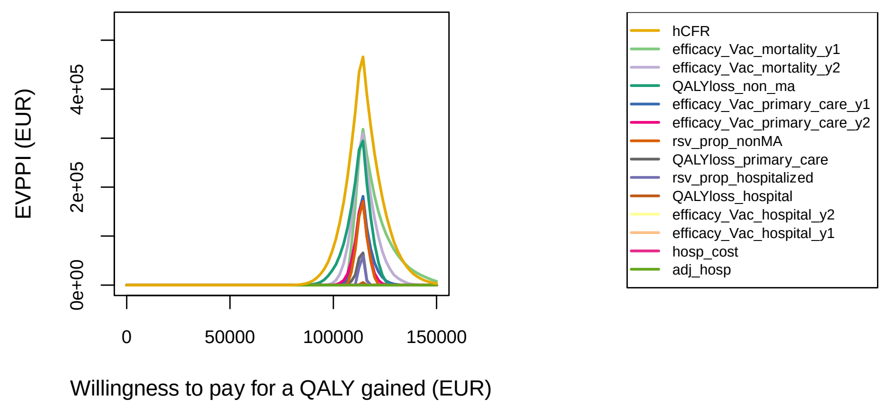

*Abbreviations: QALY: quality-adjusted life-year, EUR: euro, hCFR: in-hospital case fatality ratio, adj: adjustment factor, hosp: hospitalisation, RSV: respiratory syncytial virus, ma: medically attended, y: year*

#### 2.4 Price threshold analysis

Figure 5 in main text, S. Figure 17 and S. Figure 18 present the preferred RSV vaccination strategy at different vaccine prices for a range of WTP values per QALY gained from the HCP perspective of each country. They illustrate that the preferred strategy depends heavily on the intervention's price for a given WTP value.

Using adjusted RSC-ICD-coded hospitalisations, S. Figure 17 shows consistent findings. In Denmark, at any price, 'no intervention' would be cost-effective at WTP value up to €150,000 per QALY gained. If the vaccine is priced at €50 per dose, the 75y+ strategy can be cost-effective at WTP of €80,000 in the Netherlands, €40,000 in Finland, and the 65y+ strategy would become cost-effective at higher WTP ranges. However, when the vaccine is priced at €250 per dose, no intervention would be cost-effective at €250 per dose for any WTP value up to €150,000 per QALY gained.

Using the adjusted RSV-confirmed data, the price threshold analysis in S. Figure 18 shows consistent findings as using the RSV-ICD-coded data in Denmark. In Finland, the WTP values were €10 000 lower for 75y+ strategy become preferred at vaccine price of €50 per dose. In Spain-Valencia, at €50 per dose, the 65y+ strategy would be cost-effective at a WTP value of €55,000 per QALY gained, whereas the 60y+ strategy would be cost-effective at a WTP value of €90,000 per QALY gained.

Using TSM-estimates, the plot and the interpretation are shown in the main text (Figure 5).

*S. Figure 17: Using adjusted ICD-coded hospitalisation estimates: Price threshold analysis for different RSV vaccine prices for a range of willingness-to-pay values (from €0 to €80,000 per QALY gained) from healthcare payers' perspective (€ 2023 value). The assumed RSV vaccine price per dose is €150. The results are from healthcare payers' perspective.*

| Denmark | Finland |
|---|---|

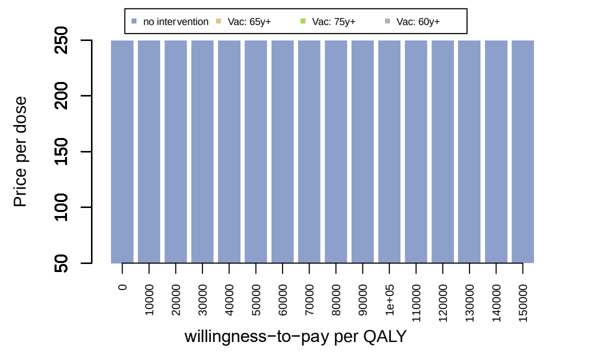

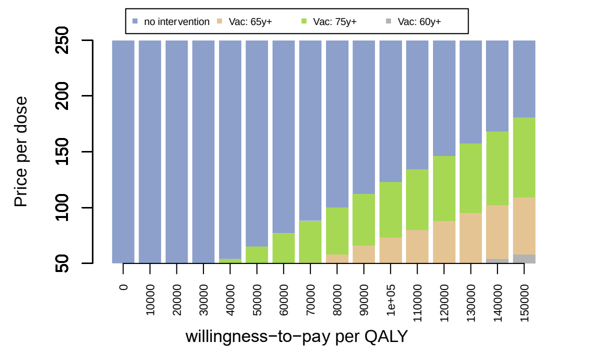

The Netherlands

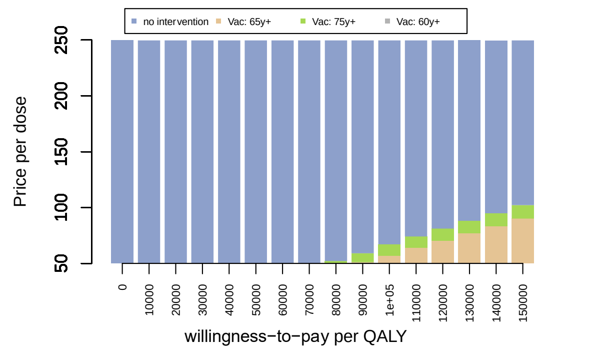

*S. Figure 18: Using adjusted RSV-confirmed hospitalisations: Price threshold analysis for different RSV vaccine prices for a range of willingness-to-pay values (from €0 to €80,000 per QALY gained) from healthcare payers' perspective (€ 2023 value). The assumed RSV vaccine price per dose is €150. The results are from healthcare payers' perspective.*

Denmark

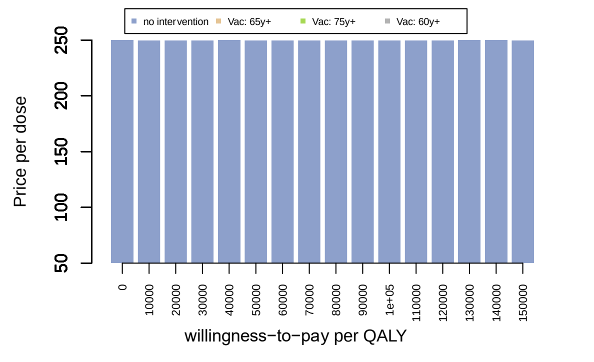

Finland

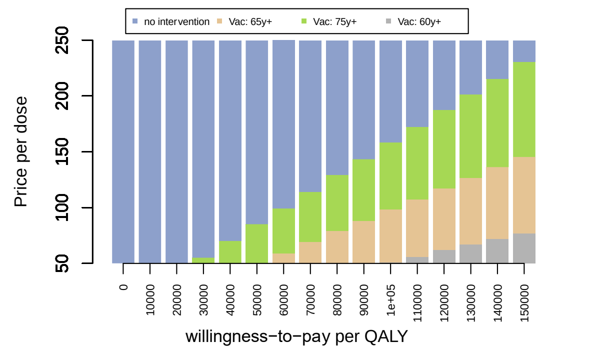

Spain-Valencia

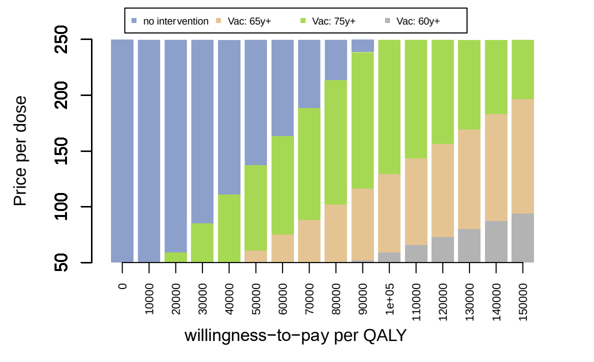

#### 2.5 Scenario analyses

##### 2.5.1 Results of the scenarios related to the assumed waning characteristics

Overall, assuming different waning characteristics had impacts on the cost-effectiveness results given we are only considering a single hypothetical vaccine. As shown in S. Figure 19, with longer duration of protection, the 75y+ strategy would become the preferred strategy at lower WTP values per QALY gained in all countries regardless of the shape of the waning curves. However, the differences between 24 months of protection and 48 months of protection are modest; this can most likely be attributed to a reduction in vaccine efficacy by one-third at the end of the second RSV season. Our assumed further reductions in RSV vaccine protective efficacy in the third and/or fourth seasons resulted in a relatively limited additional impact on disease burden.

It concludes that the assumed duration of protection tends to have more impact than the type of waning curve assumed. The exponential waning curves yielded the most favourable outcomes for all strategies.

*S. Figure 19: Using TSM estimates: outputs of scenarios related to the assumed waning characteristics: linear, exponential, 1-exponential (temporal in the graph), stepwise + linear and duration of protection: 24 month, 36 months and 48 months from healthcare payers' perspective. The assumed RSV vaccine price per dose is €150.*

| Denmark |
|---|

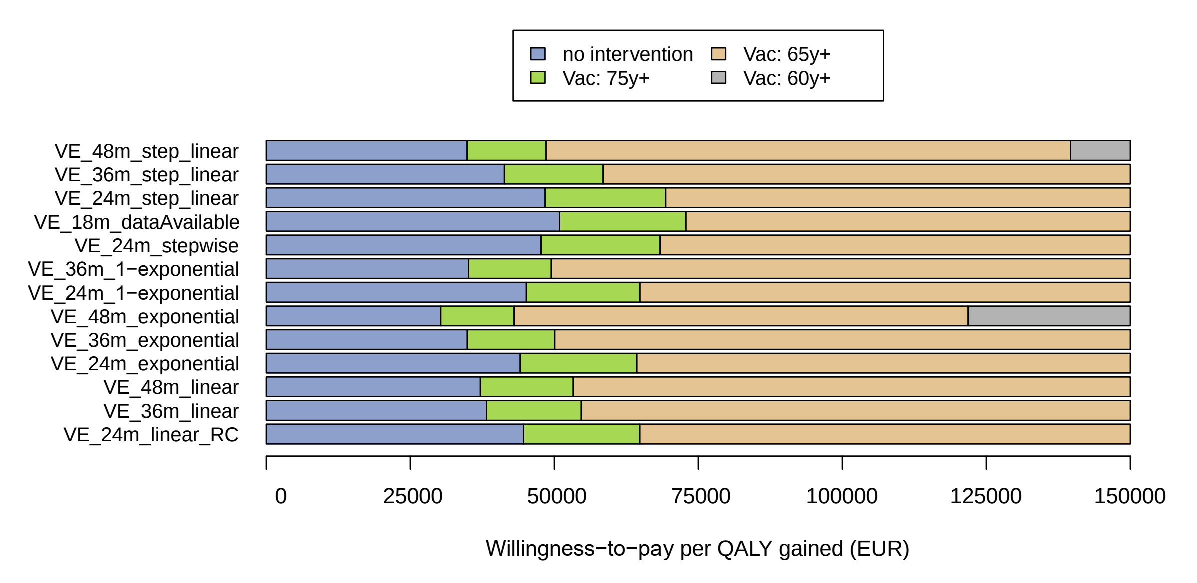

Finland

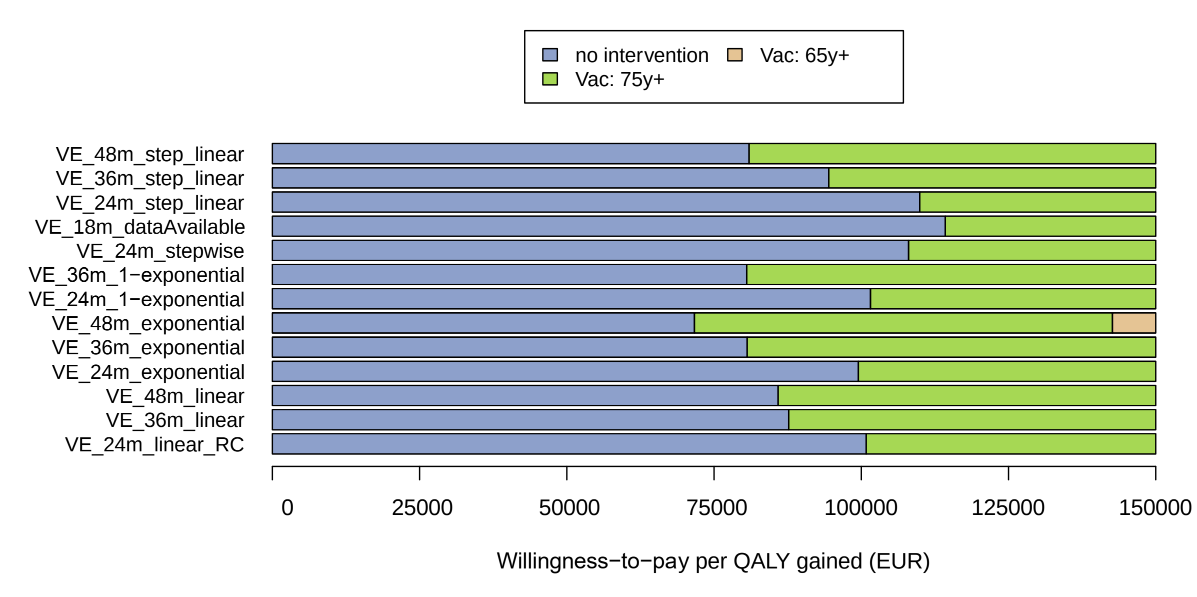

The Netherlands

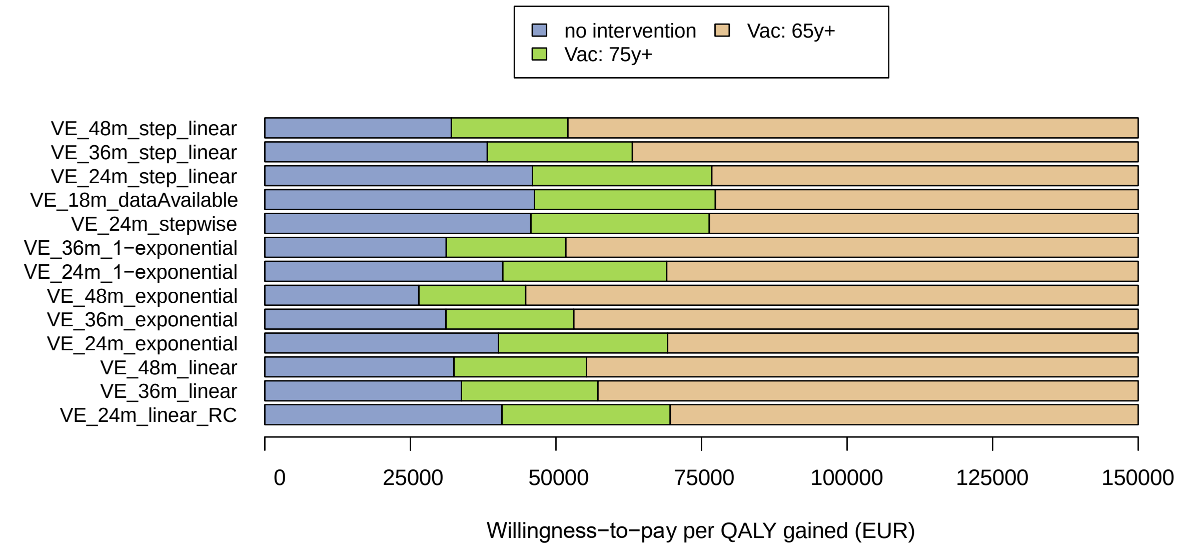

Spain-Valencia

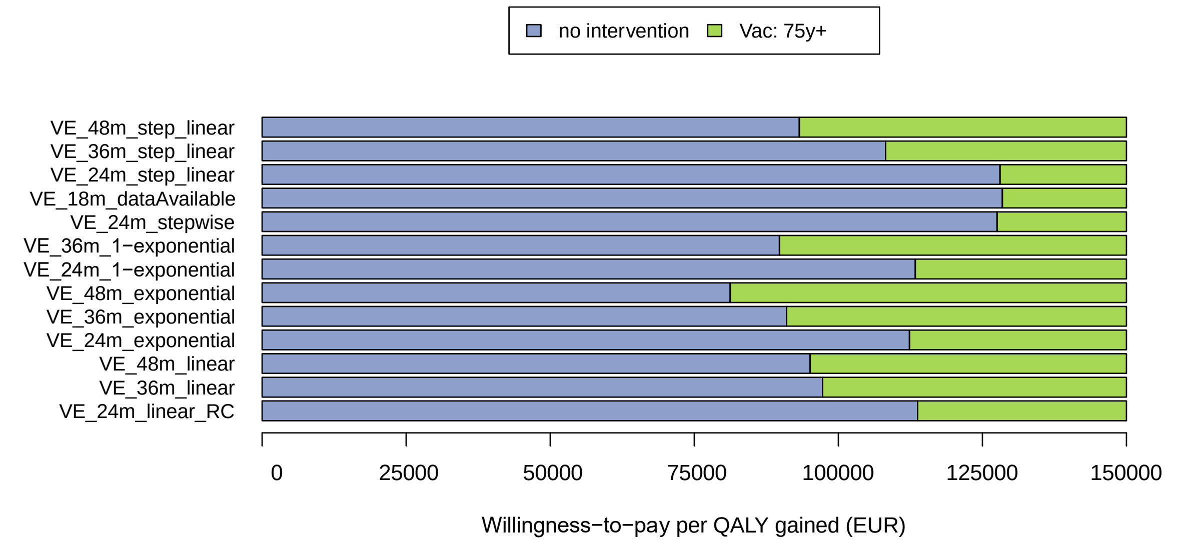

*Abbreviations: RC: reference scenario 0, m: month, VE: vaccine efficacy, DNK: Denmark, FIN: Finland, NLD: the Netherlands, ESP: Spain, EUR: euro, Vac: vaccine, QALY: quality-adjusted life-year*

##### 2.5.2 Results of scenarios unrelated to the assumed waning characteristics

In addition, assuming a higher QALY loss due to RSV illness and a higher primary care ratio would lead to more favourable results than the country reference analysis. In contrast, assuming 50% lower efficacy values in adults aged 80 years and above would have less favourable results in terms of the cost-effectiveness of RSV vaccination programmes, especially in Denmark, the 75y+ strategy was dominated by 65y+ strategy.

S. Figure 20 shows findings consistent with those in Figure 2-7 in the main text. It confirms that the decision uncertainty in all countries stems mostly from the uncertainty about which source data best reflects the incidence of RSV hospital admissions (RSV ICD-coded, RSV-confirmed and RSV attributable), and secondly, on the intervention's price. As discussed in the main text, the age-specific hCFR is the top-ranking influential driver.

Considering changes in the RSV seasonality, we performed a scenario simulating the seasonal shift as observed during the COVID-19 pandemic (peri-COVID_season in S. Figure 19). If the peak of the RSV season shifted 4 months earlier than the "typical" peak, it would lead to a relatively large negative impact on the RSV vaccination strategies in the Netherlands and Spain-Valencia, as administrating RSV vaccine in October might miss almost the entirety of the peak during the initial RSV season. However, the negative impact is smaller in Denmark and Finland, because of only a portion the peak might be missed.

If a biannual pattern of the "severe" and "mild" seasons would occur in all countries like Finland, introducing the RSV vaccination programme before the predicted "severe" season would lead to more favourable results than introducing it before the predicted "mild" season. However, our model did not evaluate the possibility of re-vaccination.

In addition, assuming a higher QALY loss due to RSV illness and a higher primary care ratio would lead to more favourable results than the country reference analysis. In contrast, assuming 50% lower efficacy values in adults aged 80 years and above would have less favourable results in terms of the cost-effectiveness of RSV vaccination programmes, especially in Denmark, the 75y+ strategy was dominated by 65y+ strategy.

*S. Figure 20: Outputs of scenarios unrelated to the assumed waning characteristics: the bottom bar in each graph represents the country reference analysis (TSM hospitalisation estimates in Denmark, Finland and the Netherlands, laboratory-confirmed hospitalisations in Spain-Valencia). The assumed RSV vaccine price per dose is €150. All results are from the healthcare payers' perspective unless otherwise stated*

Denmark

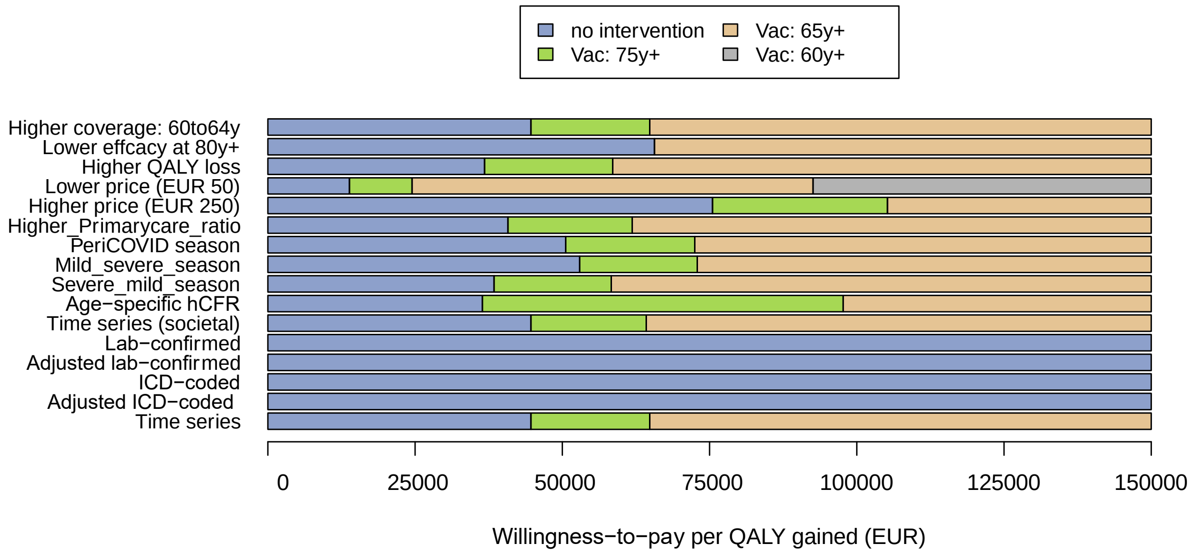

Finland

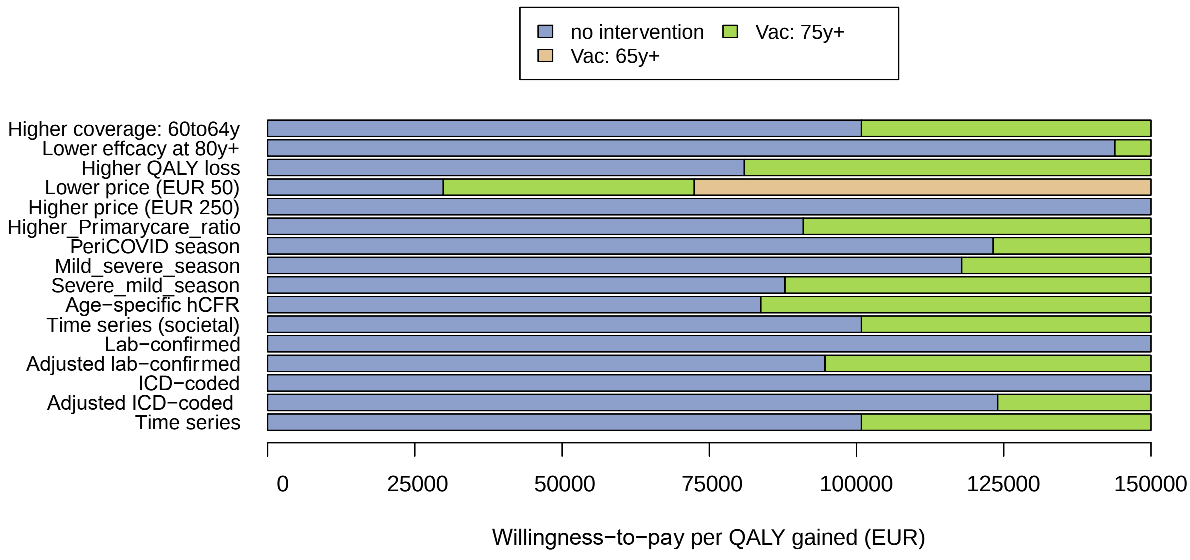

The Netherlands

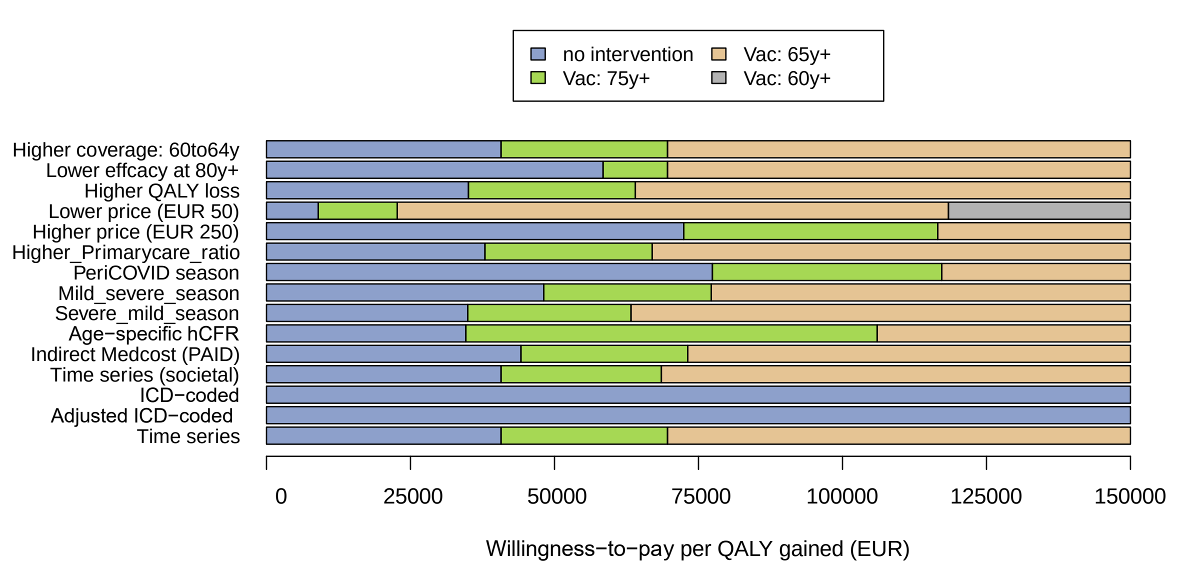

Spain-Valencia

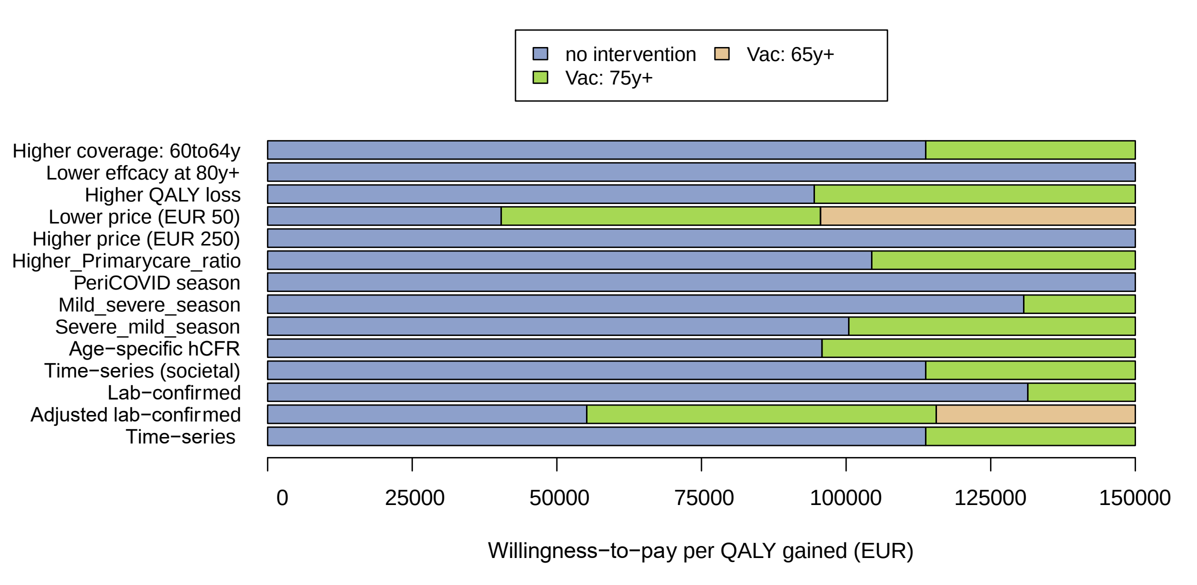

*Abbreviations: EUR: euro, QALY: quality-adjusted life-year, Vac: vaccine.*

33. **HICP - annual data (average index and rate of change)** [https://ec.europa.eu/eurostat/databrowser/view/prc_hicp_aind/default/table?lang=en&category=prc.prc_hicp]

34. GSK: **Arexvy product leaflet (ESWI conference)**. In. The European Scientific Working Group on Influenza (ESWI) 2023.

35. Drummond MF, Sculpher MJ, Claxton K, Stoddart GL, Torrance GW: **Methods for the Economic Evaluation of Health Care Programmes**, Fourth edn. Oxford: Oxford University Press; 2015.

36. **HONORARTABEL DAGTID Overenskomst om almen praksis** [https://laeger.dk/media/owclyvfi/honorartabel_2022_oktober.pdf]

37. Jacob J, Biering-Sorensen T, Holger Ehlers L, Edwards CH, Mohn KG, Nilsson A, Hjelmgren J, Ma W, Sharma Y, Ciglia E *et al*: **Cost-Effectiveness of Vaccination of Older Adults with an MF59((R))-Adjuvanted Quadrivalent Influenza Vaccine Compared to Standard-Dose and High-Dose Vaccines in Denmark, Norway, and Sweden**. *Vaccines (Basel)* 2023, **11**(4).

38. Mäklin S, Kokko P: **Terveyden- ja sosiaalihuollon yksikkökustannukset Suomessa vuonna 2017**. In.; 2020.

39. Nieminen H, Hakulinen T, Puumalainen T, Siren P, Palmu AA: **Time and labour costs of preventive health care, including vaccinations, in Finnish child health clinics**. *PLoS One* 2022, **17**(10):e0270835.

40. Hakkaart-van Roijen L, Peeters S, Kanters T: **Kostenhandleiding voor economische evaluaties in de gezondheidszorg: Methodologie en Referentieprijzen (Herziene versie 2024)**. In. Edited by Institute for Medical Technology Assessment Erasmus Universiteit Rotterdam. Online; 2024.

41. **Declareren vaccinaties** [https://www.snpg.nl/article/griep-ha-afronden/declareren-griepvaccinaties/vergoeding/#:~:text=U%20ontvangt%20een%20vergoeding%20voor,14%2C01%20per%20toegediend%20vaccin.]

48. **Consumer price index** [https://www.dst.dk/en/Statistik/emner/oekonomi/prisindeks/forbrugerprisindeks]

49. **HISB9: Life table (5 years tables) by sex, age and life table** [www.statbank.dk/HISB9]

50. **ECU/EUR exchange rates versus national currencies** [https://ec.europa.eu/eurostat/databrowser/view/tec00033/default/table?lang=en]

51. Heins M, Knottnerus B, Matser A, Hooiveld M: **Vaccine Coverage Dutch National Influenza Prevention Program 2022: brief monitor**. In.: Netherlands Institute for Health Services Research (NIVEL); 2023.

52. **Influenza season 2021/2022** [https://en.ssi.dk/surveillance-and-preparedness/surveillance-in-denmark/annual-reports-on-disease-incidence/influenza-season-2021-2022]

53. Alban A, Gyldmark M, Pedersen AV, Sogaard J: **The Danish approach to standards for economic evaluation methodologies**. *Pharmacoeconomics* 1997, **12**(6):627-636.

54. **Population by labour force status, sex and age, 2009-2023** [https://pxdata.stat.fi/PxWeb/pxweb/en/StatFin/StatFin__tyti/statfin_tyti_pxt_13aj.px/table/tableViewLayout1/]

55. **Deaths, Statistics Finland** [https://www.stat.fi/index_en.html]

56. Koskinen S, Lundqvist A, Ristiluoma N: **Health, functional capacity and welfare in Finland in 2011**. In. Helsinki, Finland: National Institute for Health and Welfare (THL); 2012: 290.

57. **Preparing a Health Economic Evaluation to Be Attached to the Application for Reimbursement Status and Wholesale Price for a Medicinal Product. Application Instructions (December 2019)** [https://tools.ispor.org/PEguidelines/source/2011Pricing_Board_Guidance_HEevaluation_english.pdf]

58. **Employment rates by sex, age and citizenship** [https://ec.europa.eu/eurostat/databrowser/view/lfsa_ergan/default/table?lang=en]

59. **Consumer prices** [https://www.cbs.nl/en-gb/figures/detail/83131ENG]

60. **Levensverwachting; geslacht, leeftijd (per jaar en periode van vijf jaren)** [https://opendata.cbs.nl/statline/#/CBS/nl/dataset/37360ned/table?dl=9F607]

61. **Life Tables** [https://www.ine.es/dyngs/INEbase/en/operacion.htm?c=Estadistica_C&cid=1254736177004&menu=resultados&idp=1254735573002]

62. **Marciano Gómez anima a la vacunación como medida más eficaz para protegerse frente a los virus respiratorios** [https://comunica.gva.es/es/detalle?id=377968997&site=373422400]

63. Lopez Bastida J, Oliva J, Antonanzas F, Garcia-Altes A, Gisbert R, Mar J, Puig-Junoy J: **[A proposed guideline for economic evaluation of health technologies]**. *Gac Sanit* 2010, **24**(2):154-170.

64. Alarid-Escudero F, Enns EA, Kuntz KM, Michaud TL, Jalal H: **"Time Traveling Is Just Too Dangerous" but Some Methods Are Worth Revisiting: The Advantages of Expected Loss Curves Over Cost-Effectiveness Acceptability Curves and Frontier**. *Value Health* 2019, **22**(5):611-618.

65. Bilcke J, Beutels P: **Generating, Presenting, and Interpreting Cost-Effectiveness Results in the Context of Uncertainty: A Tutorial for Deeper Knowledge and Better Practice**. *Med Decis Making* 2022, **42**(4):421-435.
